# A time-resolved proteomic and diagnostic map characterizes COVID-19 disease progression and predicts outcome

**DOI:** 10.1101/2020.11.09.20228015

**Authors:** Vadim Demichev, Pinkus Tober-Lau, Tatiana Nazarenko, Charlotte Thibeault, Harry Whitwell, Oliver Lemke, Annika Röhl, Anja Freiwald, Lukasz Szyrwiel, Daniela Ludwig, Clara Correia-Melo, Elisa T. Helbig, Paula Stubbemann, Nana-Maria Grüning, Oleg Blyuss, Spyros Vernardis, Matthew White, Christoph B. Messner, Michael Joannidis, Thomas Sonnweber, Sebastian J. Klein, Alex Pizzini, Yvonne Wohlfarter, Sabina Sahanic, Richard Hilbe, Benedikt Schaefer, Sonja Wagner, Mirja Mittermaier, Felix Machleidt, Carmen Garcia, Christoph Ruwwe-Glösenkamp, Tilman Lingscheid, Laure Bosquillon de Jarcy, Miriam S. Stegemann, Moritz Pfeiffer, Linda Jürgens, Sophy Denker, Daniel Zickler, Philipp Enghard, Aleksej Zelezniak, Archie Campbell, Caroline Hayward, David J. Porteous, Riccardo E. Marioni, Alexander Uhrig, Holger Müller-Redetzky, Heinz Zoller, Judith Löffler-Ragg, Markus A. Keller, Ivan Tancevski, John F. Timms, Alexey Zaikin, Stefan Hippenstiel, Michael Ramharter, Martin Witzenrath, Norbert Suttorp, Kathryn Lilley, Michael Mülleder, Leif Erik Sander, PA-COVID-19 Study group, Markus Ralser, Florian Kurth

**Affiliations:** Charité – Universitätsmedizin Berlin, Department of Biochemistry, Berlin, Germany; The Francis Crick Institute, Molecular Biology of Metabolism Laboratory, London, UK; The University of Cambridge, Department of Biochemistry and Cambridge Centre for Proteomics, Cambridge, UK; Charité – Universitätsmedizin Berlin, Department of Infectious Diseases and Respiratory Medicine, Berlin, Germany; Charité – Universitätsmedizin Berlin, Medical Department of Hematology, Oncology & Tumor Immunology, Virchow Campus & Molekulares Krebsforschungszentrum, Berlin, Germany; Charité – Universitätsmedizin Berlin, Department of Nephrology and Internal Intensive Care Medicine, Berlin, Germany; Bernhard Nocht Institute for Tropical Medicine, Department of Tropical Medicine, and University Medical Center Hamburg-Eppendorf, Department of Medicine, Hamburg, Germany; University College London, Department of Mathematics, London, UK; National Phenome Centre and Imperial Clinical Phenotyping Centre, Department of Metabolism, Digestion and Reproduction, Imperial College London, London, UK; Lobachevsky University, Department of Applied Mathematics, Nizhny Novgorod, Russia; University College London, Department of Women’s Cancer, EGA Institute for Women’s Health, London, UK; University of Hertfordshire, School of Physics, Astronomy and Mathematics, Hatfield, UK; Sechenov First Moscow State Medical University, Department of Paediatrics and Paediatric Infections Diseases, Moscow, Russia; Lobachevsky University, Laboratory of Systems Medicine of Healthy Ageing, Nizhny Novgorod, Russia; Chalmers University of Technology, Department of Biology and Biological Engineering, Gothenburg, Sweden; University of Edinburgh, Centre for Genomic and Experimental Medicine, Institute of Genetics and Molecular Medicine, UK; University of Edinburgh, Usher Institute, Edinburgh, UK; University of Edinburgh, MRC Human Genetics Unit, Institute of Genetics and Molecular Medicine, Edinburgh, UK; Medical University of Innsbruck, Department of Internal Medicine II, Innsbruck, Austria; Medical University of Innsbruck, Christian Doppler Laboratory for Iron and Phosphate Biology, Department of Internal Medicine I, Innsbruck, Austria; Medical University of Innsbruck, Institute of Human Genetics, Innsbruck, Austria; Berlin Institute of Health, Berlin, Germany; Charité – Universitätsmedizin Berlin, Core Facility - High-Throughput Mass Spectrometry, Berlin, Germany; German Centre for Lung Research, Germany; Medical University Innsbruck, Division of Intensive Care and Emergency Medicine, Department of Internal Medicine, Innsbruck, Austria; Imperial College London, Section of Bioanalytical Chemistry, Division of Systems Medicine, Department of Metabolism, Digestion and Reproduction, London, UK

## Abstract

COVID-19 is highly variable in its clinical presentation, ranging from asymptomatic infection to severe organ damage and death. There is an urgent need for predictive markers that can guide clinical decision-making, inform about the effect of experimental therapies, and point to novel therapeutic targets. Here, we characterize the time-dependent progression of COVID-19 through different stages of the disease, by measuring 86 accredited diagnostic parameters and plasma proteomes at 687 sampling points, in a cohort of 139 patients during hospitalization. We report that the time-resolved patient molecular phenotypes reflect an initial spike in the systemic inflammatory response, which is gradually alleviated and followed by a protein signature indicative of tissue repair, metabolic reconstitution and immunomodulation. Further, we show that the early host response is predictive for the disease trajectory and gives rise to proteomic and diagnostic marker signatures that classify the need for supplemental oxygen therapy and mechanical ventilation, and that predict the time to recovery of mildly ill patients. In severely ill patients, the molecular phenotype of the early host response predicts survival, in two independent cohorts and weeks before outcome. We also identify age-specific molecular response to COVID-19, which involves increased inflammation and lipoprotein dysregulation in older patients. Our study provides a deep and time resolved molecular characterization of COVID-19 disease progression, and reports biomarkers for risk-adapted treatment strategies and molecular disease monitoring. Our study demonstrates accurate prognosis of COVID-19 outcome from proteomic signatures recorded weeks earlier.

## Introduction

COVID-19 has created unprecedented societal challenges, particularly for public health and the global economy^1–3^. Efficient management of these challenges is hampered by the variability of clinical manifestations, ranging from asymptomatic SARS-CoV-2 infection to death, despite maximal intensive care. Biomarkers and molecular signatures that enable accurate prognosis of future disease courses are needed to optimize resource allocation and to personalize treatment strategies. Patients likely to suffer progression to severe disease and organ failure and those likely to remain stable could be identified early, which is particularly valuable in scenarios where health care systems reach capacity limits. Prognostic panels would also optimize the monitoring of novel treatments, thereby accelerating clinical trials^4–6^. Knowledge of factors that differentiate recovery from deterioration over the course of the disease will enhance our understanding of the inflammatory host response as well as the underlying pathophysiology, and provide new therapeutic targets.

A number of biomarkers that classify COVID-19 severity have recently been described, based on clinical chemistry, enzyme activities, immunoprofiling, single cell sequencing, proteomics, and metabolomics^7–15^. These severity classifiers characterize COVID-19 pathology and host responses mainly in blood, serum, plasma or immune cells. Markers of dysregulated coagulation, inflammation, and organ dysfunction have also been established as risk factors for severe illness, including low platelet count, elevated levels of D-dimer, C-reactive protein (CRP), interleukin 6 (IL-6), ferritin, troponin, and markers of kidney injury^16,17^. By capturing host responses in an unbiased fashion, ‘omic’ investigations have the potential to go beyond classification of patients, as they capture disease-associated molecular signatures and put them into context with homeostatic pathways. Proteomic investigations have shown that the host response to COVID-19 is characterized by activation of the complement cascade and the acute phase response which center around IL-6-driven pathways. Other common antiviral pathways, such as type I interferons (IFN), do not dominate the overall response to COVID-19, probably reflecting evasion of the IFN system by SARS-CoV-2 and subsequent activation of inflammatory cascades^18,19^. Further, proteomic data and diagnostic parameters can directly point to underlying pathological mechanisms and possible therapeutic targets. For instance, using high-throughput proteomics, we reported a decline in plasma levels of gelsolin (GSN) in patients with severe COVID-19 in a previous study^7^, and recombinant human GSN is currently undergoing clinical testing for COVID-19 pneumonia in a phase II trial (ClinicalTrials.gov identifier: NCT04358406).

While many host response factors have been associated with COVID-19, it is still not understood how they change over time, how they depend on disease progression, and how they depend on one another or converge on common biological mechanisms. Also, while it is important to classify the severity of COVID-19 at the point of sampling, much higher clinical and biological value would be generated by markers that predict the outcome and future disease progression. Here, we performed a time-resolved deep clinical and molecular phenotyping of 139 adult patients with COVID-19 during hospitalization. At 687 sampling points, we measured a compendium of physiological parameters and routine diagnostic markers using gold standard, accredited clinical tests, and combined these measurements with untargeted high-throughput plasma proteomics^7^ to obtain a comprehensive picture of how the patients’ molecular phenotype develops over time. We study the associations between different markers and the dynamic changes in their levels to differentiate between distinct trajectories that characterize mild and severe COVID-19. We report that the inflammatory host response in the early disease stage is a major determinant of clinical progression, and facilitates accurate COVID-19 prognoses from the molecular patient phenotype of the early disease stage. We describe biomarker profiles and diagnostic parameters that classify treatment requirements, in particular the need for mechanical ventilation. Further, we report the future prediction of recovery time in mildly ill patients as well as the individual risk of worsening. In severely ill patients, proteomes that are recorded weeks before the clinical outcome accurately predict survival. Our study provides the scientific basis as well as a proof-of-principle for the predictability of COVID-19 disease trajectories based on the molecular phenotype of the early disease stage.

### Covariation of clinical diagnostic parameters and the plasma proteome characterizes the host response to COVID-19

Several recent investigations have identified protein biomarkers and clinical parameters that classify COVID-19 patients according to disease severity^7–15^. To identify *i*) which of these markers and parameters correlate with each other by being attributed to a common biological or physiological response, and *ii*) which markers predict outcome, we longitudinally phenotyped 139 individuals admitted at Charité University Hospital, Berlin, Germany, between March 01, 2020 and June 30, 2020 due to PCR confirmed SARS-CoV-2 infection (Supp. Fig. SF1). The patients exhibited highly variable disease courses, graded by the WHO ordinal scale for clinical improvement (Supp. Table 1), ranging from WHO grade 3, mild disease without need for oxygen support, to WHO grade 7, severe COVID-19 requiring invasive mechanical ventilation and additional organ support therapies^20^. Twenty-three patients were stable without the need of oxygen support (WHO grade 3) and could be discharged after a median of 7 days of inpatient care (Supp. Table 2, Supp. Fig. SF1). Forty seven (34%) patients required either low-flow or high-flow supplemental oxygen and sixty-nine (50%) patients either presented with severe COVID-19 (WHO grade 6 or 7, i.e. requiring mechanical ventilation) or deteriorated and required invasive mechanical ventilation during their hospitalization. Forty six (33%) required renal replacement therapy (RRT) and twenty two (16%) were treated with extracorporeal membrane oxygenation (ECMO). Twenty (13%) patients died, including one patient who died due to non-COVID-19 related cause. Patients with a severe course of disease were older than those with mild disease (49 years [IQR 35-70] for WHO grade 3 *vs* 62 years [IQR 53-73] for WHO grade 7, *P* = 0.02) and age of 65 years or older was associated with a higher risk of death (OR 4.1 [95% CI 1.5 - 11.5]). The median duration of hospitalization was 20 days (IQR 9 - 48) and correlated with severity (7 days for WHO grade 3 *vs* 51 days for WHO grade 7). The median time from admission to death was 26 days (IQR 16-43).

To capture the diverse disease trajectories on a molecular and biochemical level, we systematically collected 86 clinical and routine diagnostic parameters as measured with certified diagnostic tests, and monitored the development of risk scores such as the ‘sequential organ failure assessment’ (SOFA) score, blood gas analyses, blood cell counts, enzyme activities and inflammation biomarkers (Supp. Table 3). To complement these parameters with an untargeted analysis, we employed a recently developed high-throughput proteomics platform that bases on high flow-rate chromatography tandem mass spectrometry (LC-MS/MS) and acquired the data using SWATH-MS^7,21^. Creating a large mass spectrometry-based clinical proteomic dataset for a deeply characterized patient group, we measured a total of 687 human plasma proteomes, which required a total of 1169 non-blank LC-MS/MS injections, and quantified 309 plasma protein groups.

To identify interdependencies of the clinically established routine diagnostic parameters and the plasma proteomes, we characterized their covariation and present a direct correlation map (Fig. 1c, Supp. Fig. SF2, SF3), which comprehensively characterizes the host response to SARS-CoV-2 infection at a molecular level. To highlight some key observations, we report a robust positive or negative correlation of IL-6 levels and other inflammatory markers (CRP, procalcitonin) with acute phase proteins (APPs) (APOA2, APOE, CD14, CRP, GSN, ITIH3, ITIH4, LYZ, SAA1, SAA2, SERPINA1, SERPINA3, AHSG; protein names corresponding to the gene identifiers are provided in Supp. Table 3), coagulation factors and related proteins (FGA, FGB, FGG, F2, F12, KLKB1, PLG, SERPINC1), and the complement system (C1R, C1S, C8A, C9, CFB, CFD, CFHR5). Our data therefore links the prominent role of the IL-6 response in COVID-19^11^ to coagulation and the complement cascade. Consistently, in our data, markers of cardiac (troponin T, NT-proBNP) and renal (creatinine, urea) function as well as anaemia and dyserythropoiesis (hemoglobin, hematocrit, erythrocytes, and red blood cell distribution width) were correlated with various acute phase proteins (APPs) (APOA2, APOE, CD14, GSN, LYZ, SAA1, SAA2, SERPINA3, Fig. 1c), supporting the role of inflammation in COVID-19-related organ damage and its impact on erythropoiesis.

**Figure 1.**
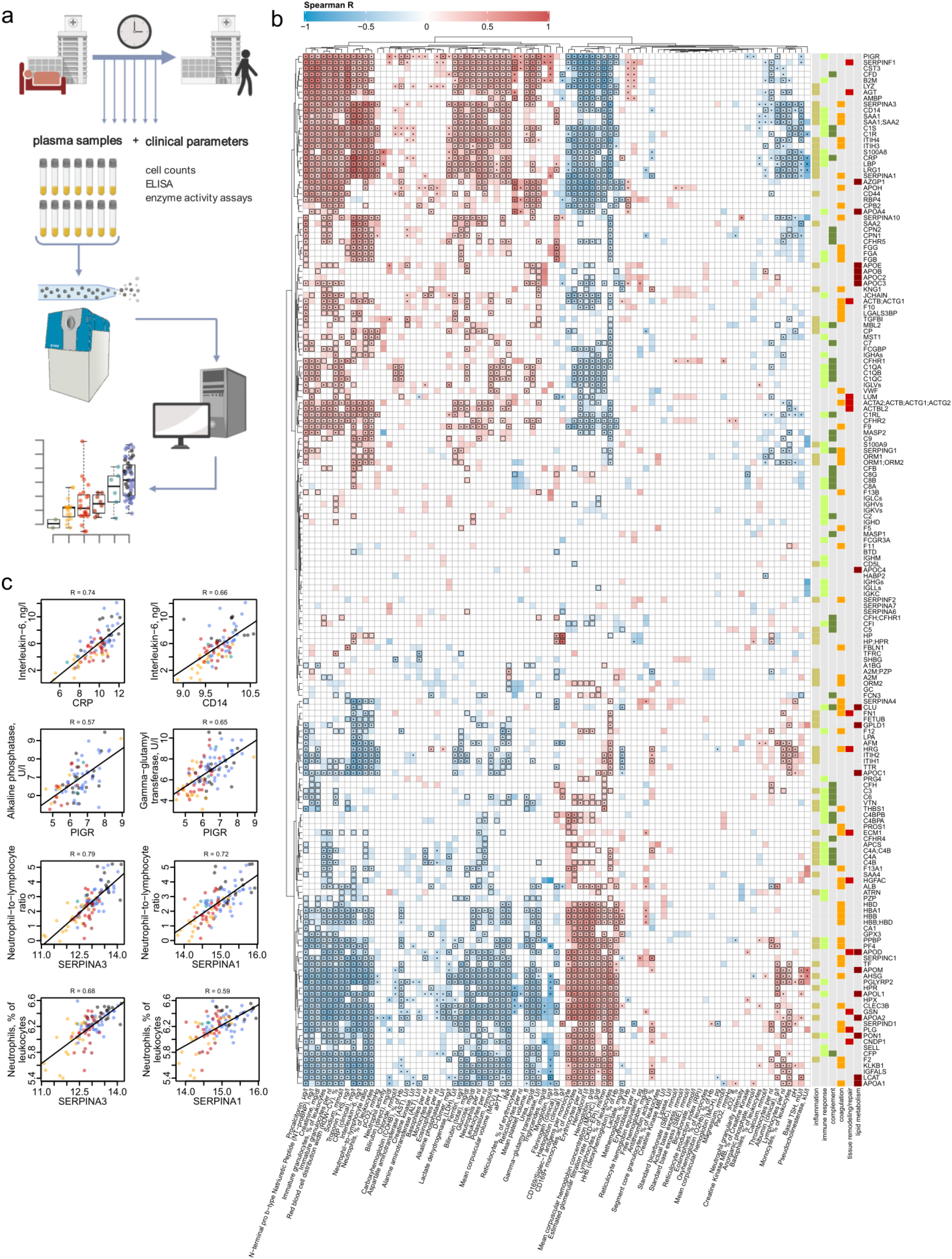
Interdependence of clinical, diagnostic, physiological and proteomic parameters in COVID-19. **a**. Study design. Schematic of the cohort of 139 patients with PCR confirmed SARS-CoV-2 infection treated at Charité University Hospital Berlin. Plasma proteomics and accredited diagnostic tests were applied at 687 sampling points, to generate high-resolution time series data for 86 routine diagnostic parameters and 309 protein quantities **b**. Covariation map for plasma proteins and routine diagnostic and physiological parameters (log2-transformed). Statistically significant correlations (Spearman; P < 0.05) are coloured. Dots indicate statistical significance after row-wise multiple-testing correction (FDR < 0.05), black rectangles – column-wise. The panel on the right of the heatmap provides manual functional annotation for the proteins. **c**. Covariation of key diagnostic parameters and plasma protein markers (log2-transformed) in COVID19.

Increased levels of neutrophils and occurrence of immature granulocyte precursors as markers of emergency myelopoiesis have been linked to severe COVID-19^12^. Our data reveals covariation between neutrophil counts and the levels of two inhibitors of neutrophil serine proteases, SERPINA1 and SERPINA3 (Fig. 1c). These two proteins show the highest correlation (0.72 and 0.79 Spearman R, respectively) with the neutrophil-to-lymphocyte ratio (NLR), a prognostic marker for COVID-19^22,23^. We further report a strong correlation (Fig. 1c) of alkaline phosphatase and gamma-glutamyl transferase activities, both characteristic of biliary disorders^24^, with plasma levels of the polymeric immunoglobulin receptor (PIGR). We note that cholangiocytes (bile duct epithelium cells) express ACE-2 and can be directly infected with SARS-CoV-2^25^, leading to host viral response-induced expression of PIGR and cell destruction^26^.

### A map of plasma proteins and clinical diagnostic parameters depending on age and disease severity

We identified 113 proteins and 55 diagnostic parameters in our cohort that increase or decrease according to the severity of COVID-19 (Fig. 2, Supp. Fig. SF4 and SF5; Methods). To our knowledge, more than 30 of these proteins have not been associated with COVID-19 severity previously (Supp. Table 3). These include mediators of inflammation and immune response (CD44, B2M, PIGR, A2M), components of the complement cascade (CFD, CFHRs), and several apolipoproteins (APOA2, APOC3, APOD, APOE and APOL1). Furthermore, numerous markers of organ dysfunction (cardiac: NT-proBNP, troponin T; renal: creatinine, urea; liver: aspartate aminotransferase, alanine aminotransferase, gamma-glutamyl transferase, total bilirubin) and, inversely, anemia (hemoglobin, erythrocytes, hematocrit) were correlated with disease severity. In order to further dissect the proteomic signatures of the most severely ill patients (WHO grade 7), we specifically characterized the impact of organ support treatments (RRT and ECMO) on the patients’ molecular phenotype (Supp. Fig. SF6 and SF7). We discuss these findings in Supp. Note 1.

**Figure 2.**
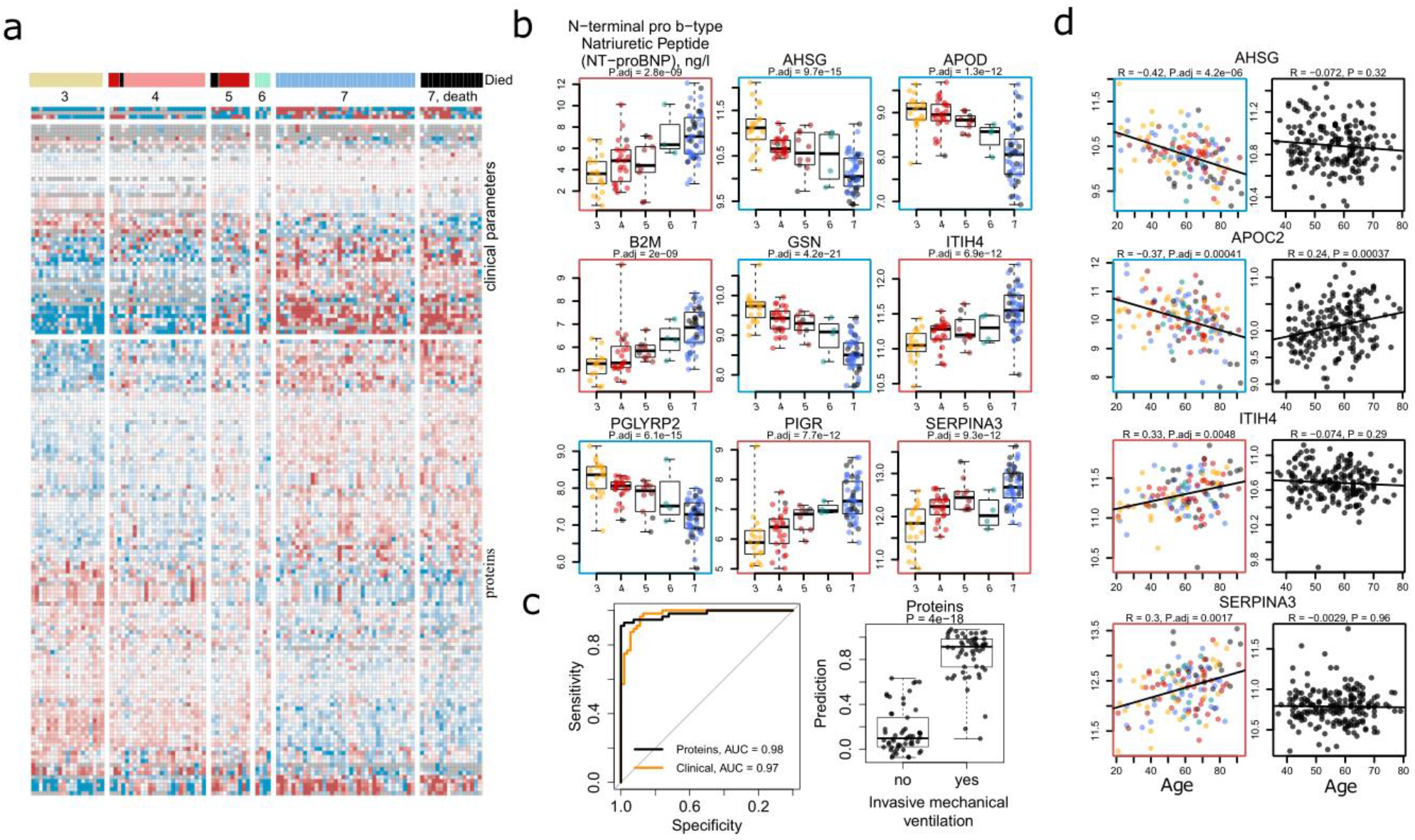
The molecular phenotype of patients with COVID-19 and its dependency on severity and age. **a**. Abundance of plasma proteomes and clinical-diagnostic parameters dependent on COVID-19 severity irrespective of age. Individual patients are grouped by severity. The severity of COVID-19 for each patient is represented by the maximal clinical treatment received (WHO ordinal scale, Supp. Table 1). 113 proteins and 55 routine diagnostic parameters vary significantly (FDR < 0.05) between patients of the different WHO groups upon accounting for age as a covariate using linear modelling^28^, Methods. A fully annotated heatmap is provided in Supp. Fig. SF4. **b**. Selected protein markers and routine diagnostic parameters (log2-transformed) plotted against the WHO ordinal scale indicating the disease severity of the patient. **c**. Performance of a machine learning model characterising the need for invasive mechanical ventilation, based on either the proteomic data or a signature assembled from accredited diagnostic parameters (Methods). Performance assessment of the classifier was carried out on the test samples, which were held out during training in leave-one-out fashion. **d**. Selected proteins differentially abundant depending on age (FDR < 0.05). Left, coloured: this data set (log2-transformed levels; statistical testing was performed by accounting for the WHO grade as a covariate^28^ (Methods); for visualisation only, the data were corrected for the WHO grade); right, black: general population (log2-transformed levels; Generation Scotland cohort).

A total of 62 proteins and 17 diagnostic parameters varied with patients’ age (Supp. Fig. SF8). Out of these, 37 proteins do not change with age in a pre-COVID-19 general population baseline (Generation Scotland cohort^27^), for which proteomes have been measured with the same proteomic technology^7^ (Supp. Fig. SF9). We note that markers that increase with age in COVID-19 patients are mostly driven by disease severity, which on average is higher in older patients (Supp. Table 2). To identify markers that are upregulated or downregulated in older patients in comparison to younger patients with comparable disease severity, we tested the relationship between omics feature levels and age by accounting for WHO severity grade as a covariate using linear modelling^28^ (Methods). This analysis identified 36 proteins and 12 clinical laboratory markers that show up-or are downregulation with age in COVID-19 patients (Fig. 2d, Supp. Fig. SF10, summarized in Fig. 5). Out of these, 20 proteins do not change with age in the pre-COVID-19 population baseline (Generation Scotland cohort proteome data^7^, Supp. Fig. SF9), or show the opposite correlation with age in the general population (e.g. APOC2, Fig. 2d). These proteins that only show an age-dependency in COVID-19 patients but not in the general population point to age-dependent differences in host response patterns to SARS-CoV-2, and include markers involved in inflammation (SERPINA3, ITIH4, SAA1, SAA1, SAA2, ITIH3, CFB, C7, AHSG), lipid metabolism (APOC1, APOC2, APOC3, APOB, APOD), and coagulation (KLKB1, FBLN1). We consider the implications of these findings in Supp. Note 2.

Using a machine learning algorithm based on gradient boosted decision trees (Methods), we then evaluated to what extent routine diagnostic parameters and proteins can accurately classify patients according to treatment requirements. Both proteomes and clinical diagnostic parameters were highly discriminative of the patient being on invasive mechanical ventilation (WHO grade 6 or 7, clinical laboratory values AUROC = 0.97, proteomic data AUROC = 0.98) (Fig. 2c). Scores reflecting the contribution of individual proteins and clinical parameters to the machine learning model are provided in Supp. Table 3.

### Time-dependent alleviation of severity indicators highlights the role of the early host response in COVID-19 progression

The time-resolved nature of our study facilitated a covariation analysis of protein levels and routine diagnostic parameters along the patient trajectory (Supp. Fig. SF11). Correlating the dynamics of omics features during the peak period of the disease (Methods), we noted covariation of inflammatory markers, acute phase proteins (APPs), fibrinogen precursor proteins, and the neutrophil-to-lymphocyte ratio. The correlation between APPs and the markers of cardiac and renal impairment, observed across different patients without taking into account any temporal resolution (Fig. 1b) was not reflected as a trend over time (Supp. Fig. SF12).

To further dissect the dynamics of omics markers during the course of COVID-19, we determined the longitudinal trend for all protein and routine diagnostic parameters during the peak period of the disease (i.e. while receiving maximal treatment; Methods). In total, 89 proteins and 37 clinical parameters significantly changed over time (Fig. 3c, trends across all time points at the maximum WHO grade are provided in Supp. Fig. SF13; Methods). As a general pattern, proteins and diagnostic parameters upregulated in severe COVID-19 were downregulated over time and vice versa. We further observed that signals of the markers that varied with COVID-19 severity were most prominently changed in the early samples (Supp. Fig. SF5), but alleviated with time (Supp. Fig. SF13; summarized in Fig. 5). For example, components of the coagulation cascade with known acute phase activity, such as fibrinogen and complement factors, were significantly downregulated over time during the peak period of the disease. Proteins indicative of inflammatory response (e.g. ORM1, SERPINA1 and SERPINA3, SAA1, SAA2^29–31^) and clinical markers of inflammation, like CRP or IL-6, also decreased over time. Conversely, extracellular matrix proteins, such as ECM1, LUM, immunoregulatory factors (e.g. A2M^32^, HRG^33^) and proteins involved in lipid metabolism (e.g. APOC1, APOD, APOM, GPLD1, PON1), and negative APPs (e.g. ITIH1, Fig. 3b), which are downregulated in severe COVID-19 (Supp. Fig. SF5, summarized in Fig. 5), showed an increase over time. This general alleviation of the initial molecular phenotype of COVID-19 was consistently detected in both mildly and severely ill patients. Indeed, only 13 proteins showed differences in trend depending on the disease severity (Supp. Fig. SF14). Of note, patients who died of COVID-19 exhibited different dynamics of certain omics markers over time, including for example an increase in several APPs (discussed in the next section and Supp. Note 3). We provide visualization of individual trajectories for all omics features measured between the first and the last time points sampled at the peak of the disease (Supp. Fig. SF15 - SF19).

**Figure 3.**
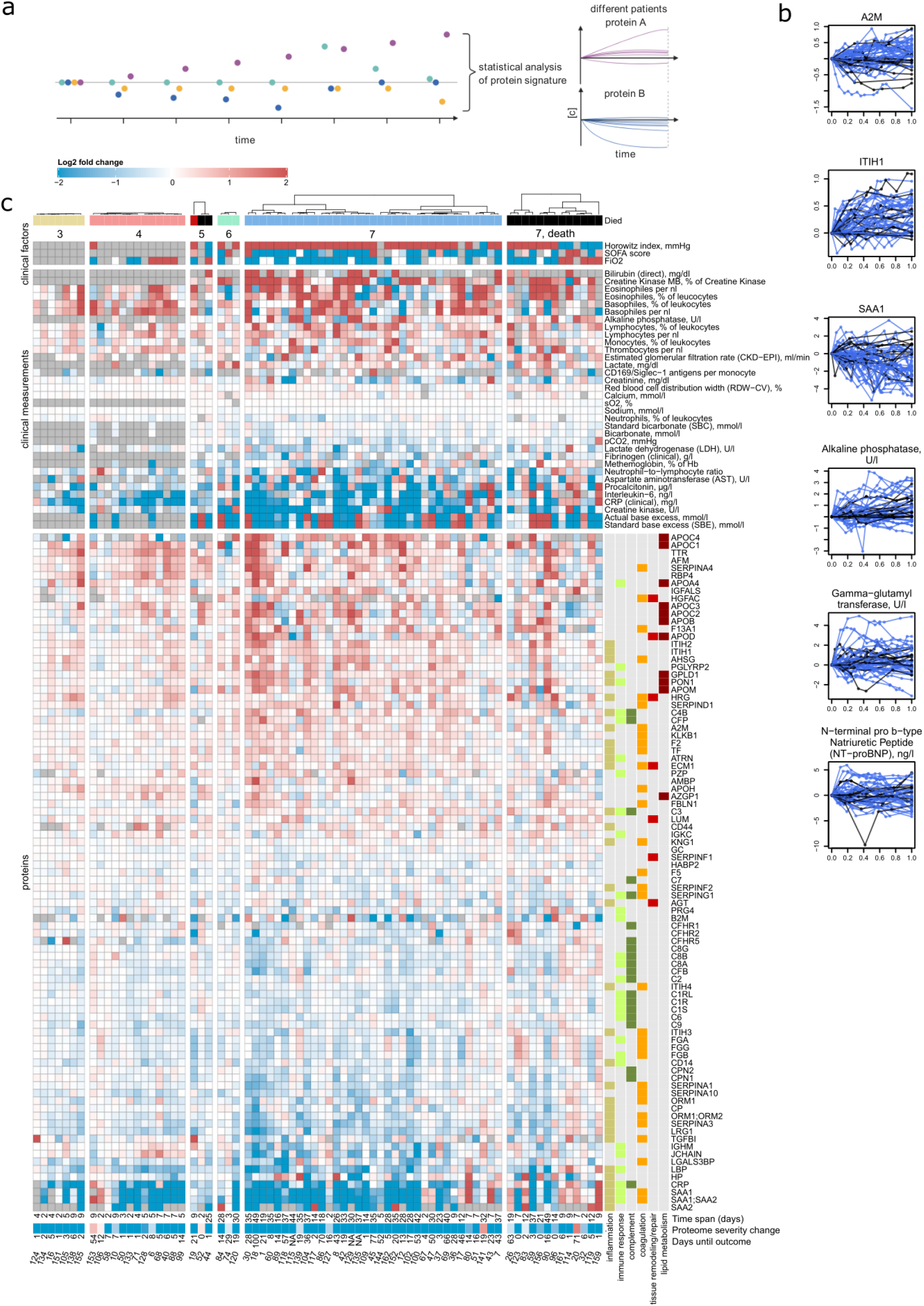
The progression of the COVID-19 molecular patient phenotype over time. **a**. Schematic: each patient is followed during inpatient care by repetitive sampling, and the “trajectory” of each of the proteins and the routine diagnostic features is analyzed (levels of different features are indicated with points of different colors at each time point). **b**. Trajectories (change of log2-transformed levels with time) for selected proteins and routine diagnostic parameters, for patients with maximum WHO grade 7. Sampling points during the peak period of the disease (Methods) are considered (blue – survivors, black – patients who died). X-axis: 0 – first time point measured at the peak of the disease, 1 – last. **c**. Protein levels and routine diagnostic parameters that change significantly (FDR < 0.05) over time during the peak of the disease, shown for individual patients stratified by their maximum treatment received (WHO grade): 89 proteins, 37 clinical laboratory measurements show time dependency during the disease course (illustrated as log2-fold changes). The panel on the right from the heatmap provides manual functional annotation for the proteins. Below the heatmap, the time span between the first and the last sampling time point at the peak of the disease is indicated, along with the change in the ‘proteome severity’ score computed with machine learning (for visualisation only; Methods) as well as the remaining time until the outcome (discharge or death).

Importantly, a number of organ function markers did not demonstrate an alleviation with time, particularly in severely ill patients. For example, NT-proBNP and gamma-glutamyl transferase activity remained persistently high on average, whereas the levels of alkaline phosphatase increased (Fig. 3b, Supp. Fig. SF13). However, there was significant heterogeneity in this response (see e.g. Fig. 3b, 3c), indicating a high diversity among the patients regarding indicators of organ damage.

Overall, the molecular patient phenotype reflects an initial spike in the systemic inflammatory response, which is gradually alleviated and followed by a protein signature indicative of tissue repair, metabolic reconstitution and immunomodulation. This was observed in both mildly and severely ill patients, highlighting the early disease phase as a major molecular determinant of the COVID-19 phenotype.

### Proteomes and diagnostic clinical markers predict future course of the disease and outcome

We hypothesized that the molecular signature of the initial host response can be exploited for prediction of the future disease course and outcome. We started by investigating the predictability of the remaining time in hospital for mildly ill patients with max WHO grade 3. We identified 26 protein biomarkers and 14 routine diagnostic markers (Fig. 4a, Supp. Fig. SF20) that correlate with the time between the first sampling point and release from inpatient care. The proteomic signature associated with longer duration of inpatient treatment and hence more severe COVID-19 progression includes upregulated components of the complement system (C1QA, C1QB, C1QC) and reflects altered coagulation (KLKB1, PLG, SERPIND1) and inflammation (CD14, B2M, SERPINA3, CRP, GPLD1, PGLYRP2, AHSG).

**Figure 4.**
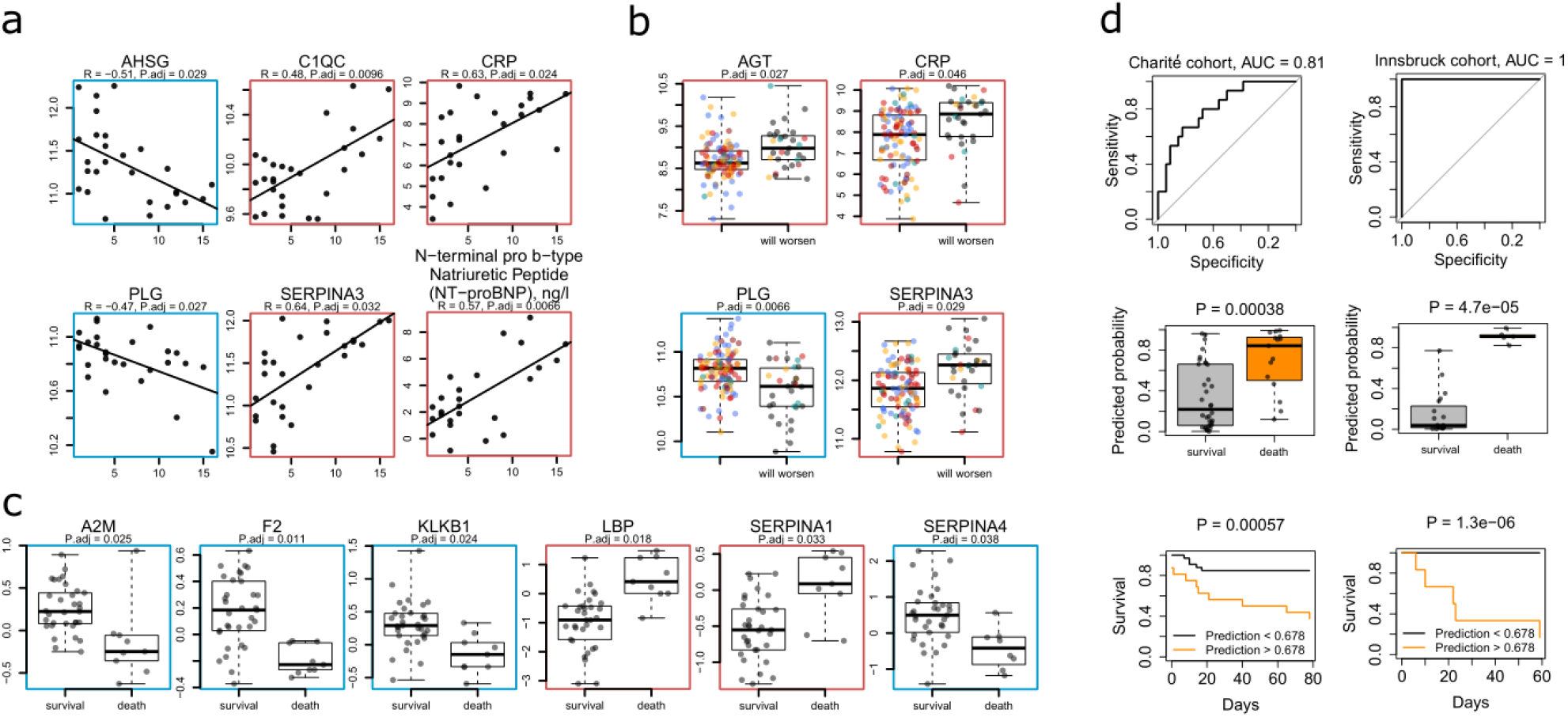
COVID-19 prognosis based on early molecular patient phenotype. **a**. Selected proteins and routine diagnostic parameters predictive (FDR < 0.05) of the remaining time in hospital for mildly ill (WHO grade 3) patients. Statistical testing was performed by including patient’s age as a covariate (Methods). Illustrated are the log2-transformed levels of the proteins (upon correction for age as a covariate, for visualisation only) at the first sampling point, plotted against the remaining time in hospital (days). **b**. Selected proteins that are predictive (FDR < 0.05) of the future clinical worsening of the disease (that is progression to a higher WHO grade in the future; Methods). Illustrated are the log2-transformed levels of the proteins at the first sampling point, upon correction (for visualisation only) for the impact of the WHO grade and age as a covariates^28^. **c**. Selected proteins (FDR < 0.05), for which time-dependent concentration changes (y-axis: log2 fold change) during the peak of the disease (Methods) are indicative of poor prognosis (death) in critically ill patients (maximum WHO grade 7). Patients who died but did not receive the full therapy according to their (presumed) wish (DNI, secondary DNR) were excluded from this analysis (Methods). **d**. Prediction of survival or death in critically ill patients, from the first sampling time point at critical treatment level (WHO grade 7) using a machine learning model based on parenclitic networks (Methods). Left panels: A classifier was trained on the proteomic data measured for the Charité cohort (WHO grade 7 only, survival n=34, death n=15, median time between sampling and outcome (i.e. discharge or death) 39 days, interquartile range 16 - 64 days). The performance was assessed on the test samples, which were held out during training (Methods). Right panels: A classifier trained on the Charité cohort was tested on the proteomic data measured for an independent cohort treated at the Medical University of Innsbruck (Austria) (WHO grade 6 and 7 patients, survival n=19, death n=5, median time between sampling and outcome 22 days, interquartile range 15 - 42 days). Bottom panels: Kaplan-Meier survival curves for Charité and Innsbruck cohorts using a threshold of predicted probability (0.678) chosen to maximize Youden’s J index (J = sensitivity + specificity - 1) of data from Charité cohort. Log-rank test was used to compare survival rates between patients with predicted death risk < 0.678 (black) and > 0.678 (orange).

**Figure 5.**
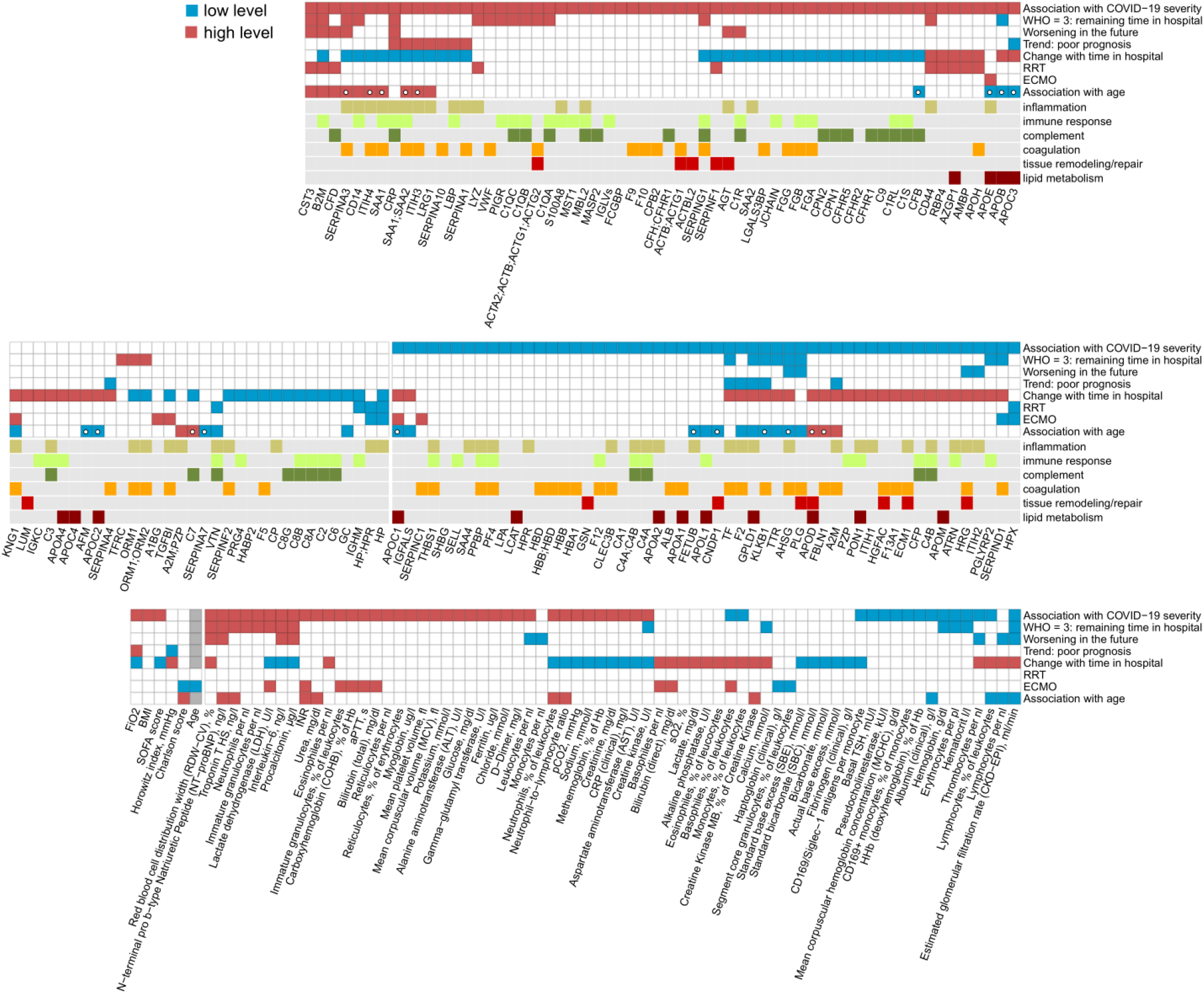
Summary: Association of individual plasma proteins, routine diagnostic and physiological parameters with treatment, progression and outcome of COVID-19. For each statistical test considered (association with COVID-19 severity, prediction of the remaining time in hospital for patients at WHO grade 3, prediction of worsening, i.e. progression to a higher WHO grade in the future, association of the trend during the peak period of the disease with the chance of death in patients with maximum WHO grade 7, trend during the peak period of the disease, association with renal replacement therapy (RRT), association with ECMO and association with higher patient age), measurements which show significant differences are highlighted, with the color indicating the trend, e.g. red for CST3 in the “Association with COVID-19 severity” test indicates higher levels of CST3 in severely ill patients. Proteins which change significantly with age in the Charité COVID-19 cohort (FDR < 0.05), but do not change significantly (P < 0.05) with age in the general population (Generation Scotland cohort), are highlighted with a white circle in the 8-th row (“Association with age”).

Next we explored the potential of using the levels of proteins and diagnostic parameters for prediction of future clinical worsening, defined as progression to a higher severity grade on the WHO scale, i.e. requirement for supplemental low-flow oxygen, high-flow oxygen or invasive mechanical ventilation (Fig. 4b, Supp. Fig. SF21; Box 1). Upon using a linear model to account for disease severity and age as covariates, 11 proteins and 9 clinical laboratory markers were identified as predictors of future worsening, across all severity grades (Methods). All 11 proteins are also among the markers of severity (Fig. 5, Supp. Fig. SF5), but increased or decreased plasma levels of these proteins preceded clinical deterioration. They reflect inflammation (CRP, ITIH2, SERPINA3, AHSG, B2M), coagulation (HRG, PLG), and complement activation (C1R, CFD). Further, higher levels of AGT and CST3 were also predictive of clinical deterioration.

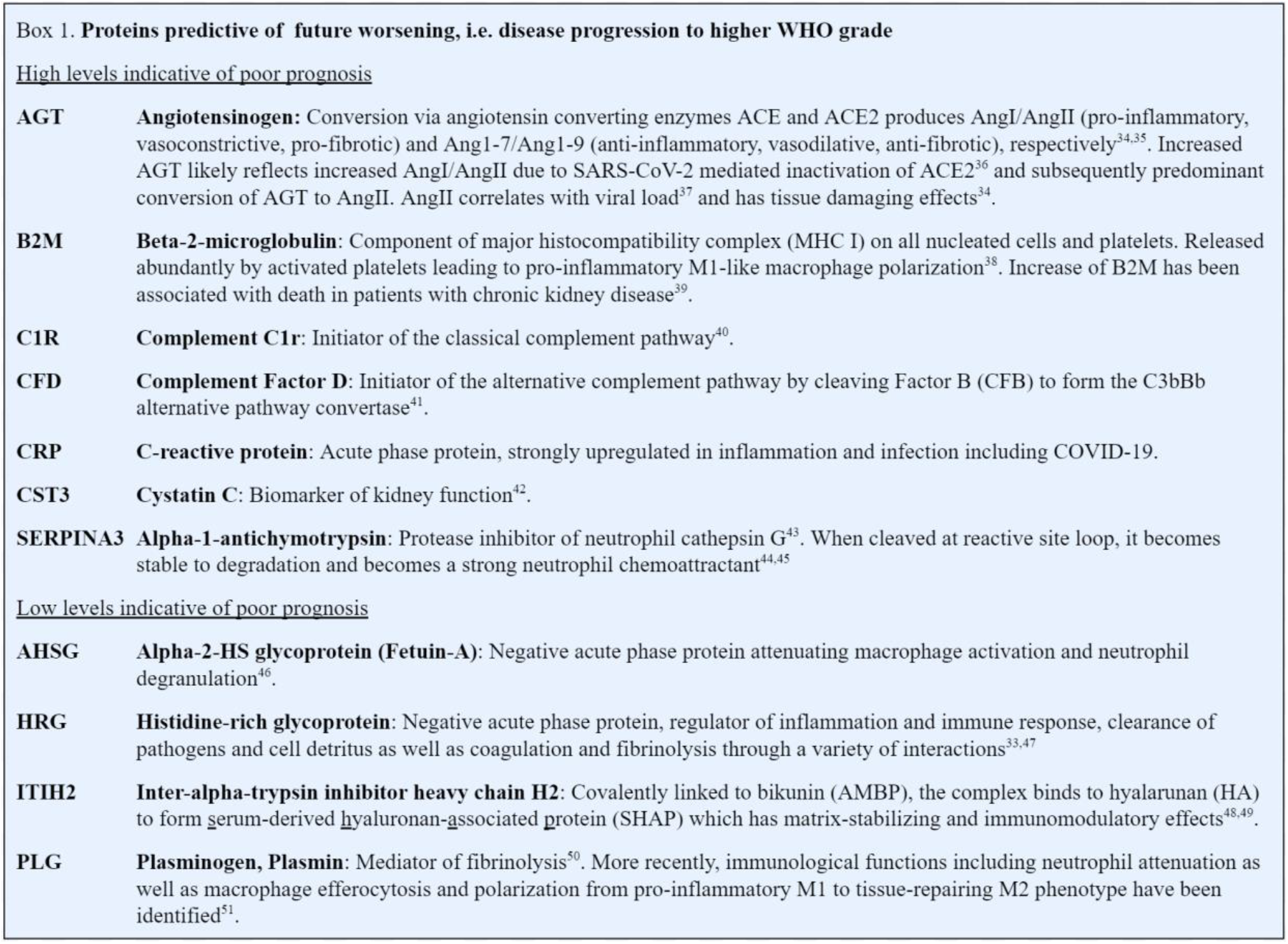

We further investigated whether the changes in the patients’ molecular phenotype over time were indicative of survival chances in critically ill patients (i.e. patients on invasive mechanical ventilation, with maximum WHO grade 7). Fourteen protein markers and 2 clinical measurements distinguished survivors from non-survivors (Fig. 4c, Supp. Fig. SF22). We observed a significant increase in inflammatory and acute phase proteins over time (e.g. SAA1;SAA2, CRP, ITIH3, LRG1, SERPINA1, and LBP) in patients with fatal outcome, indicating a persistent pro-inflammatory signature. Similarly, further decline of three key players in coagulation, F2, KLKB1 and the protease inhibitor A2M (all also generally decreased in severe COVID-19, Fig. 3c, Supp. Fig. SF5), was associated with adverse outcome.

In a last step, we established a machine learning model based on parenclitic networks^52,53^ (Methods) to predict survival in critically ill patients (WHO grade 7, n=49) from the proteome of the earliest single time point sample at WHO grade 7 for a particular patient. On the test subjects (the data of whom were excluded when training the machine learning model), high prediction accuracy was achieved, with AUC = 0.81 for the receiver-operating characteristic (ROC) curve. Of note, the model correctly predicted survival or adverse outcome weeks in advance (median time from prediction to outcome 39 days, interquartile range 16 - 64 days). To independently validate the machine learning model (trained on the ‘Charité’ cohort), we examined its performance on an independent cohort of 24 patients with COVID-19 from Austria (‘Innsbruck’ cohort, Methods). Despite considerable differences between the training and validation cohort, for instance regarding sampling time points, the machine learning model demonstrated high predictive power on the independent cohort (AUROC = 1.0, P = 0.00038, Fig. 4d). Using the cutoff value for survival prediction derived from the Charité cohort, the model correctly predicted the outcome for 18 out of 19 patients who survived and for 5 out of 5 patients who died in this independent cohort (Fig 4d).

## Discussion

Upfront clinical decision making is essential for optimal treatment allocation to patients as well as for efficient resource management within the hospital. For instance, early referral to intensive care treatment units has been shown to improve prognosis and outcome for patients with severe COVID-19^54^. One of the peculiarities of COVID-19 is that the examinable clinical conditions of patients often do not reflect the true severity of the disease, e.g. with respect to pulmonary insufficiency. In contrast to patients with severe bacterial pneumonia, patients with COVID-19 often clinically appear to be only slightly affected, despite being severely pulmonary insufficient, a phenomenon termed ‘happy’ hypoxemia^55^. Clinical decisions therefore need to be supported by objective, molecular diagnostics.

Previously, we and others investigated the association of plasma and serum biomarkers with COVID-19 severity^7–11,14^. In other studies, the potential prognostic value of several established and newly discovered blood markers for predicting the future course of disease was investigated, e.g. for IL-6, ferritin or resistin^56–58^. Yet, only very few studies set the prognostic value in relation to patient age and current disease severity, the two most important apparent characteristics for prognosis in COVID-19. For instance, a patient at WHO grade 5 (high-flow oxygen) is significantly more likely to progress to mechanical ventilation and subsequently die than a patient just admitted at WHO grade 3 (i.e. not requiring oxygen support). Likewise, a 90-year old patient at WHO grade 3 is significantly more likely to progress to more severe disease and to stay in the hospital longer than a 20-year old patient at the same WHO grade. Accounting for both WHO grade and age as covariates is thus relevant for establishing the prognostic value of biomarkers in COVID-19.

In this work we report proteomic and diagnostic markers that are measured with accredited clinical assays, that possess prognostic value for disease progression in COVID-19. These result from a comprehensive and time-resolved molecular phenotyping of a precisely characterized patient cohort which reflects the spread of varying COVID-19 trajectories (Supp. Note 4 for the resources generated). The measurements were performed in the early period of COVID-19. i.e. before dexamethasone became standard of care for patients with severe COVID-19^59^. Our data, which depict the initial spike in early inflammatory host response as determinant for the future course of the disease, is hence not confounded by standard anti-inflammatory treatment and will serve as particularly valuable resource and reference for future studies. As our results indicate, the patients in our cohort show proteomic signatures of higher basal inflammation with increasing age, which might be partially responsible for the higher risk of severe COVID-19 in older individuals. Whereas several approaches of targeted anti-inflammatory treatment have not been successful to prevent clinical deterioration in COVID-19 so far^60^, our study indicates that this special patient population might benefit particularly from treatments that mitigate the early inflammatory host response.

We use the time-resolved nature of our data to gain insights into the pathophysiology of COVID-19 disease progression. On the systems level, we observed an ‘alleviation’ of most key markers of disease severity over time (Fig. 3c, Fig. 5), in both mildly and severely ill patients, reflecting the gradual reversion of the patient’s molecular phenotype to baseline. It is followed by a protein signature indicative of tissue repair, metabolic reconstitution and immunomodulation. Exceptions are markers of organ dysfunction, especially markers of the cardiac system, which persist in the trajectory of severely ill patients.

Exploiting the early host molecular phenotype for the future projection of the disease course, our data reveals markers that correlate with the remaining time in hospital for mildly ill patients (at WHO grade 3), which we consider as an approximation of the time to recovery. Conversely we reveal markers that are prognostic for clinical worsening (i.e. that predict the future requirement of a greater degree of oxygen/organ support) for patients at all severity grades. Finally, we show that the molecular patient phenotype during the early disease stage distinguishes at high precision the survival chances among the most severely ill (WHO grade 7) patients. Training parenclitic neworks with proteomic measurements taken from samples that have large time intervals (median of 39 days) between collection and outcome (i.e. death or discharge), resulted in a machine learning model that can accurately predict outcome. Reassuringly, the model achieved even higher predictive power in an independent cohort of COVID-19 patients from another hospital.

While prognostic assessments using measurements at single time points might be useful for timely patient management and resource allocation, measuring dynamic parameters over time can be particularly valuable for treatment monitoring, including evaluation of experimental therapies as part of clinical trials. Moreover, the trends of certain proteins over time provide insights into the pathophysiology of COVID-19 and reveal potential targets for therapeutic interventions. As an example, we observed that a decline in SERPINA4 (kallistatin) levels over time in patients with severe disease was associated with an increased risk of death (Fig 4c). Kallistatin is known to possess protective properties in acute lung injury and should be considered as potential candidate for clinical testing in severe COVID-19, according to our data^61^.

We observed a considerable overlap between prognostic markers and those that are classifiers of COVID-19 severity (Fig. 5). Indeed, out of 61 prognostic markers, 50 are also classifiers of disease severity and displayed the same trend. As an example, SERPINA1 (Alpha-1 antitrypsin) and SERPINA3 (Alpha-1 antichymotrypsin) can be used for both classification of current severity and prediction of future disease course. Both proteins possess anti-inflammatory properties and are involved in protection of tissues from neutrophil elastase- and cathepsin G-mediated tissue damage^43^. Our data show strong correlation of both serpins with levels of neutrophils and neutrophil-to-lymphocyte ratio (NLR) in peripheral blood. SERPINA1 is mainly produced by the liver but also in epithelial cells, pulmonary alveolar cells, tissue macrophages, blood monocytes and granulocytes, so this finding could reflect a global response to the increased NLR. Yet, after binding to effector enzymes, SERPIN-proteinase complexes are normally rapidly cleared from the blood, but become resistant to degradation when cleaved at the reactive site loop^62^. Cleaved SERPINA1 and SERPINA3 have been shown to act as strong neutrophil chemoattractants^44,45^. The observed increase in levels of SERPINA1 and SERPINA3 might therefore partly reflect the more stable, chemoattractant, pro-inflammatory cleaved forms, rather than the short-lived tissue protective proteins in severe COVID-19. Given the prominent role of neutrophil-activation in severe COVID-19^12^, this finding merits further investigation.

Our data reveal AGT as another important marker predicting clinical worsening and indicating severe disease. Activation of angiotensinogen occurs via the protease renin and the endogenous angiotensin converting enzymes ACE or ACE2. ACE converts angiotensin I (AngI) into pro-inflammatory, vasoconstrictive, and pro-fibrotic angiotensin II (AngII)^34^. ACE2, in contrast, mediates conversion of angiotensins I and II into anti-inflammatory, vasodilative, anti-fibrotic and anti-oxidant angiotensins 1-9 (Ang1-9) and 1-7 (Ang1-7)^35^. SARS-CoV-2 invades host cells of the lung, heart, kidneys and other organs via ACE2, resulting in internalization and downregulation of ACE2^34,36,63^. Subsequently, angiotensinogen is converted predominantly via ACE to AngII and is less degraded by ACE2, resulting in AngII accumulation^64,65^. We can thus assume that the higher plasma levels of AGT gene products in severely ill patients, as measured in our study, mainly reflect the higher levels of AngII. Importantly, we observed a strong correlation of AGT with markers of acute kidney injury (AKI; creatinine, urea; Supp. Fig. SF2 and SF12), a frequent complication of COVID-19 and a risk factor for poor prognosis and fatal outcome^66^. Aggravated by the absence of tissue protective Ang1-7, elevated levels of AngII lead to activation of the renin-angiotensin-system (RAS) and contribute to hypoxic kidney injury^67^. Of note, apart from tissue damaging effects, AngII has been shown to linearly correlate with viral load and lung injury in SARS-CoV-2 infection^37^.

We observed that many of the markers that are both classifiers and predictors of the future disease course are initiators of the inflammatory response. This group includes some of the key initiators of the complement cascade, C1QA, C1QB, C1QC, C1R, CFD. In contrast, severity markers without prognostic value largely include downstream effectors of inflammatory-associated damage, such as GSN and the circulating actins ACTBL2 and ACTB, ACTG1, and ECM1.

We note that a number of markers identified as predictors are already approved as biomarkers by FDA, and for many of the proteins standardized quantification assays by ELISA or Selective Reaction Monitoring (SRMs) are available^68^. Hence, although the high-resolution high-throughput mass spectrometry platform as used in our study^7^ may be too complex for the use in routine laboratories, the panels can be put into practice via the routine assays. Future studies need to address the robustness of the predictors when measured as part of the clinical routine. The use of machine learning is currently not a certified method to inform clinical decisions. However, in addition to multiple works which have successfully used machine learning for clinical prognosis previously (see recent reviews^69–73^), our results bear a strong implication of the future potential of machine learning for clinical applications, including personalized medicine. This calls for a worldwide effort aimed at developing procedures which would allow reliable clinical validation of machine learning predictors, their approval and their routine deployment in the clinic.

In summary, by following a deeply phenotyped COVID-19 patient cohort over time at the level of the proteome and established diagnostic biomarkers and physiological parameters, we have created a rich data resource for understanding the progression of COVID-19 until the outcome. We have shown that an early spike in the inflammatory response is a key determinant of progression of COVID-19, and that future disease progression is predictable by using panels of routine diagnostic parameters as well as proteomic measurements from early time point samples. Indeed, we achieved an accurate prediction of survival of severely ill COVID-19 patients from plasma proteome samples that were, on average, recorded 39 days prior to the outcome. Our study provides comprehensive information about the key determinants of the varying COVID-19 disease trajectories as well as marker panels for early prognosis that can be exploited for clinical decision making, to devise personalized therapies, as well as for monitoring the development of much needed COVID-19 treatments.

## Data Availability

The datasets generated during and/or analysed during the current study are available from the corresponding author on reasonable request

## Supplementary Note 1. Diagnostic parameters and Proteome signatures that indicate therapeutic interventions

We investigated to what extent specific therapies, such as organ support in severely ill patients (renal replacement therapy (RRT), extracorporeal membrane oxygenation (ECMO)) were reflected in the proteome and at the level of routine diagnostic parameters. HP and HPX were reduced in patients on RRT and ECMO, reflecting hemolysis in the extracorporeal circuits (Supp. Fig. SF6 and SF7). Elevated SERPINC1 levels mirror substitution of antithrombin during ECMO. The reason for elevated levels of APOE in patients with ECMO is unclear, but is in line with reports on increased levels of APOE in pediatric patients after cardiopulmonary bypass^74^. The proteins increased in patients receiving RRT mainly reflect impaired kidney function and have been associated with RRT before (AMBP, B2M, CST3, LYZ, RBP4, Supp. Fig. SF6)^75^. Of note, increased levels of AMBP, B2M and LYZ have been associated with death in chronic kidney disease^39^. Levels of CFD and APOH, both involved in the complement system, were also increased^41,76^. CFD is eliminated renally and accumulates in end stage renal disease, possibly leading to enhanced complement activation via the alternative pathway^77^. In contrast, levels of APOH have even been reported to be slightly lower following high-flux hemodialysis^78^.

We note that the analysis of the effect of treatments on the proteome has two limitations. First, some of the markers identified might be prognostic for the treatment rather than reflect its effect. Age and the Charlson comorbidity index belong to this category: patients receiving ECMO were significantly younger and had a lower number of pre-existing chronic conditions than those who did not. Second, the results might be partially confounded by the time elapsed from the onset of the disease, as we have shown (Fig. 3) that omics signature does change with time in COVID-19 patients while on invasive mechanical ventilation.

## Supplementary Note 2. Age-specific response to COVID-19 in the context of severity markers

Older age is one of the most significant risk factors for severe disease and adverse outcome in COVID-19. Enhanced understanding of underlying mechanisms for the age-specific response to SARS-CoV-2 infection is therefore important and needed for the development of effective age-specific strategies for prevention and treatment. Furthermore, dissecting the age-specific components of the host response will improve our knowledge of the pathogenicity of similar viruses, making the world better prepared for future pandemics. Current theories characterizing the link between the higher age and risk for severe disease include immunosenescence, elevated baseline inflammation, or altered protein glycosylation landscape leading to impaired antiviral response or reduced immune tolerance^36,79–81^. However, a detailed and mechanistic understanding of the relation between COVID-19 and aging is lacking. In this work, we leverage the large size and high precision of the proteomic data acquired to map the age-related response to COVID-19, to provide a reference dataset (Fig. 2d, Fig. 5, Supp. Fig. SF10) for future studies addressing this problem.

We report elevation of several inflammatory and acute phase proteins such as SERPINA3, ITIH4, SAA1, and ITIH3 in older patients with COVID-19. SAA1 has been shown to induce macrophage polarization to the M2-type which promotes tissue repair but also possesses pro-fibrotic properties involved in the pathogenesis of pulmonary fibrosis^82–84^. Moreover, SAA1 mediates displacement of APOA1 from HDL leading to loss of the cardio- and vasoprotective properties of HDL^85^. SERPINA3, as discussed above, has an ambivalent role as a neutrophil proteinase inhibitor but also a powerful neutrophil chemoattractant. Upregulation of SERPINA3 with age in COVID-19, along with the higher neutrophil-to-lymphocyte ratio, suggests that excessive neutrophil response is one of the aggravating factors in older COVID-19 patients. Taken together, our findings point towards a disproportionately dysregulated inflammatory response to SARS-CoV-2 with age, which may be explained by an increased baseline inflammation and immunosenescence in older patients^86–88^. Age-dependent increase of FBLN1 and decrease of KLKB1 reflect alterations in blood coagulation which may aggravate this effect by predisposing older patients to thromboembolic events, one of the key clinical characteristics of severe COVID-19.

Interestingly, a number of apolipoproteins displayed a strong age-specific signature in COVID-19. For instance, APOC2, a component of chylomicrons, very low density lipoprotein (VLDL) and high density lipoprotein (HDL), and activator of lipoprotein lipase involved in triglyceride metabolism^89^, was downregulated with age in COVID-19, but upregulated with age in the general population^90,91^ (Fig. 2d). Dysregulation of apolipoproteins has been observed in community acquired pneumonia and associated with unfavourable outcome^92^. Remarkably, contrary to the general trend, APOD, APOC3 and APOE show opposite trends in older COVID-19 patients and in severe disease (Fig. 5). APOD is expressed by many tissues, including the brain^93^. An increase in APOD has been previously observed in ischemic stroke and CNS inflammation and may reflect (subclinical) involvement of the central nervous system especially in older patients with more severe inflammation and more comorbidities^94^. Conversely, high levels of APOD have been shown to temper coronavirus-mediated encephalitis in mice, indicating its role as a marker of CNS damage as well as tissue protection and repair^95^. APOE, involved in inflammation, immune response and lipid metabolism, is upregulated in severe COVID-19 but downregulated with age in this cohort. APOE typically mediates anti-inflammatory effects by downregulation of NFκB and inhibition of macrophage response to IFNy and TLR3, both mediators of viral immune response. Moreover, it neutralizes bacterial LPS and enhances the adaptive immune response by facilitating antigen presentation^96^. Downregulation with age may reflect a compromised immune response leading to over-activation of NFκB and insufficient pathogen clearance in older patients. Finally, APOE has been described to reduce proliferation of myeloid progenitor cells^97^ and to reduce myeloid derived suppressor cell (MDSC) survival in mice^98^. Thus, lower levels of APOE in the elderly may favor expansion of immature and dysfunctional neutrophils that have been described as a hallmark in severe COVID-19^12^. This broad involvement of APOE merits further investigation in future studies.

## Supplementary Note 3. Diverging trends at the proteome level during the disease peak in individual patients

Some patients (59, 90, 96, 123) who died exhibited protein concentration trajectories distinctly similar to “typical” survivors (Fig. 3c). Two of them (59, 90) had a prolonged ICU stay with repeated septic episodes and finally defined limitations of therapy according to presumed patients’ wishes (“secondary DNR”). Their protein signatures probably reflect the phenomenon of immune paralysis that can follow bacterial sepsis associated with a prolonged ICU treatment^99^. One patient (96) was receiving ongoing immunosuppressive therapy for an autoimmune disorder, and a fourth patient (123) had a history of kidney transplantation, both died of septic shock. Whether the particular group of solid organ recipients shows a distinct protein signature associated with the outcome requires further investigation.

We also note that some survivors do not show a trajectory characteristic of the typical ‘alleviation’ of the proteomic phenotype (WHO = 4: 58, 106, 153; WHO = 6 or 7: 43, 80). Specifically, the proteomic response in patients 106, 153 and 141 was indicative of overall ‘worsening’ of the proteome (Fig. 3c). In contrast, patients 43 and 80 exhibited the overall ‘alleviation’ of the proteome, except for the spike in the levels of CRP and serum amyloid (Fig. 3c). Shorter time spans between sampling days may explain these observations in four of these patients (43, 58, 80, 106), indicating that the host inflammatory response requires a certain time to resolve, especially in more severely ill patients, and some of the markers of systemic inflammation might linger, whereas a typical alleviation of the proteomic signature can be observed even within a few days in moderate disease courses. The unusual pattern of patient 153 was likely confounded by a skin infection that subsequently required antibiotic treatment.

## Supplementary Note 4: Overview of Resources generated

We provide deep and time-resolved resources that characterize COVID-19 at the level of plasma proteomes and established diagnostic parameters (summarized in Box 2). We demonstrate the extent to which proteomes and diagnostic parameters interdepend, in initial response to the disease and in dynamics during the disease course. We show how they change with age, differ depending on the disease severity, reflect the therapy received and evolve over time. Our data have been acquired for COVID-19 patients’ samples and analysed in the context of general population proteomics (Generation Scotland) for which samples have been measured with the same proteomic technology^7^, but we also expect it to be of high value as a reference for studies of other types of viral pneumonia as well as any investigations involving both routine clinical phenotyping and plasma proteomics.

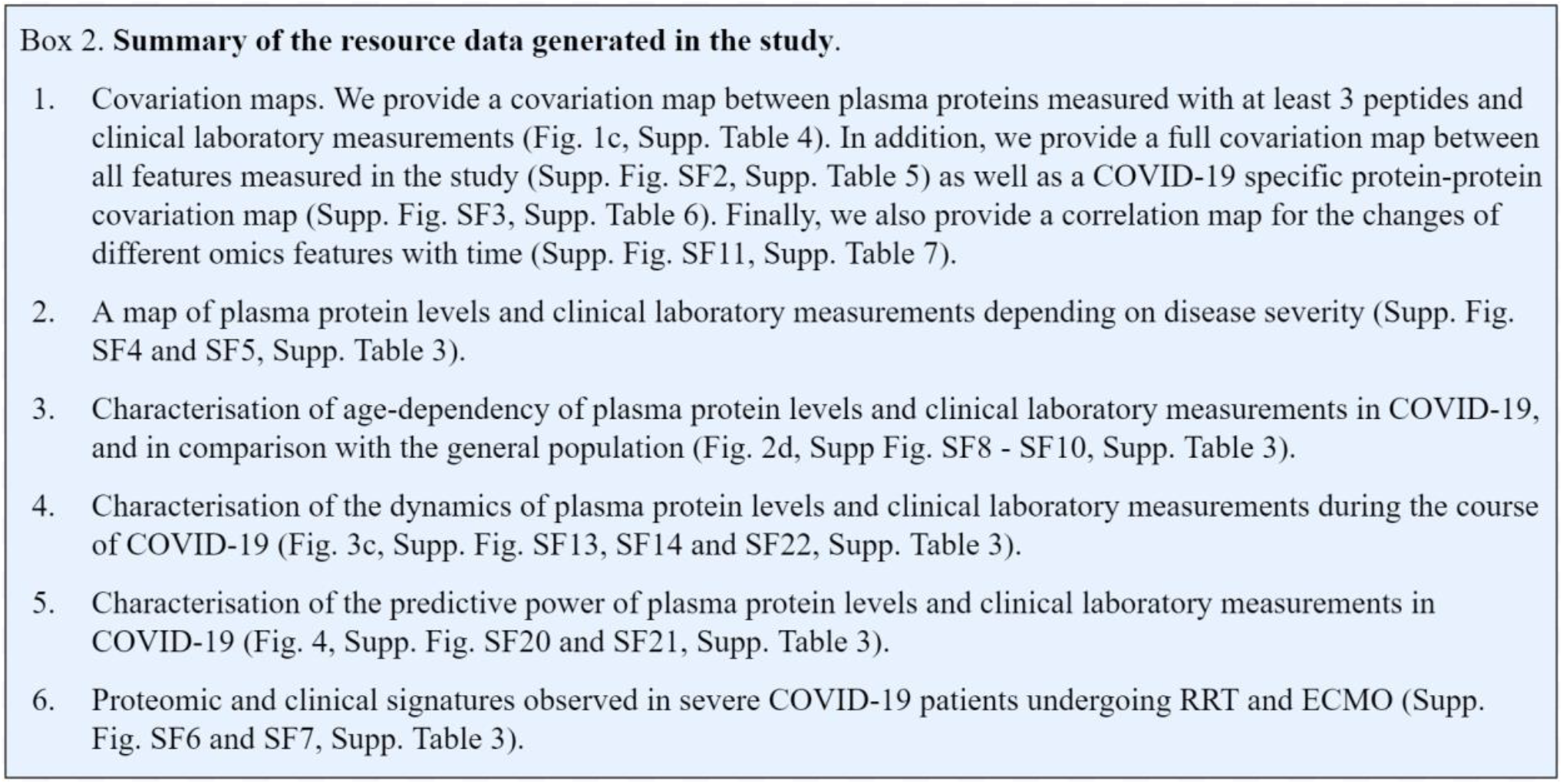

## Acknowledgements

We thank Jan-David Manntz (Beckman, Germany) for help with the Biomek i7, Robert Lane, Jean-Baptiste Vincedent and Nick Morrice (SCIEX) for help with the TripleTOF 6600. This work was supported by the Berlin University Alliance (501_Massenspektrometrie, 501_Linklab, 112_PreEP_Corona_Ralser), by UKRI/NIHR through the UK Coronavirus Immunology Consortium (UK-CIC), the BMBF/DLR Projektträger (01KI20160A, 01ZX1604B, 01KI20337, 01KX2021), Charité-BIH Centrum für Therapieforschung (BIH_PA_covid-19_Ralser), the BBSRC (BB/N015215/1, BB/N015282/1), the Francis Crick Institute, which receives its core funding from Cancer Research UK (FC001134), the UK Medical Research Council (FC001134), and the Wellcome Trust (FC001134 and IA 200829/Z/16/Z). The work was further supported by the Ministry of Education and Research (BMBF), as part of the National Research Node ‘Mass spectrometry in Systems Medicine (MSCoresys), under grant agreement 031L0220A. Leif Erik Sander is supported by the German Research Foundation (DFG, SFB-TR84 114933180) and by the Berlin Institute of Health (BIH), which receives funding from the Ministry of Education and Research (BMBF). Martin Witzenrath is supported by grants from the German Research Foundation, SFB-TR84 C06 and C09, by the German Ministry of Education and Research (BMBF) in the framework of the CAPSyS (01ZX1304B), CAPSyS-COVID (01ZX1604B), SYMPATH (01ZX1906A) and PROVID project (01KI20160A) and by the Berlin Institute of Health (CM-COVID). Stefan Hippenstiel is supported by the German Research Foundation (DFG, SFB-TR84 A04 and B06), and the BMBF (PROVID, and project 01KI2082). Norbert Suttorp is supported by grants from the German Research Foundation, SFB-TR84 C09 und Z02, by the German Ministry of Education and Research (BMBF) in the framework of the PROGRESS 01KI07114. The study was further supported by Wellcome Trust (200829/Z/16/Z). The Generation Scotland study received core support from the Chief Scientist Office of the Scottish Government Health Directorates (CZD/16/6) and the Scottish Funding Council (HR03006), and is now supported by the Wellcome Trust (216767/Z/19/Z). Archie Campbell is funded by HDR UK and the Wellcome Trust (216767/Z/19/Z). Caroline Hayward is supported by an MRC University Unit Programme Grant (MC_UU_00007/10) (QTL in Health and Disease). Riccardo Marioni is supported by an Alzheimer’s Research UK project grant (ARUK-PG2017B-10). H. Whitwell, JF. Timms, A. Zaikin and T. Nazarenko are supported by a Medical Research Council grant (MR/R02524X/1) and H. Whitwell, A. Zaikin and O. Blyuss by the Ministry of Science and Higher Education agreement No. 075-15-2020-808. H. Whitwell is supported by the National Institute for Health Research (NIHR) Imperial Biomedical Research Centre (BRC). J. Timms is supported by the National Institute for Health Research (NIHR) UCLH/UCL Biomedical Research Centre. Mirja Mittermaier is a participant in the BIH-Charité Digital Clinician Scientist Program funded by the Charité – Universitätsmedizin Berlin, the Berlin Institute of Health, and the German Research Foundation (DFG). Markus A. Keller is supported by the Austrian Science Funds (FWF; P33333) and the Austrian Research Promotion Agency (FFG, #878654). Figures were created with biorender.com.

## Methods

### Charité patient cohort and clinical data

Patients were recruited within the Pa-COVID-19 study conducted at Charité - Universitätsmedizin Berlin, a prospective observational cohort study on the pathophysiology of COVID-19. The study protocol has been described in detail before^100^. All patients with PCR-confirmed SARS-CoV-2 infection were eligible for inclusion. Refusal to provide informed consent by the patient or a legal representative and any condition prohibiting supplemental blood for serial biosampling sampling were exclusion criteria. Patients were treated according to current national and international guidelines. Three patients had *Do Not Intubate and Do Not Resuscitate* (DNI/DNR) orders in place, declining mechanical ventilation and other organ support or cardiopulmonary resuscitation. In 4 further cases, limitation of therapy was decided at a later time point according to the patient’s presumed wish (“secondary DNR”) and predictably unfavorable outcome. All other patients received maximum intensive care treatment including organ replacement therapies at the discretion of the responsible physicians.

Biosampling for proteome measurement was performed 3 times per week after inclusion. The WHO ordinal scale for clinical improvement (Table 1) was used to assess disease severity. ARDS was defined according to the Berlin ARDS criteria^101^. Sepsis was defined according to sepsis-3 criteria^102^. The study was approved by the ethics committee of Charité - Universitätsmedizin Berlin (EA2/066/20) and conducted in accordance with the Declaration of Helsinki and guidelines of Good Clinical Practice (ICH 1996). The study is registered in the German and the WHO international registry for clinical studies (DRKS00021688). Clinical data was captured in a purpose built electronic case report form data using the capture system SecuTrial®. All routine laboratory parameters were analyzed in accredited laboratories at Charité - Universitätsmedizin Berlin. Pseudonymized data exported from SecuTrial® were processed using JMP Pro 14 (SAS Institute Inc., Cary, NC, USA). If a laboratory value was missing for a given day, values from up to two preceding days were used for the analysis.

### Innsbruck Patient cohort and clinical data

Serum samples from patients admitted to the intensive care unit at the Department of Medicine, University Hospital of Innsbruck for the treatment of respiratory failure due to severe COVID-19 were collected within the first days (median 7.5, IQR 5-12) after admission. Written informed consent was either obtained before sampling or retrospectively after recovery, if patients were mechanically ventilated at the time of sampling. COVID-19 was diagnosed on the basis of a (i) positive SARS-CoV2 PCR within the last 7 days prior to study inclusion (ii) respiratory failure defined as a partial pressure of oxygen < 60 mmHg on arterial blood gas analysis or a peripheral oxygen saturation of < 90% and (iii) typical infiltrates on computed tomography scanning of the chest. Patients were treated according to national guidelines. The study was approved by the local ethics research committee EK-Nr. 1107/2020, and EK-Nr. 1103/2020 for follow-up.

### Mass spectrometry and data analysis

#### Materials

Water (LC-MS Grade, Optima; 10509404), Acetonitrile (LC-MS Grade, Optima; 10001334) and Methanol (LC-MS Grade, Optima, A456-212) were purchased from Fisher Chemicals. DL-Dithiothreitol (BioUltra, 43815), Iodoacetamide (BioUltra, I1149) and Ammonium Bicarbonate (Eluent additive for LC-MS, 40867) were purchased from Sigma Aldrich. Urea (puriss. P.a., reag. Ph. Eur., 33247H) and Acetic Acid (Eluent additive for LC-MS, 49199) were purchased from Honeywell Research Chemicals. Trypsin (Sequence grade, V511X) was purchased from Promega. Control samples were prepared from Human Serum (Sigma Aldrich, S7023-50MB) and Human Plasma (EDTA, Pooled Donor, Genetex GTX73265).

Mass spectrometry-based proteomics analysis was performed as described previously^7^ with minor adjustments to the workflow. Semi-automated sample preparation was performed in 96-well format, using previously prepared stock solution plates stored at -80°C. Briefly, 5μl of thawed plasma samples were transferred to the pre-made denaturation/reduction stock solution plates (55μl 8M Urea, 100mM ammonium bicarbonate (ABC), 50mM dithiothreitol). Subsequently, the plates were centrifuged for 15s at pulse setting (Eppendorf Centrifuge 5810R), mixed and incubated at 30°C for 60 minutes. 5μl was then transferred from the iodoacetamide stock solution plate (100mM) to the sample plate and incubated in the dark at 23°C for 30 minutes before dilution with 100mM ABC buffer (340μl). 220μl of this solution was transferred to the pre-made trypsin stock solution plate (12.5μl, 0.1μg/μl) and incubated at 37°C for 17 h (Memmert IPP55 incubator). The trypsin/total protein ratio was ∼1/40. The digestion was quenched by addition of formic acid (10% v/v, 25μl). The digestion mixture was cleaned-up using C18 96-well plates (BioPureSPE Macro 96-Well, 100mg PROTO C18, The Nest Group) and redissolved in 50μl 0.1% formic acid.

Each 96-well plate contained 8 plasma and 4 serum sample preparation controls, and the acquisition workflow included a pooled quality control sample every ∼10 injections. Liquid chromatography was performed using the Agilent 1290 Infinity II system coupled to a TripleTOF 6600 mass spectrometer (SCIEX) equipped with IonDrive Turbo V Source (Sciex). A total of 5μl was injected, and the peptides were separated in reversed phase mode using a C18 ZORBAX Rapid Resolution High Definition (RRHD) column 2.1mm x 50mm, 1.8μm particles. A linear gradient was applied which ramps from 1% B to 40% B in 5 minutes (Buffer A: 0.1% FA; Buffer B: ACN/0.1% FA) with a flow rate of 800μl/min. For washing the column, the organic solvent was increased to 80% B in 0.5 minutes and was kept for 0.2 minutes at this composition before going back to 1% B in 0.1 min. The mass spectrometer was operated in the high sensitivity mode. The DIA/SWATH method consisted of an MS1 scan from m/z 100 to m/z 1500 (20ms accumulation time) and 25 MS2 scans (25ms accumulation time) with variable precursor isolation width covering the mass range from m/z 450 to m/z 850 ^7^. An IonDrive Turbo V Source (Sciex) was used with ion source gas 1 (nebulizer gas), ion source gas 2 (heater gas) and curtain gas set to 50, 40 and 25, respectively. The source temperature was set to 450 and the ion spray voltage to 5500V.

The data were processed with DIA-NN^103^, an open-source software suite for DIA / SWATH data processing (https://github.com/vdemichev/DiaNN, commit 4498bd7) using a two-step spectral library refinement procedure as described previously^7^, with filtering at precursor level q-value (1%), library q-value (0.5%) and gene group q-value (1%). Highly hydrophobic peptides (reference retention time > 110 on the iRT scale) were discarded. Batch correction was performed at the precursor level as described previously^7^, using linear regression for intra-batch correction and control samples for inter-batch correction. Protein quantification was subsequently carried out using the MaxLFQ algorithm^104,105^ as implemented in the diann R package (https://github.com/vdemichev/diann-rpackage). The Generation Scotland cohort proteomics raw data, which we described previously^7^, have been reanalyzed using the updated software pipeline, to ensure comparability.

### Statistical analysis and multiple-testing correction

Statistical testing was performed in the R environment for statistical computing, version 3.6.0^106^. All protein and clinical laboratory measurements (except for standard and actual base excess) were first log2-transformed. For differential abundance testing, only protein groups matched to at least three different unmodified peptide sequences were considered. Significance testing for a zero median (for analysing trajectories) or against binary variables (worsening, death) was performed using the Wilcoxon W test or Mann-Whitney U test, respectively, as implemented in the “wilcox.test” function of the “stats” R package. Testing against a continuous variable (e.g. when determining significance of pairwise correlations) was performed using the Kendall Tau test, with the slope estimated using the Theil-Sen method, as implemented in the “kendallTrendTest” function of the “EnvStats”^107^ package. When covariates had to be taken into account, we used linear modelling with the “limma”^28^ R package, with P-values obtained using “eBayes”^108^. Modelling with “limma” was likewise used to correct for these covariates for visualisation purposes. WHO grade was considered as a “factor-type” covariate (resulting in a “limma” design matrix with one-hot encoding for different WHO grades). Multiple-testing correction was performed using the Benjamini-Hochberg false discovery rate controlling procedure^109^ as implemented in the “p.adjust” function of the “stats” R package. The adjusted p-values below 0.05 were considered significant. Multiple-testing correction for differential abundance analysis was performed separately for proteins with MRM assays available^68^, the rest of proteins measured, the clinical laboratory measurements and the clinical factors (age, Charlson score, BMI, Horowitz index and FiO2, SOFA score), to ensure that the false discovery rate stayed below 0.05 for each of these categories of features. Likewise, when determining the significance of correlations in correlation matrices, correction was performed for each row or each column separately, to ensure less than 5% false discoveries in each row or column, respectively. For correlation map visualisations, black points were used to indicate row-wise significant correlations, and black rectangles at the border of the respective cell - column-wise significant correlations.

Quantities of gene products corresponding to open reading frames named IGxx (i.e. different types of immunoglobulin chains) were summed together to generate quantities representative of the overall levels of immunoglobulin classes (IGHVs, IGLVs, etc). This does not affect any conclusions of this work and was done purely to improve visualization and simplify the interpretation of the heatmaps and correlation maps. Full protein level tables, including levels of individual immunoglobulin gene products, are provided in supplementary materials.

### Identifying markers of the disease severity

The first time point measured at the maximum WHO grade was chosen for each patient. For each omics feature, its log2-transformed values were tested for a trend depending on the WHO grade. Age was included as a covariate in the linear model as described above.

### Identifying markers with vary with age in COVID-19

The first sampling time point measured was chosen per patient. For each omics feature, its log2-transformed values were tested for a trend depending on age. The test was performed either using the Kendall Tau test (as described above; Supp. Fig. SF8, Supp. Fig. SF9), or by accounting for WHO grade as a covariate in the linear model (as described above; Supp. Fig. SF10, Fig. 5).

### Identifying markers of RRT and ECMO

For each omics feature, the P-value was calculated using the Mann-Whitney test, comparing between the median log2-levels across all sampling time points at WHO grade 7 in patients who did not receive the therapy and the median log2-levels across all sampling time points at WHO grade 7 after initiation of the respective therapy in patients who did.

### Identifying markers predictive of the remaining time in hospital

Patients, for which the first sampling time point before the outcome corresponded to the WHO = 3 severity grade (that is the patient did not require supplemental oxygen on that day), were considered. Thus, no correction for disease severity was necessary. Testing of log2-levels of each omics feature (measured for the first sampling time point) vs the remaining time in hospital (days) was performed by including age as a covariate in the linear model as described above.

### Identifying markers predictive of disease worsening

The first sampling time point measured was chosen per patient. Future disease worsening was defined as a future increase in the WHO grade (for patients at WHO grade < 7) or death (for patients at WHO grade 7). For each omics feature, its log2-levels were compared between patients who did not worsen and patients who did, with age and current WHO grade (as factor) included as covariates in the linear model as described above.

### Peak period of the disease definition

When studying the dynamic changes in omics values during the disease course, we focused on the time points sampled when the disease was the most severe for a particular patient. For each patient, we thus defined the “peak period of the disease” as the time when the patient was receiving the most intensive treatment during their stay in hospital, that is the time when the patient was at WHO grade 6 or 7, for patients who received invasive mechanical ventilation at some point, or otherwise at their maximum WHO grade (3, 4 or 5).

### Identifying proteins changing with time at the peak of the disease

Only patients with at least two days between the first and last sampling time points at the peak of the disease (as defined above) were considered. For each omics feature, a linear regression model was fitted for its log2-levels vs the day number, and the quantity slopeadj = (linear regression slope) * (number of days between first and last time points) was calculated. A Wilcoxon W test was then applied to compare the median of slopeadj to zero. The values of slopeadj for each feature are visualised in Supp. Fig. SF13.

### Correlation maps

General correlation maps were generated using the log2-transformed values of features at the first time point measured at the maximum WHO grade for each patient. The correlation map between feature changes during the peak of the disease (as defined above) was generated by correlating the slopeadj values (as defined above). The map of significant protein correlations not detected in the general population was generated by excluding all correlations which were either significant (P <= 0.05, without multiple-testing correction) with the same trend in the Generation Scotland cohort, or could not be calculated reliably therein (less than 20 valid points).

### Identifying omics trajectories that are predictive of survival in critical patients

For each omics feature, the difference between its log2-levels at the last and the first sampling timepoints at the peak of the disease (as defined above) was considered. Among patients which were at WHO = 7 severity grade at some point, the distribution of this difference between survivors and non-survivors was compared using the Mann-Whitney U test. Patients still in critical condition on 16 September 2020 were excluded (115, 129). Patients with therapy limitations according to (presumed) patients’ wishes (DNI, secondary DNR) were likewise excluded (26, 52, 59, 83, 90).

### Prediction of mechanical ventilation with machine learning

To reflect the power of omics measurements in characterising the phenotype, a classifier was constructed to predict mechanical ventilation (at the present time point) using either the proteome or the clinical data. All protein and clinical laboratory measurements were log2-transformed. The first time point measured at the maximum WHO grade was selected per patient. We used a gradient boosted tree algorithm implemented in the XGBoost 1.2.0^110^ under Python 3.7.4. The classifier was based on the construction of 500 gradient boosted trees using leave-one-out cross-validation and the “exact” tree method. To circumvent overfitting the shrinkage factor “eta” was set to 0.05 using a subsampling of 0.5 of the training data. In addition, the L2 regularization term “lambda” was set to 100. For the assessment of classifier performance, the leave-one-out method was applied in the following way: the prediction was made for each sample separately, by excluding (withholding) this sample from the dataset, training the classifier on the remaining (independent) samples and then predicting the withheld sample using the trained model. For the determination of the feature importances one classifier was trained on all data points using only the proteome measurements and the same setup as described above. The feature importance scores were then extracted directly from the trained classifier.

### “Proteome severity” score calculation

A proteome severity score was calculated for each sample, to be used for visualisation in Fig. 3c. This was done via a LASSO model (as implemented in the cv.glmnet() function of the “glmnet”^111^ R package), trained on the proteomes corresponding to the first time point acquired for each patient, to predict the current WHO grade. For this, all protein measurements were first log2-transformed. Only proteins quantified with at least three different peptides were considered, on which minimal-value imputation was used.

### Prediction of survival using parenclitic networks

The first time point measured at the maximum WHO grade was selected per patient. Only patients with maximum WHO grade 7 were considered. Patients still critical on 16.09.2020 were excluded (115, 129), as well as patient 26, due to the cause of death unrelated to COVID-19. To reduce the feature space used as input for the machine learning model, we limited it to the quantities of 57 proteins which are FDA-approved biomarkers with MRM assays available^68^ and which were quantified with at least three different peptides in this study. Missing values were imputed using minimal value imputation, and the data were standardized.

Machine learning was carried out using the parenclitic networks approach^52,53^. Briefly, during training, for each pair of features, a radial SVM classifier is trained (using the svm() function from the “e1071” R package with default settings). For each sample, a network is then built, wherein vertices correspond to features and the edge weight is the death probability as predicted by the SVM classifier. Maximum, mean and standard deviation of the edge weights, as well as the numbers of edges with weights greater than 0.5 (i.e. fatal outcome is predicted) and nodes with at least one such edge are calculated. A LASSO classification model (alpha = 0.01) is then constructed on these 5 features using the glmnet() function of the “glmnet”^111^ R package with default settings.

For the assessment of the classifier performance (Charité cohort), a cross-validation method was applied in the following way: the prediction was made for each sample by excluding (withholding) it from the dataset along with two other samples (chosen randomly with the constraint that out of 3 samples one corresponds to a non-survivor and two to survivors), training the classifier on the remaining (independent) samples and then generating predictions for the withheld samples using the trained model. Such leave-3-out partition was generated randomly 50 times and the predictions for each sample were averaged. For the assessment of the performance on an independent dataset (Innsbruck cohort), the classifier was trained on all the Charité samples and used to estimate the probabilities of fatal outcome on the Innsbruck cohort.

## Supplementary Figures and Tables

**Supp. Table 1.**
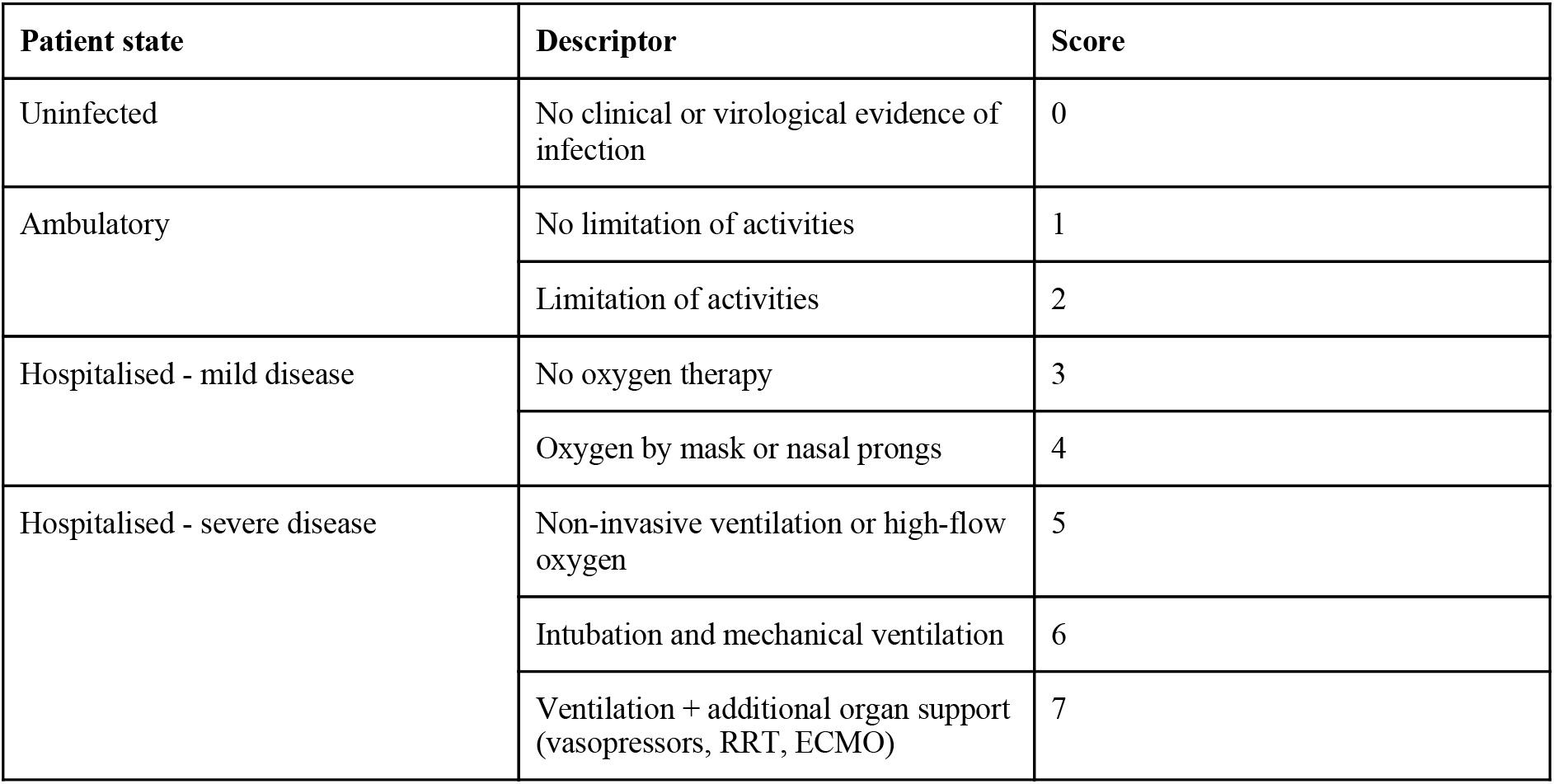
WHO ordinal scale for clinical improvement in COVID-19 as used in the study (World Health Organisation 2020).

| Patient state | Descriptor | Score |
| --- | --- | --- |
| Uninfected | No clinical or virological evidence of infection | 0 |
| Ambulatory | No limitation of activities | 1 |
|  | Limitation of activities | 2 |
| Hospitalised - mild disease | No oxygen therapy | 3 |
|  | Oxygen by mask or nasal prongs | 4 |
| Hospitalised - severe disease | Non-invasive ventilation or high-flow oxygen | 5 |
|  | Intubation and mechanical ventilation | 6 |
|  | Ventilation + additional organ support (vasopressors, RRT, ECMO) | 7 |

**Supp. Table 2.**
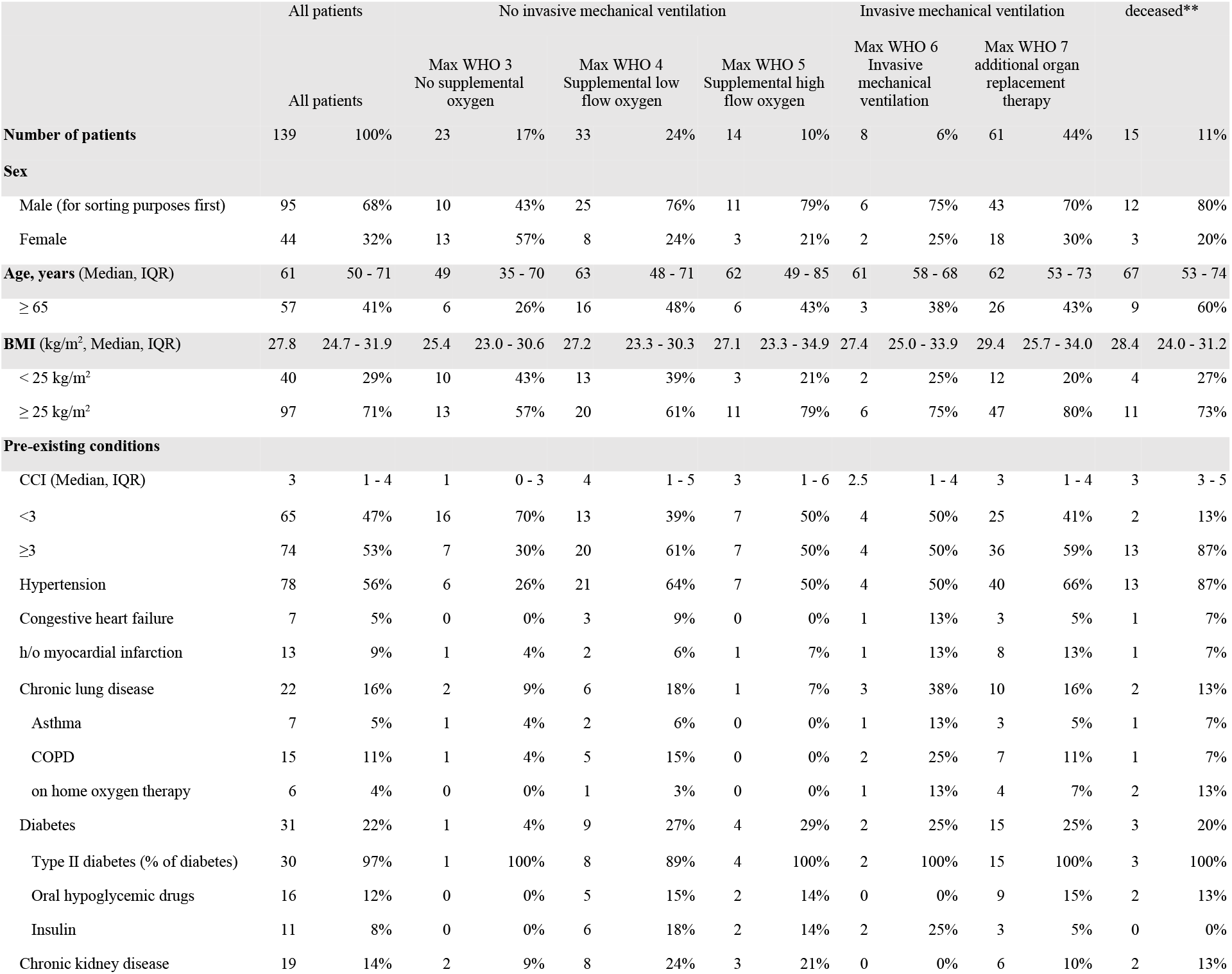
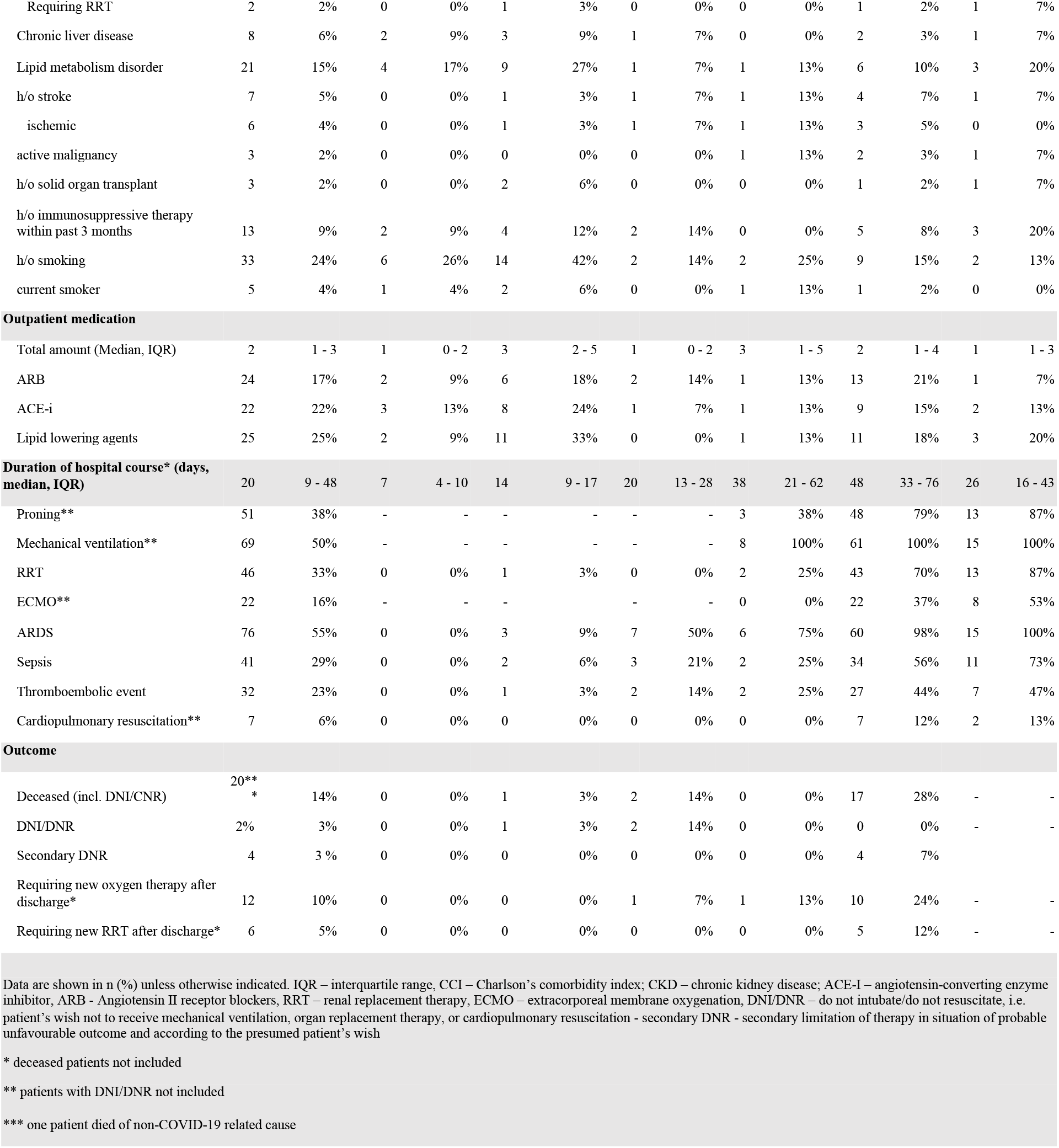
Baseline, treatment and outcome characteristics of patient cohort with COVID-19 at Charité - University hospital Berlin. Patients are stratified according to the maximum grade on WHO ordinal scale.

### The following tables are included as separate supplementary files in a .zip file attached to this medRxiv preprint

Supp. Table 3. Measurements and statistical tests summary.

Supp. Table 4. Correlations of protein and clinical lab measurements (corresponds to Fig. 1b).

Supp. Table 5. All correlations between the omics measurements used in this study (corresponds to Supp. Fig. SF2).

Supp. Table 6. COVID-19-specific protein-protein correlations (corresponds to Supp. Fig. SF3).

Supp. Table 7. Correlations between dynamic changes in omics measurements during the peak period of the disease (corresponds to Supp. Fig. SF11).

**Supplementary files** (Supp. Fig. SF1 - SF22) are provided as separate PDFs in a .zip file attached to this medRxiv preprint. All figures are also provided as separate high-resolution PDFs.

