## Supplementary Figures and Tables for "A time-resolved proteomic and diagnostic map characterizes COVID-19 disease progression and predicts outcome": SF1 - patient trajectories.pdf

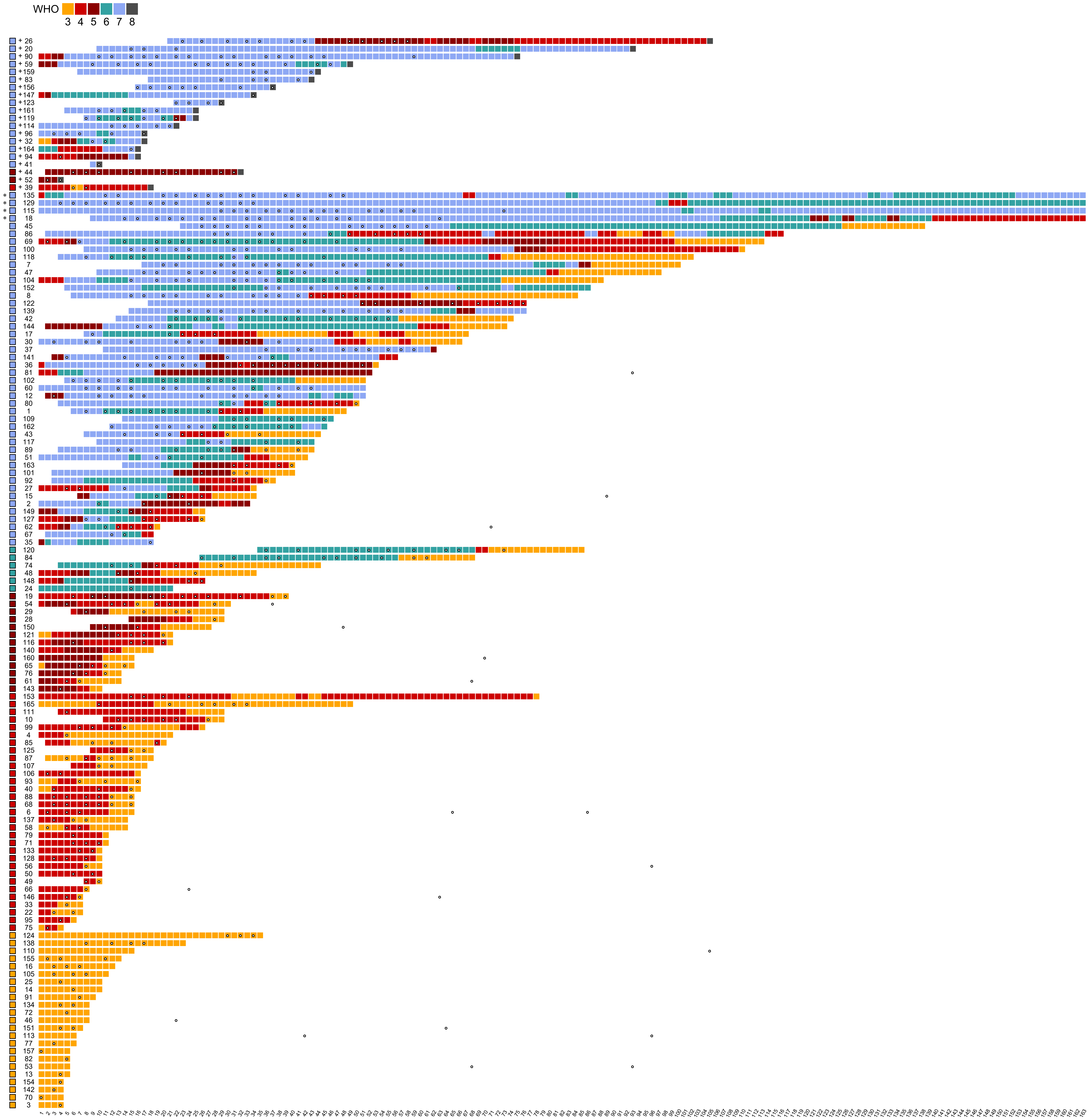

Supp. Figure SF1. **Disease trajectories from hospital admission onward**. Patient IDs are given on the y-axis, number of days since admission on the x-axis, WHO severity grade is color-coded, starting with the day the patient was admitted to Charite or transferred from another hospital. Proteomic samples (including follow-up visits after discharge) are indicated with white points.

+ deceased
* still in hospital on 30 August 2020
