## Supplementary figures and images for "A time-resolved proteomic and diagnostic map characterizes COVID-19 disease progression and predicts outcome"

### Figure 1.pdf

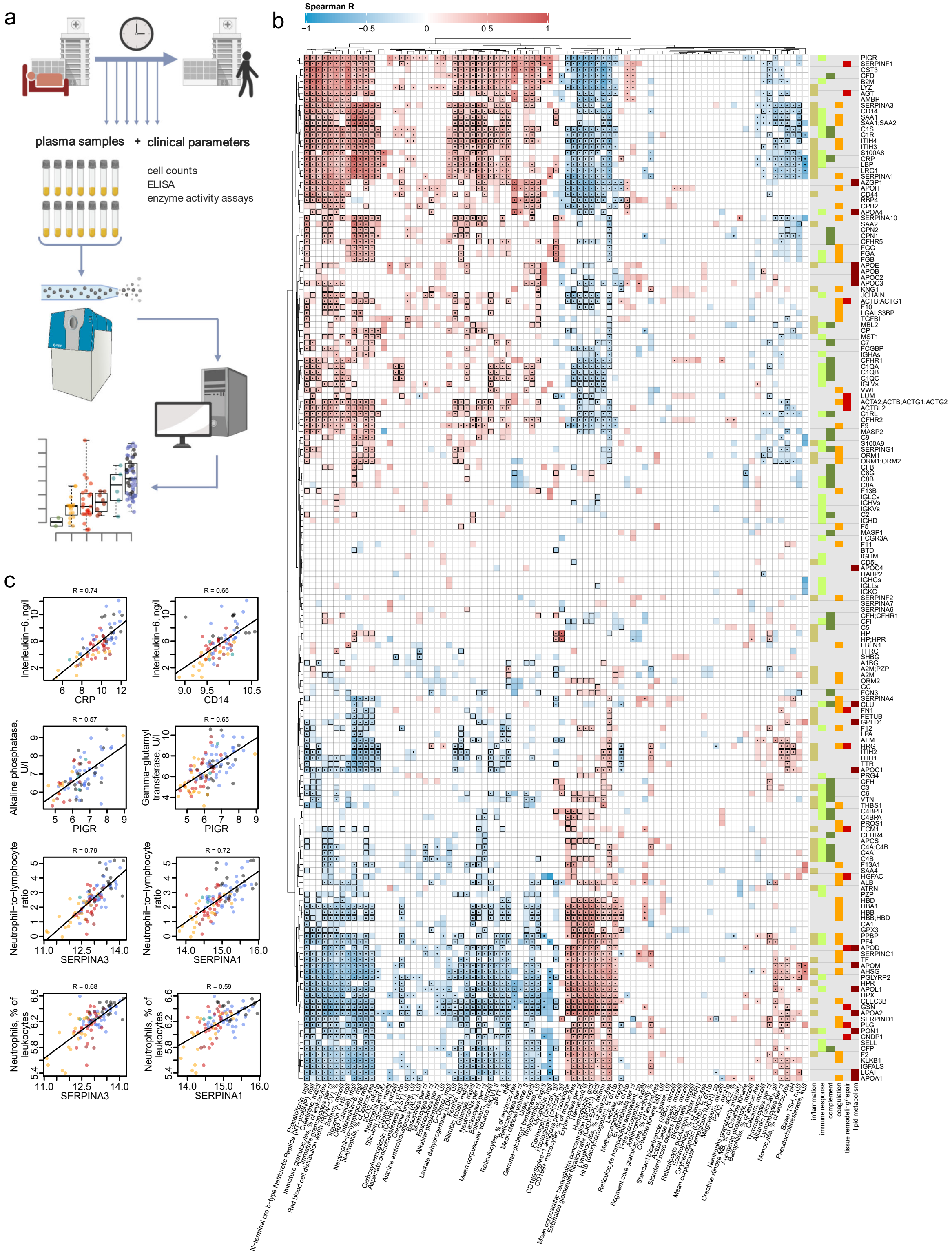

### Figure 2.pdf

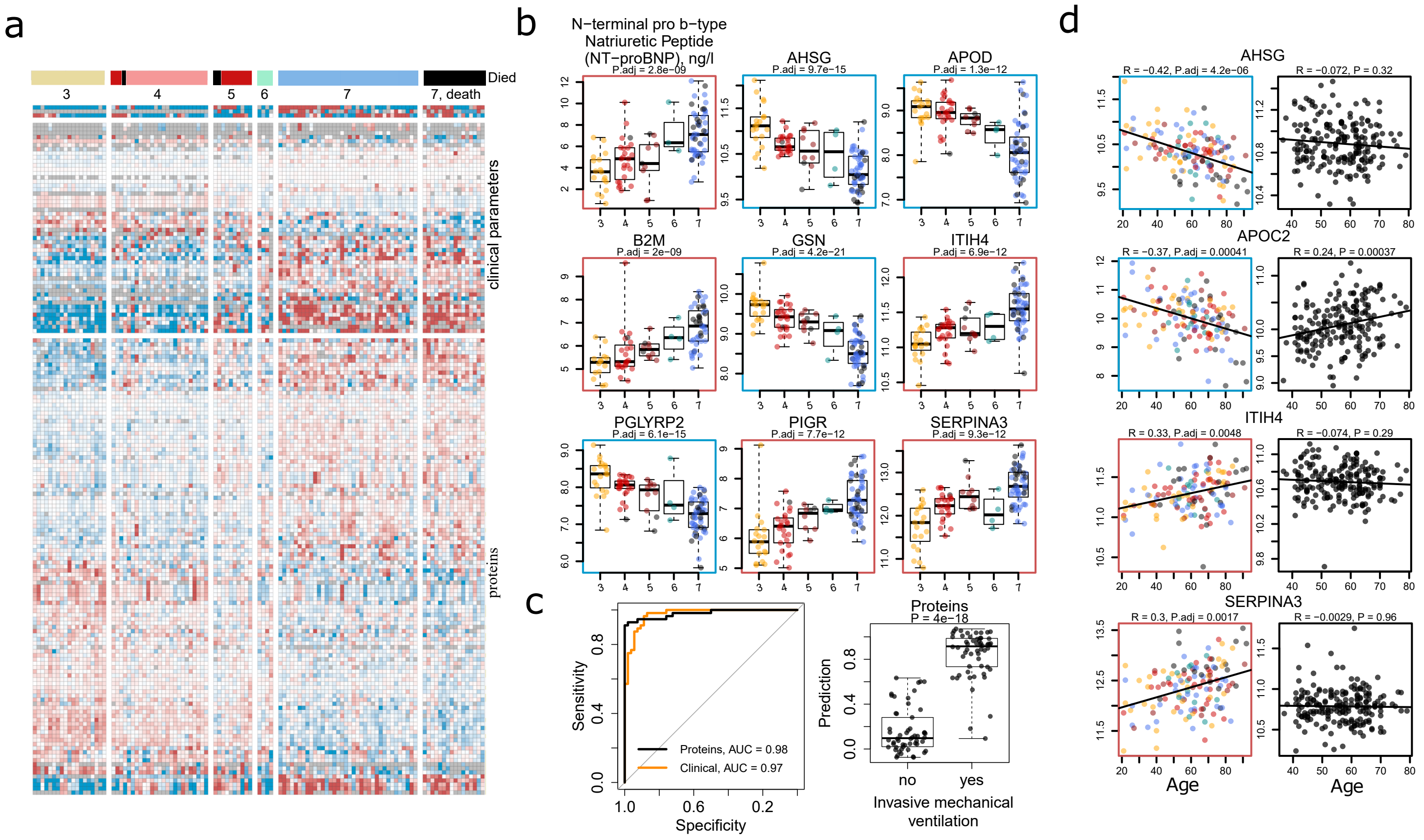

### Figure 3.pdf

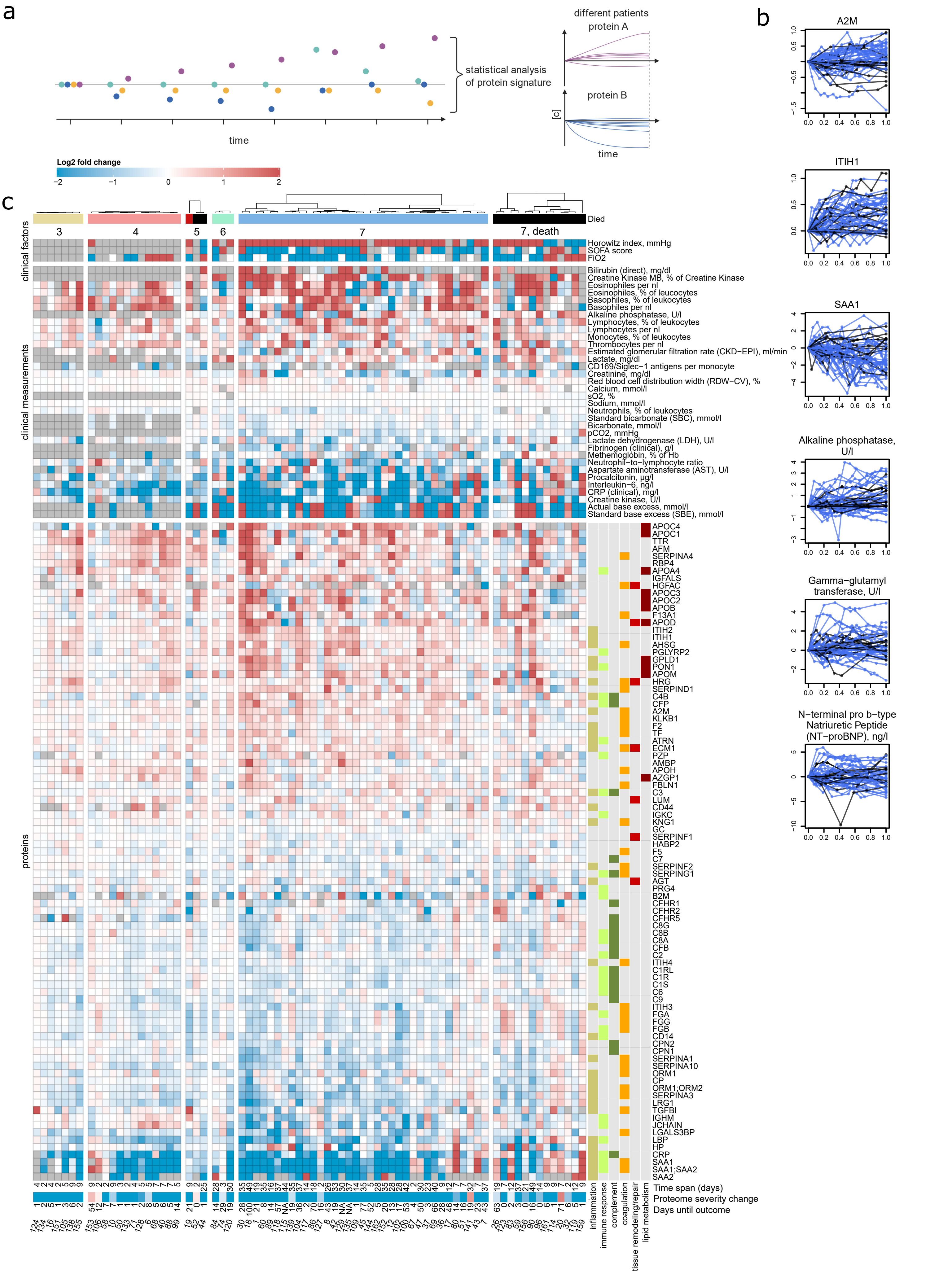

### Figure 4.pdf

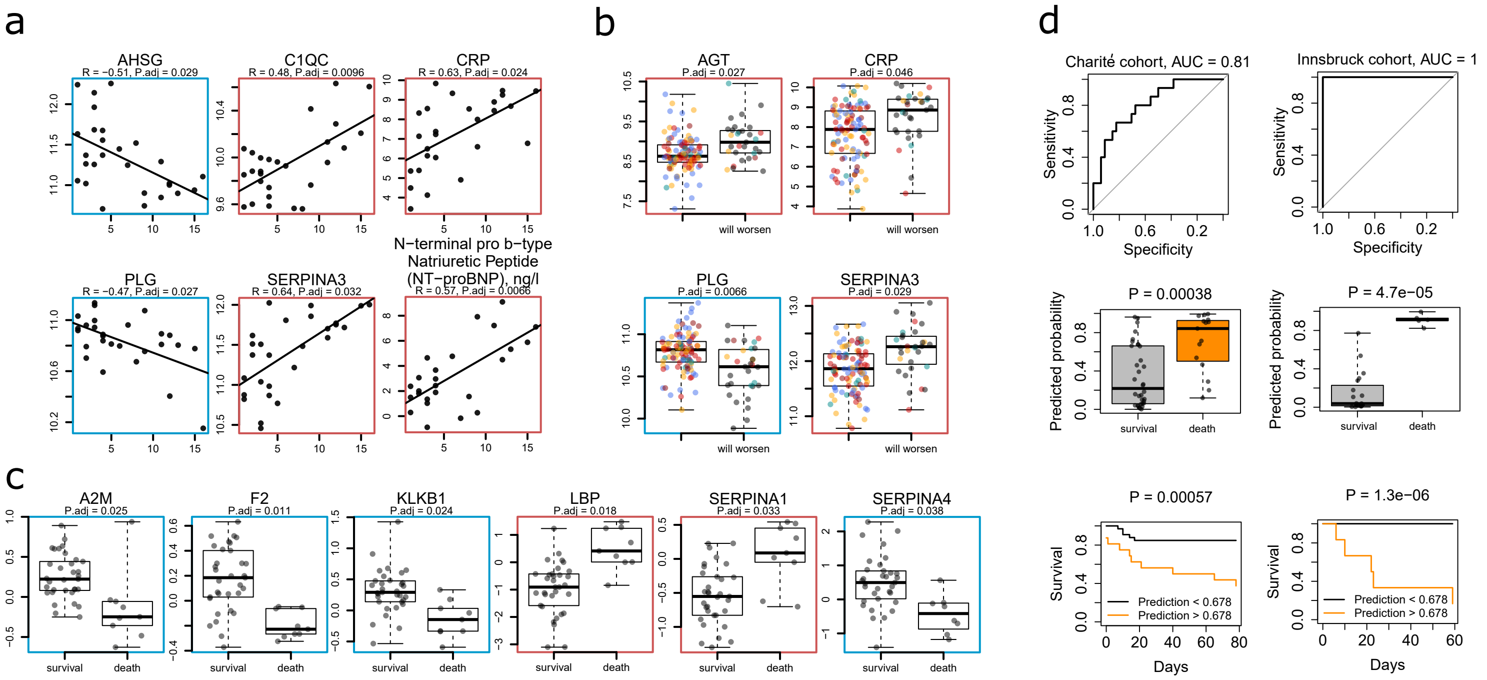

### Figure 5.pdf

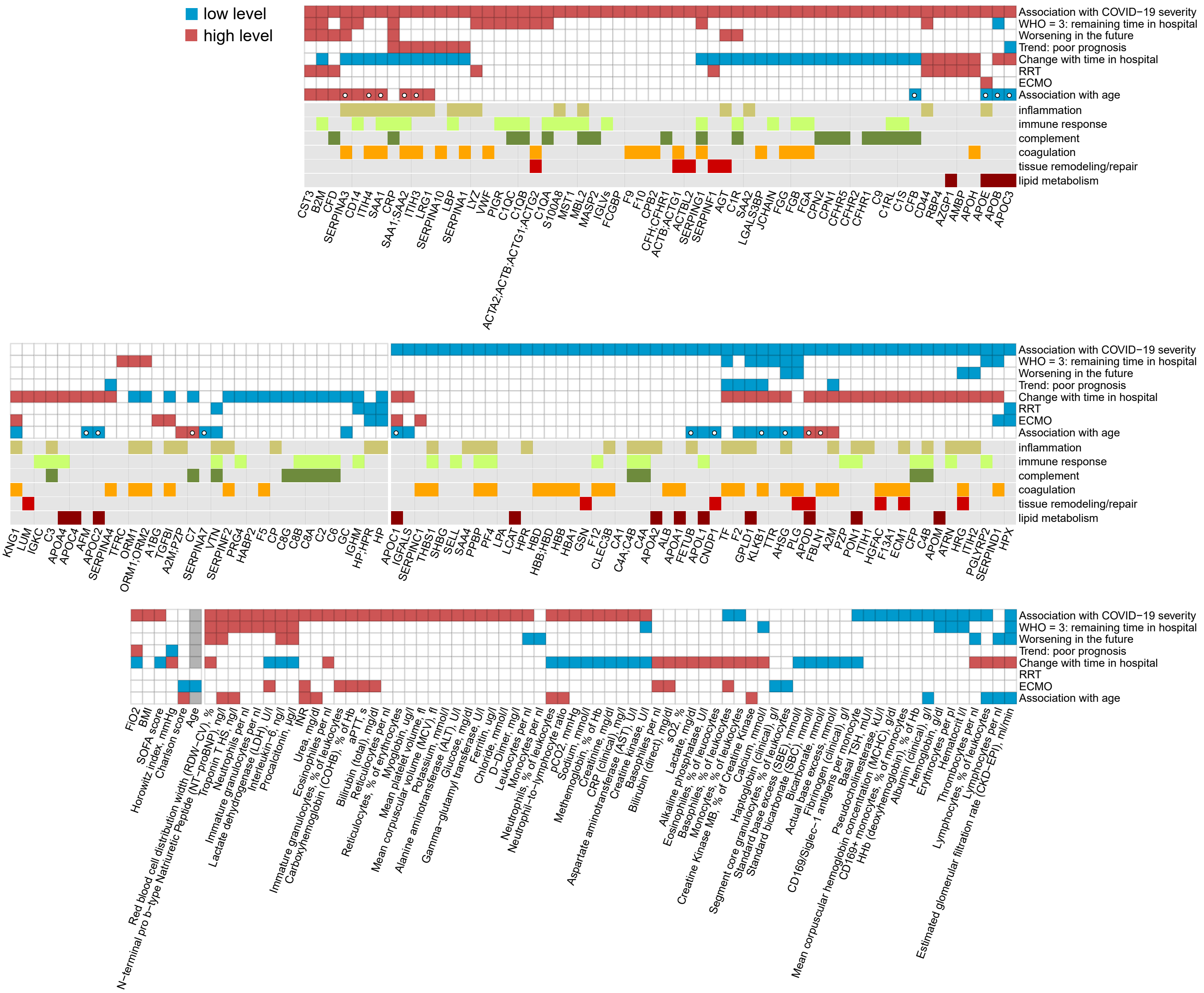

### SF2 - map of all correlations between omics measurements.pdf

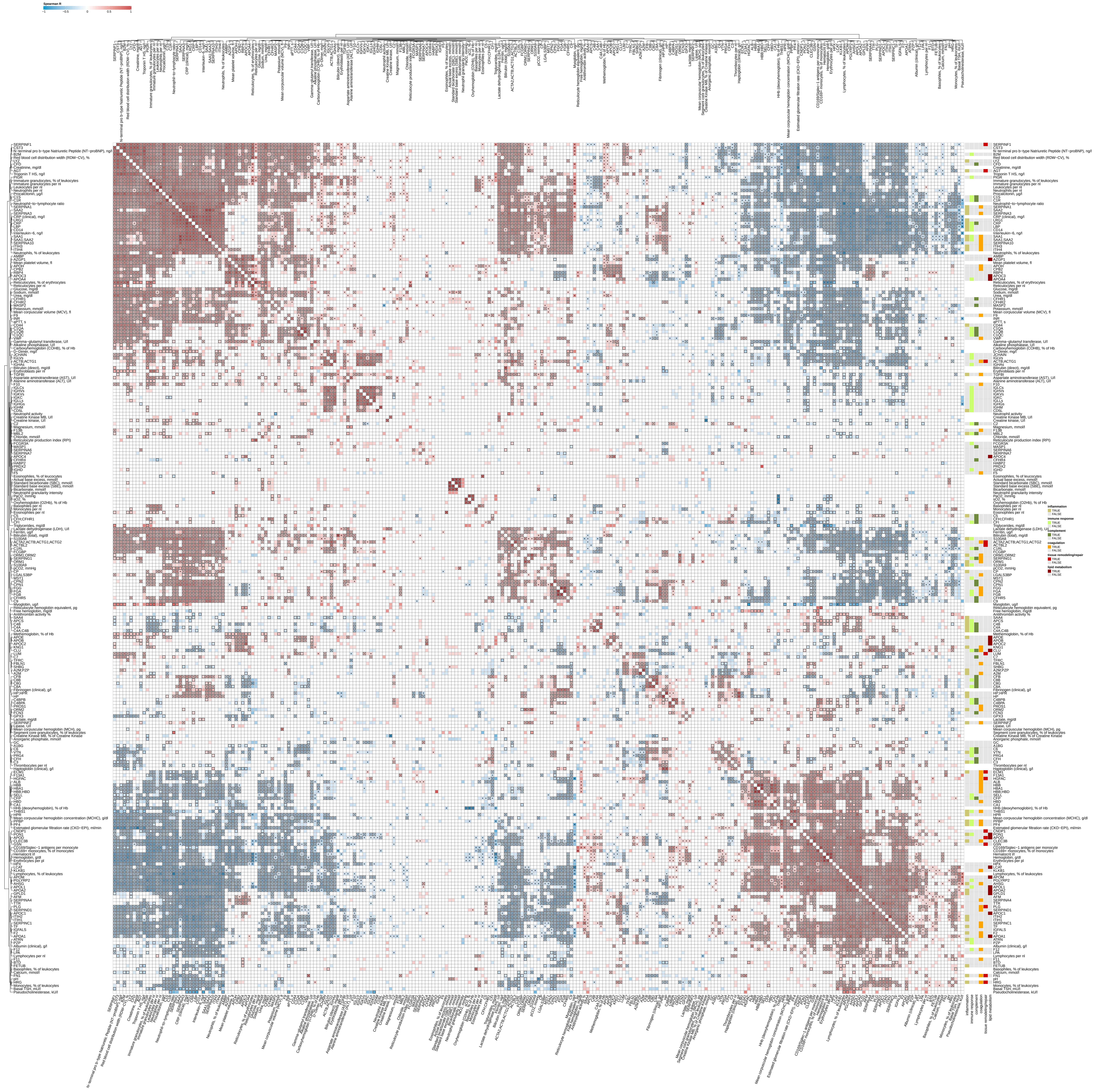

### SF3 - map of COVID-19-specific protein-protein correlations.pdf

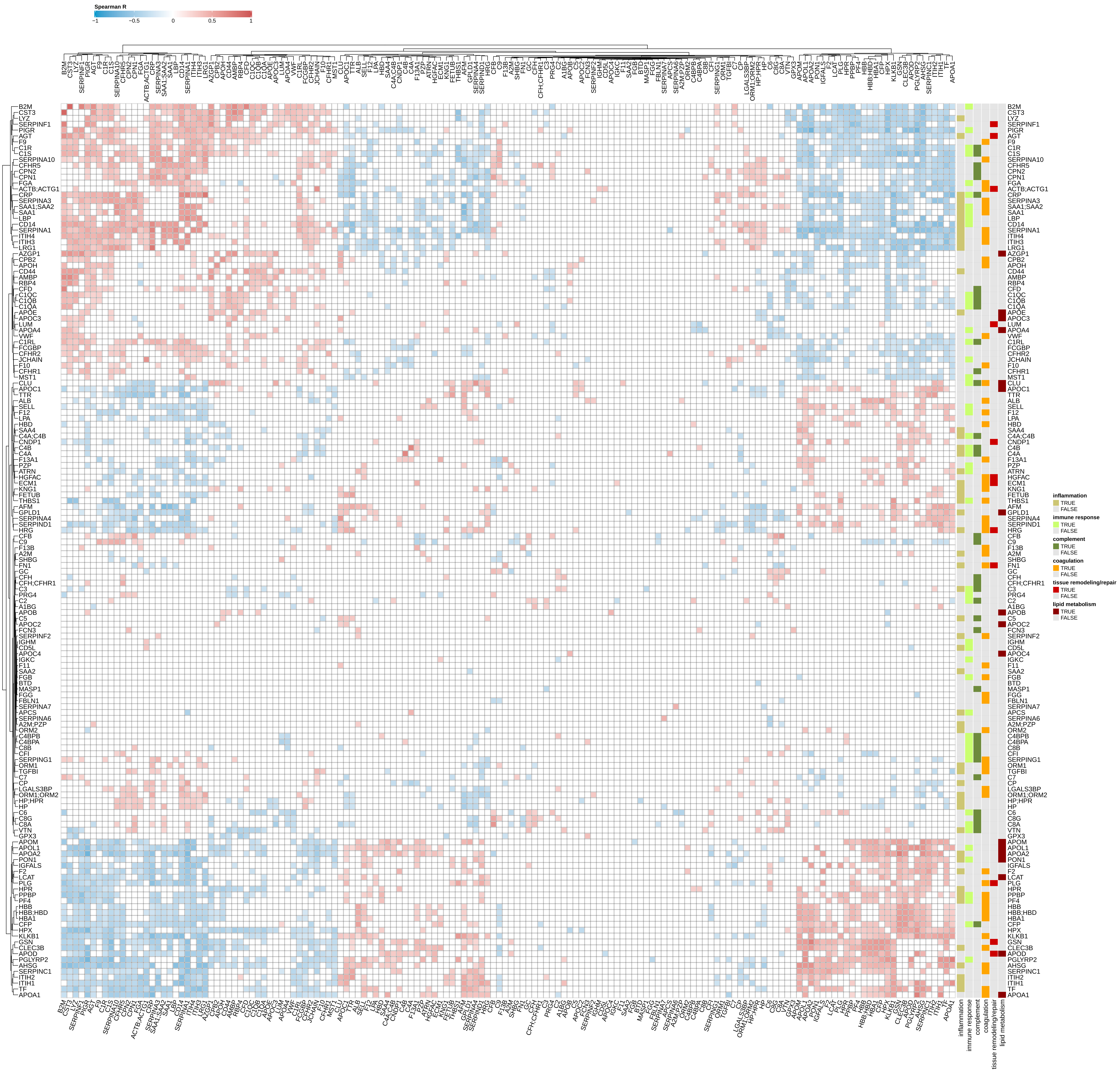

### SF4 - map of omics response depending on COVID-19 severity.pdf

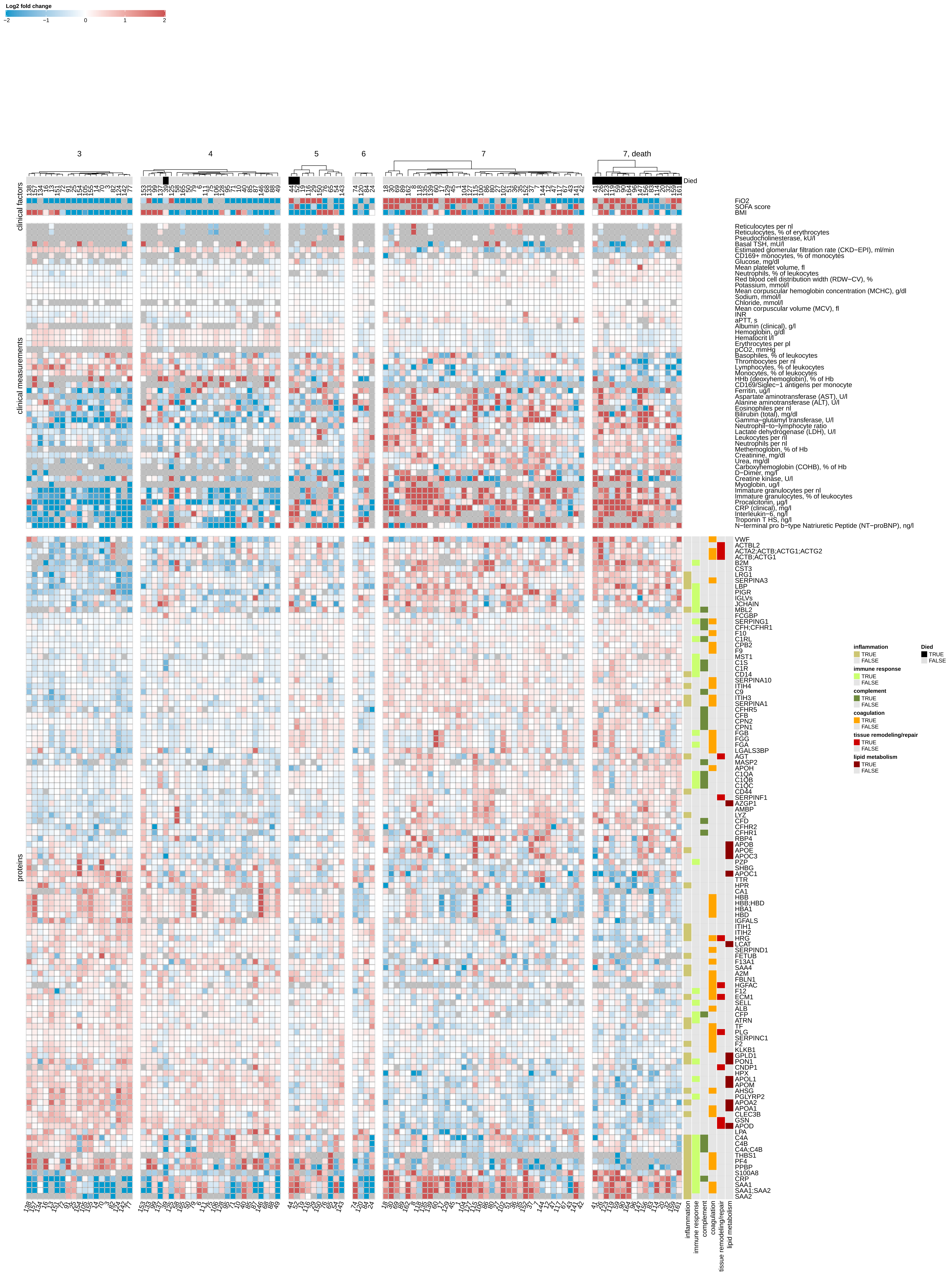

### SF5 - omics features significantly regulated depending on COVID-19 severity.pdf

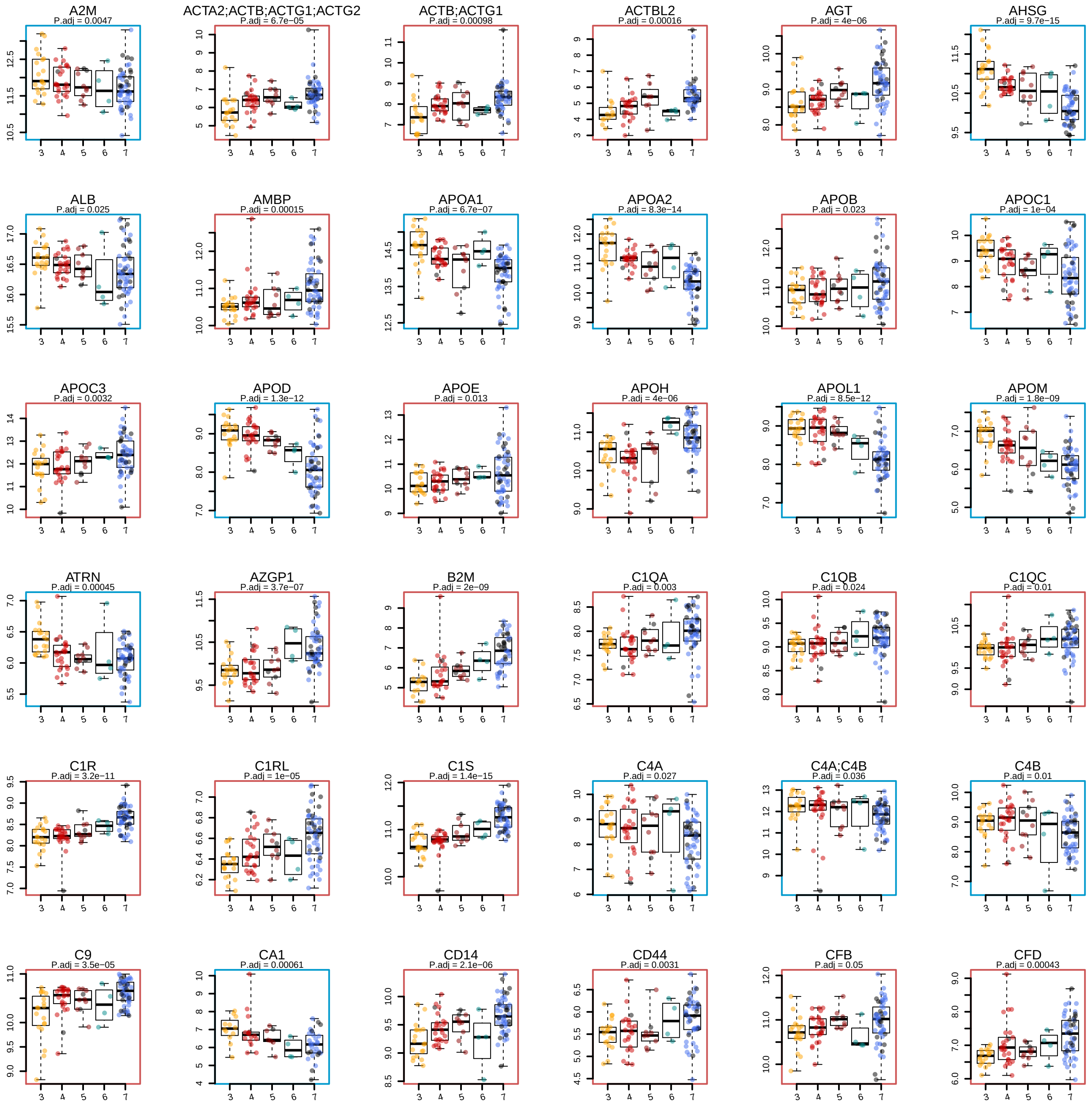

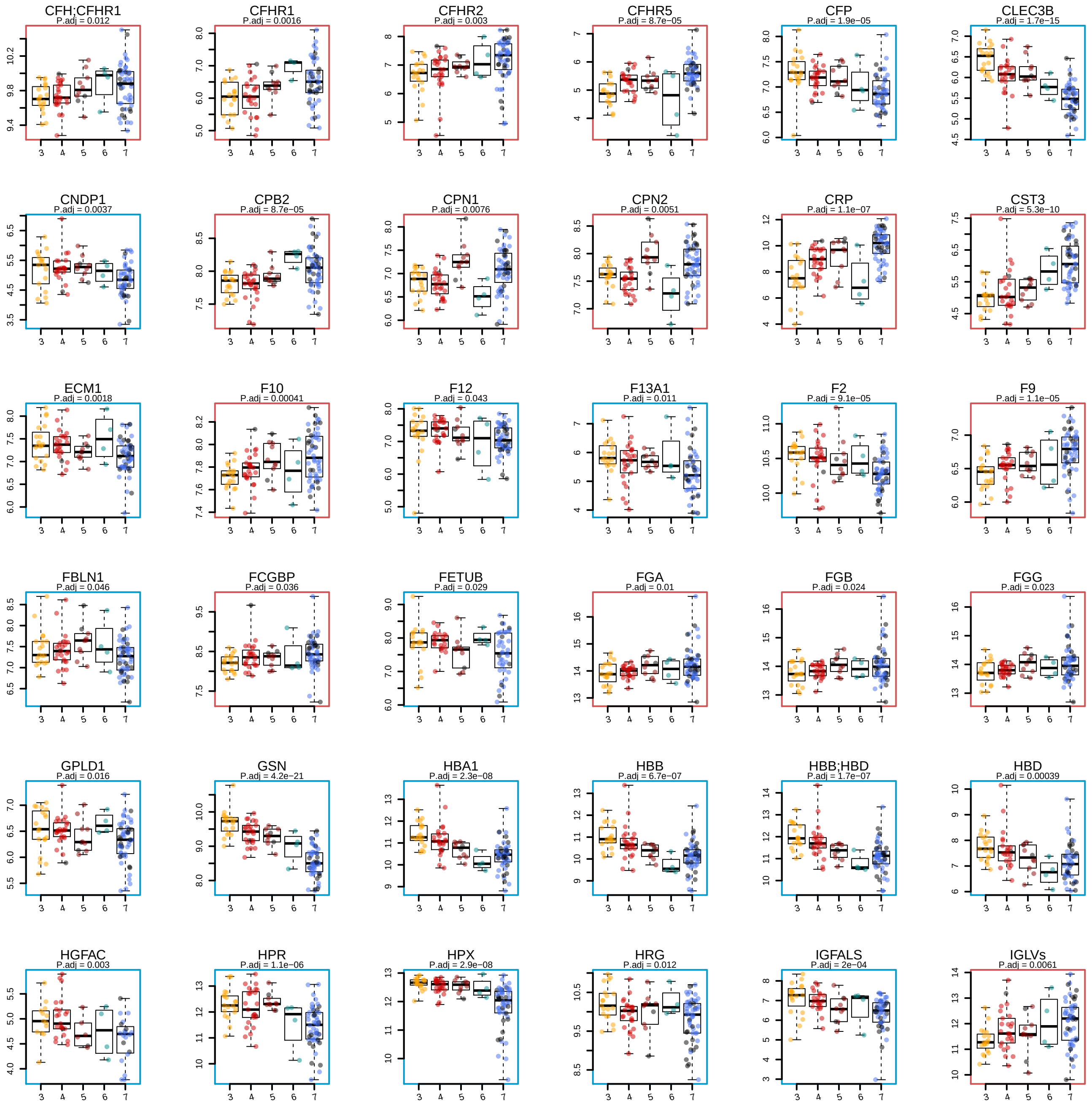

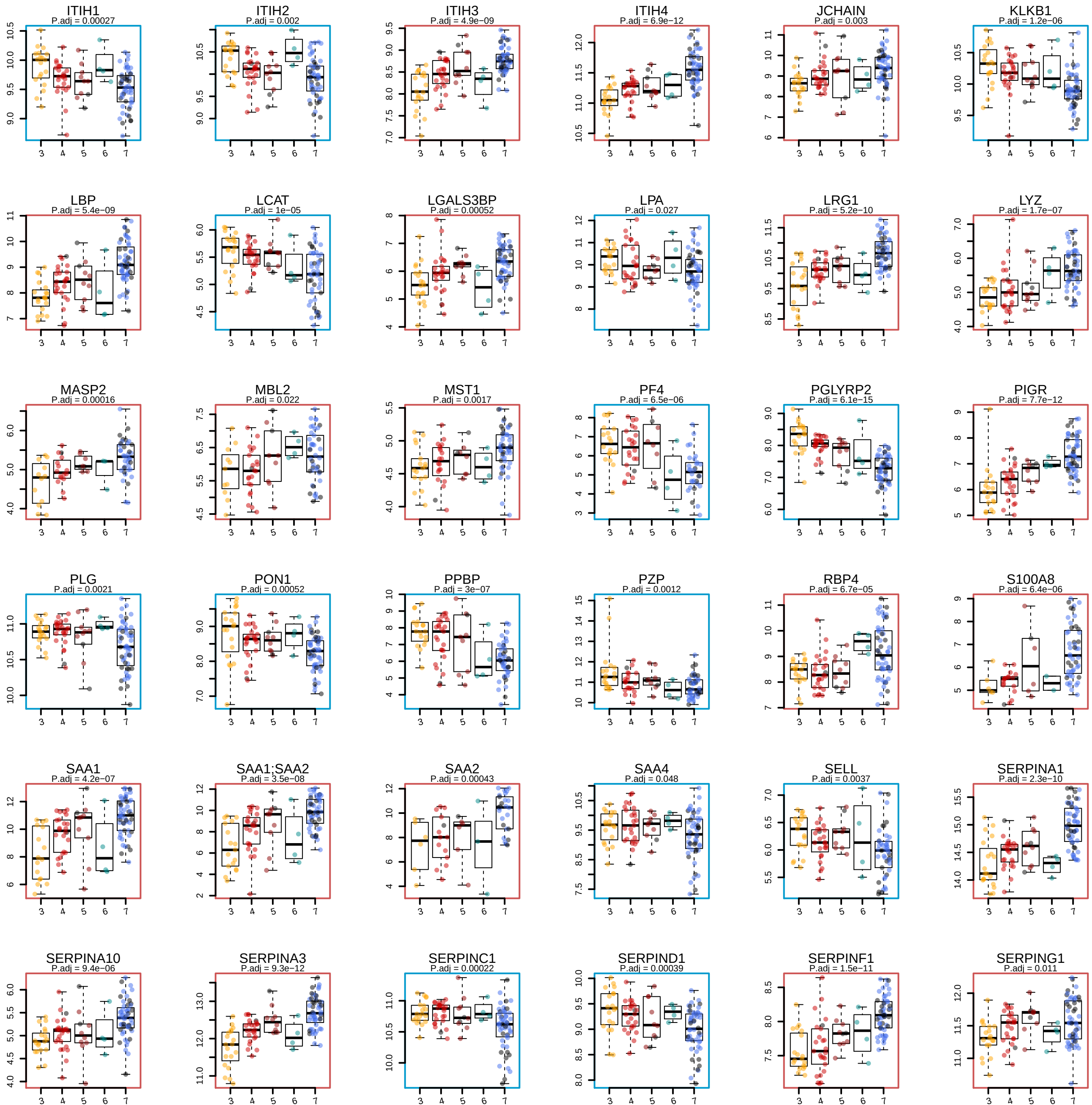

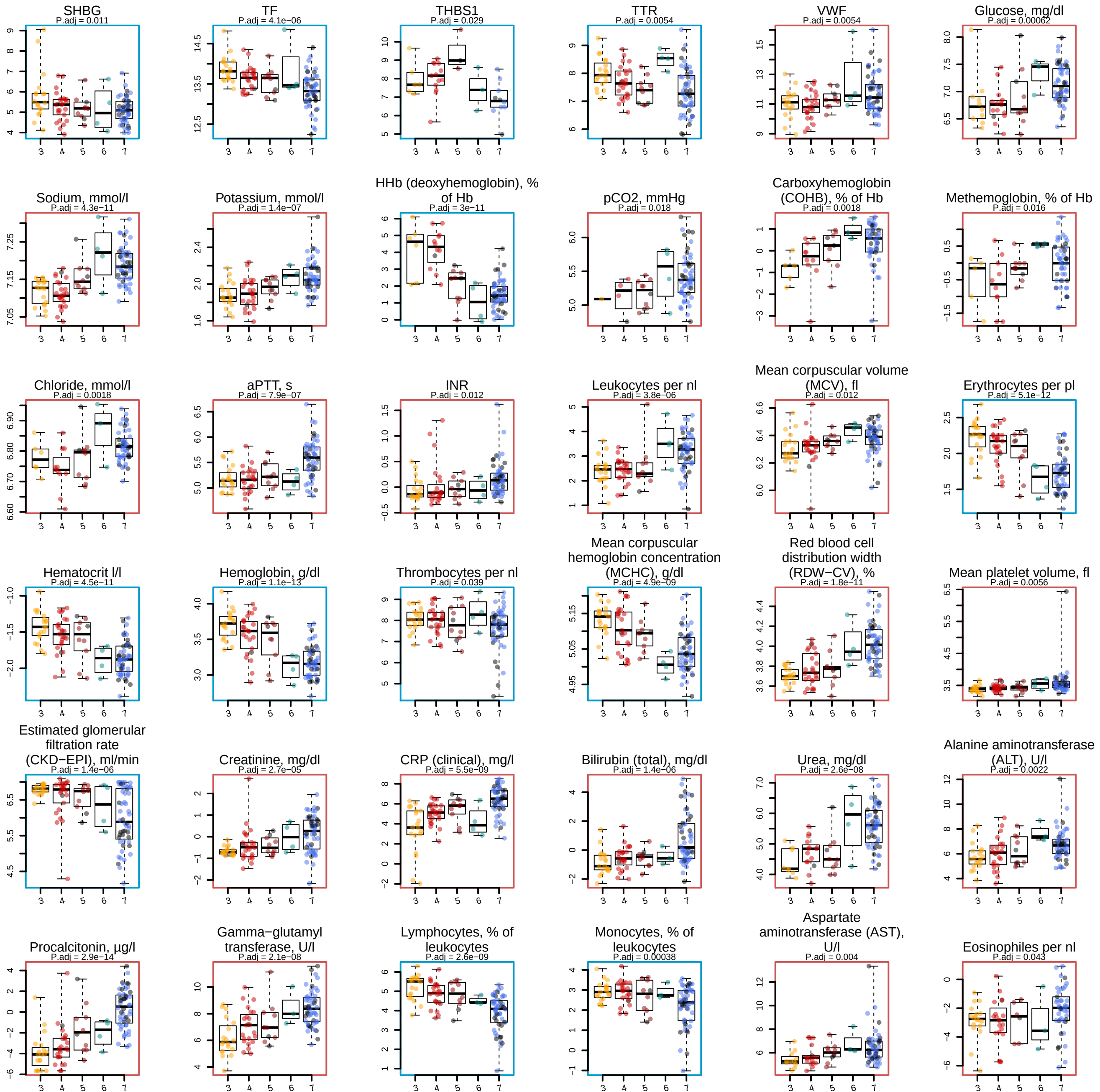

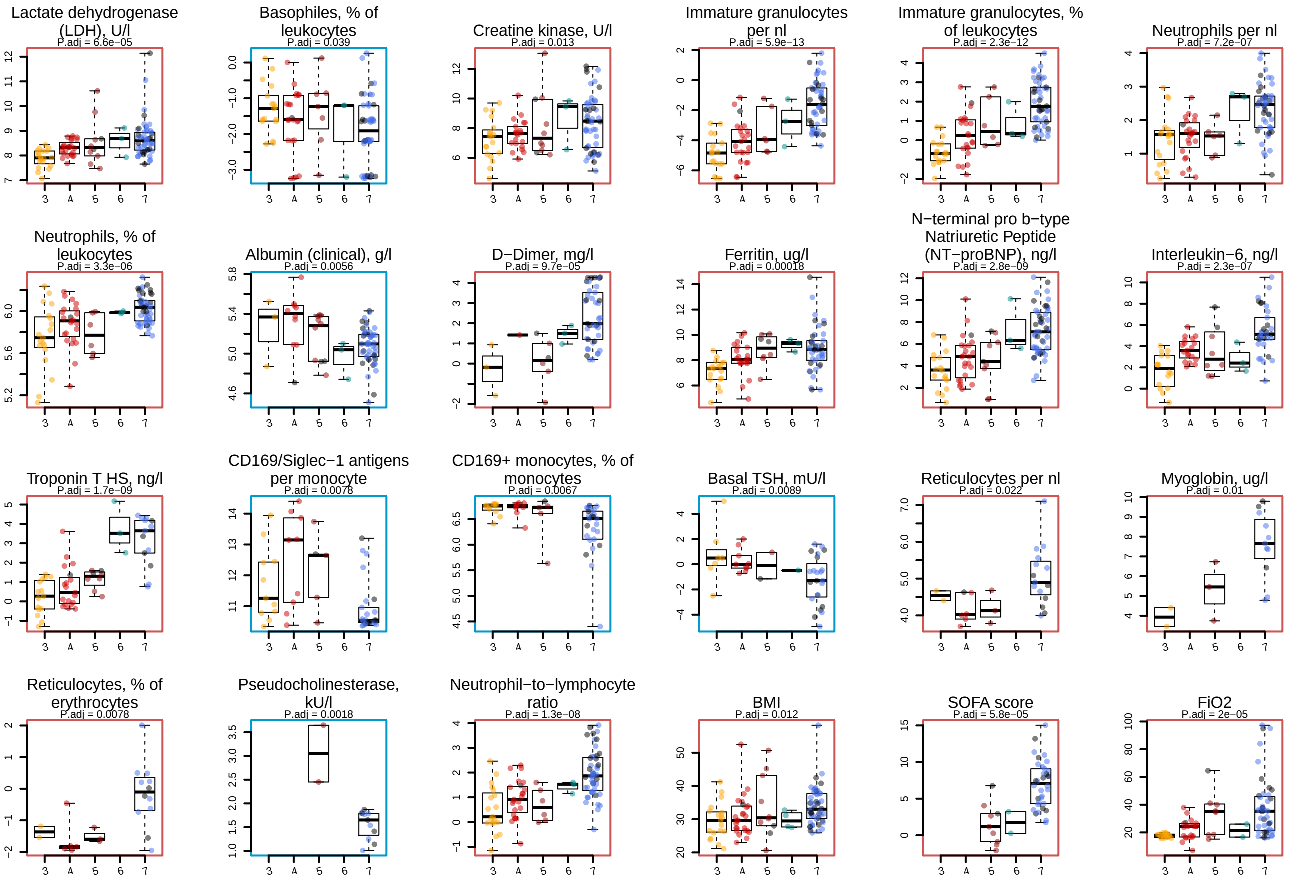

### SF6 - omics features significantly regulated upon RRT.pdf

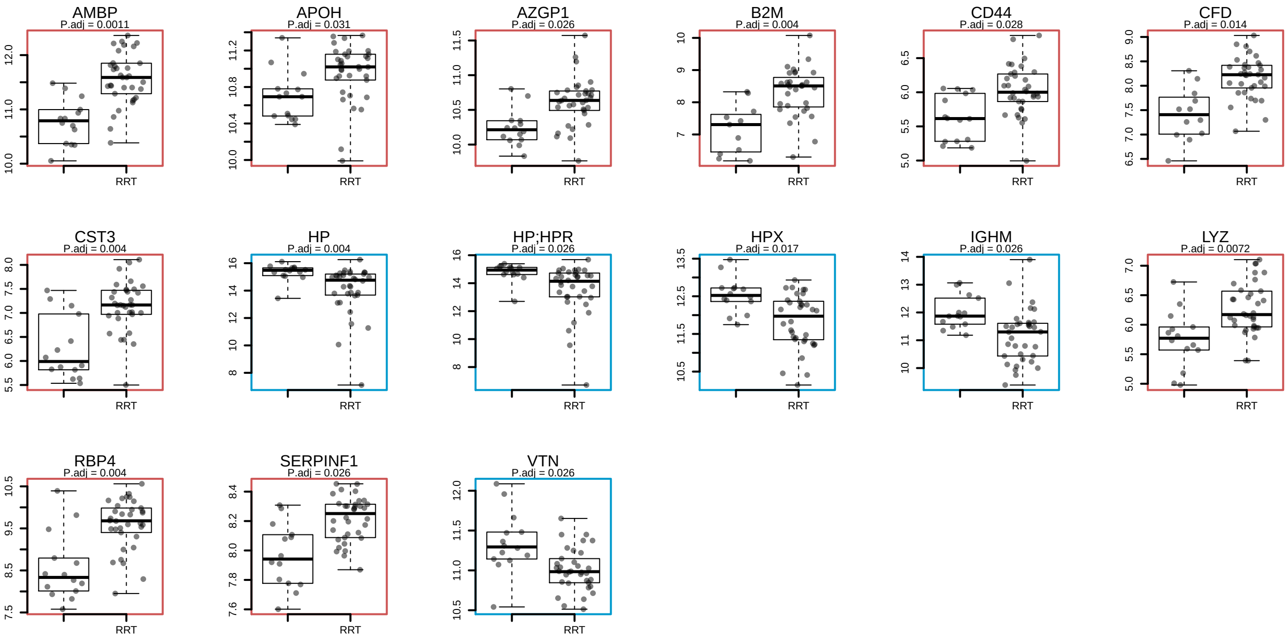

### SF7 - omics features significantly regulated upon ECMO.pdf

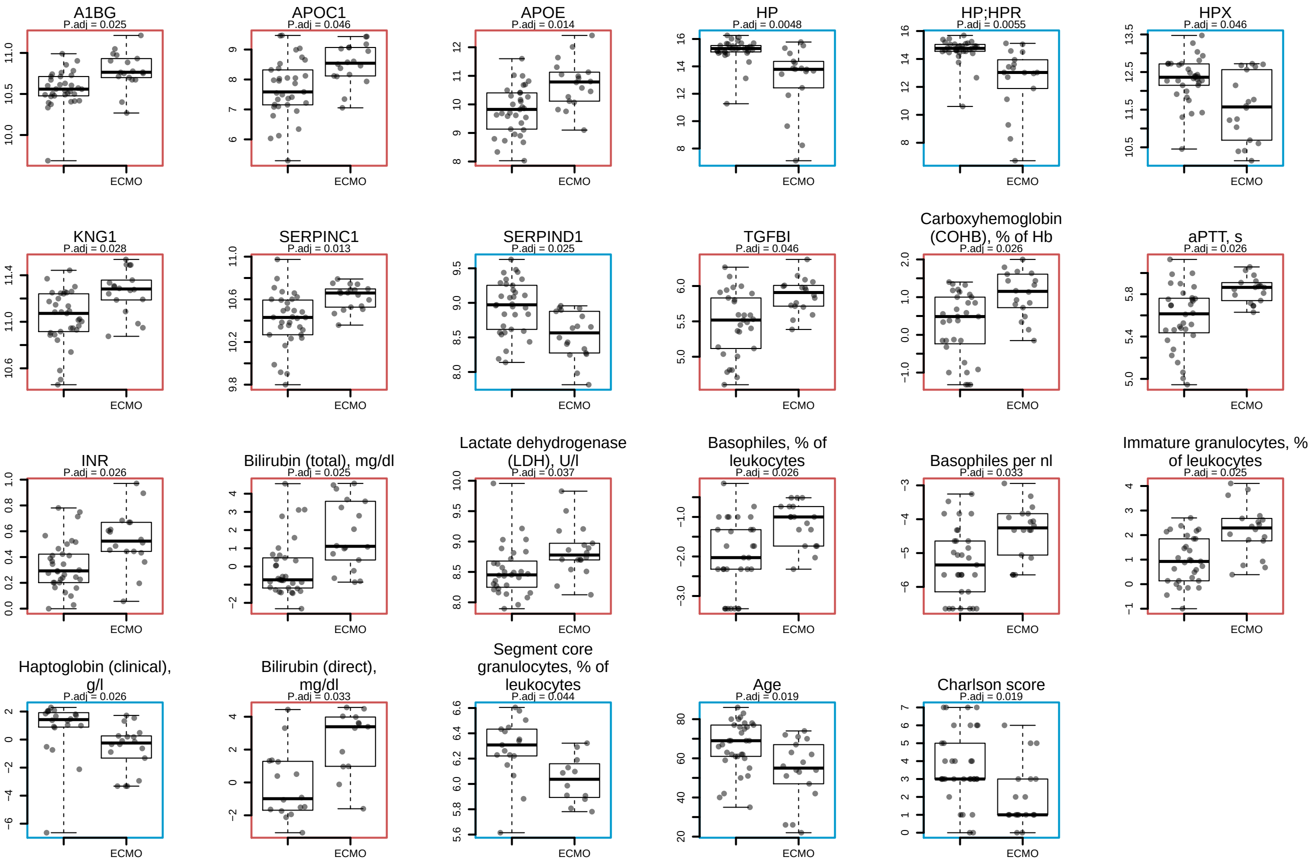

### SF8 - omics features changing with age - test without accounting for the WHO grade as a covariate.pdf

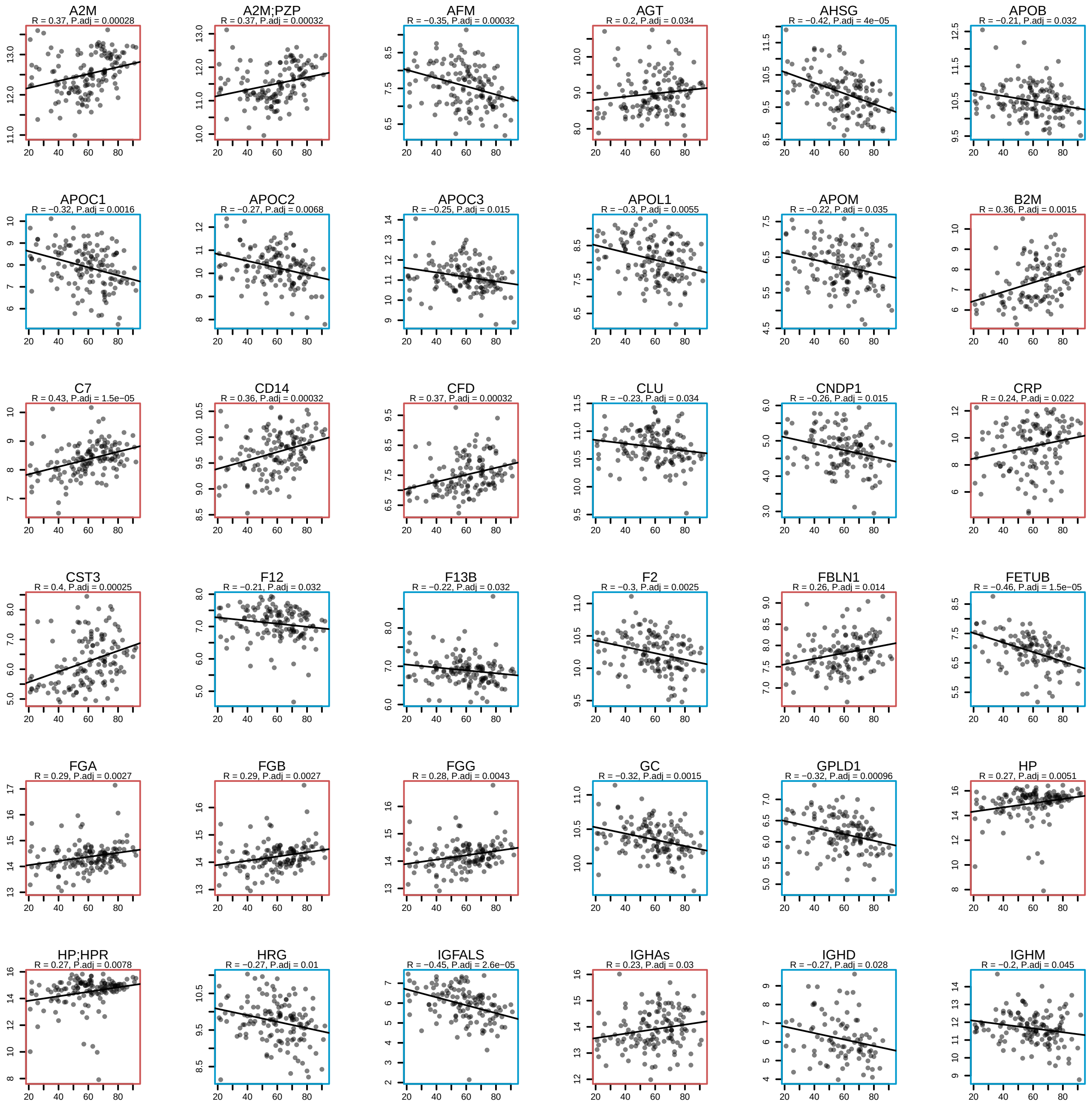

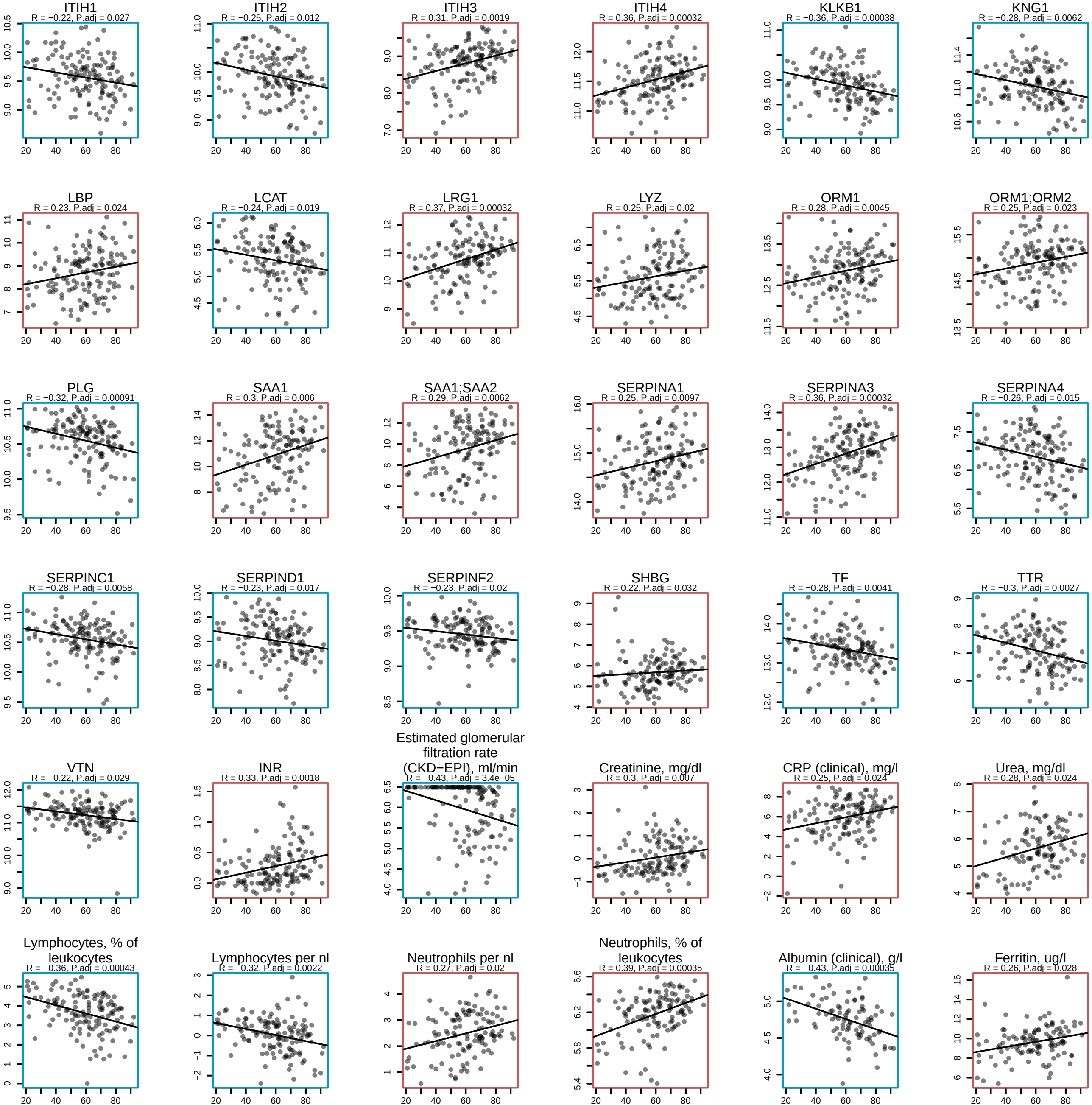

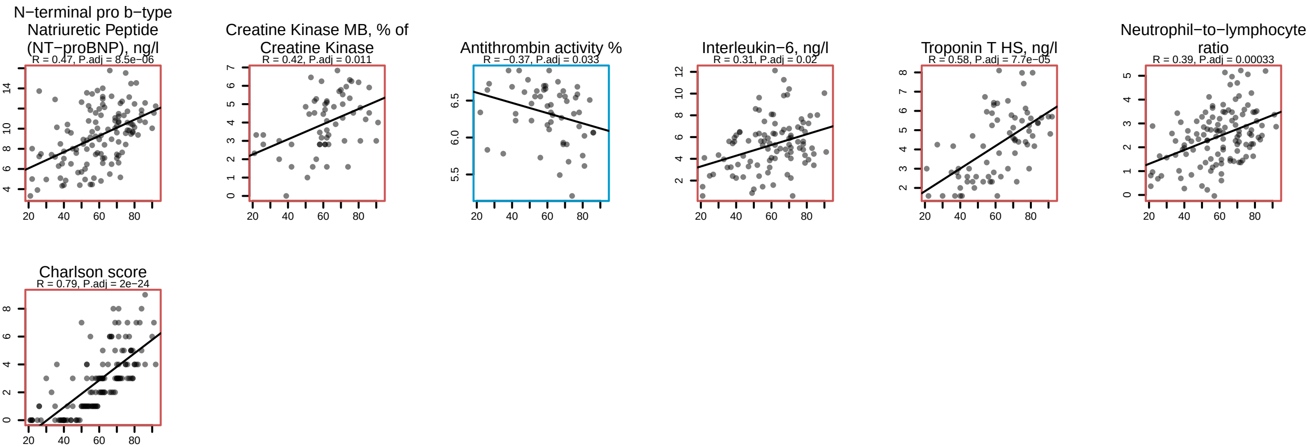

### SF9 - plasma protein levels vs age in general population based on Generation Scotland cohort.pdf

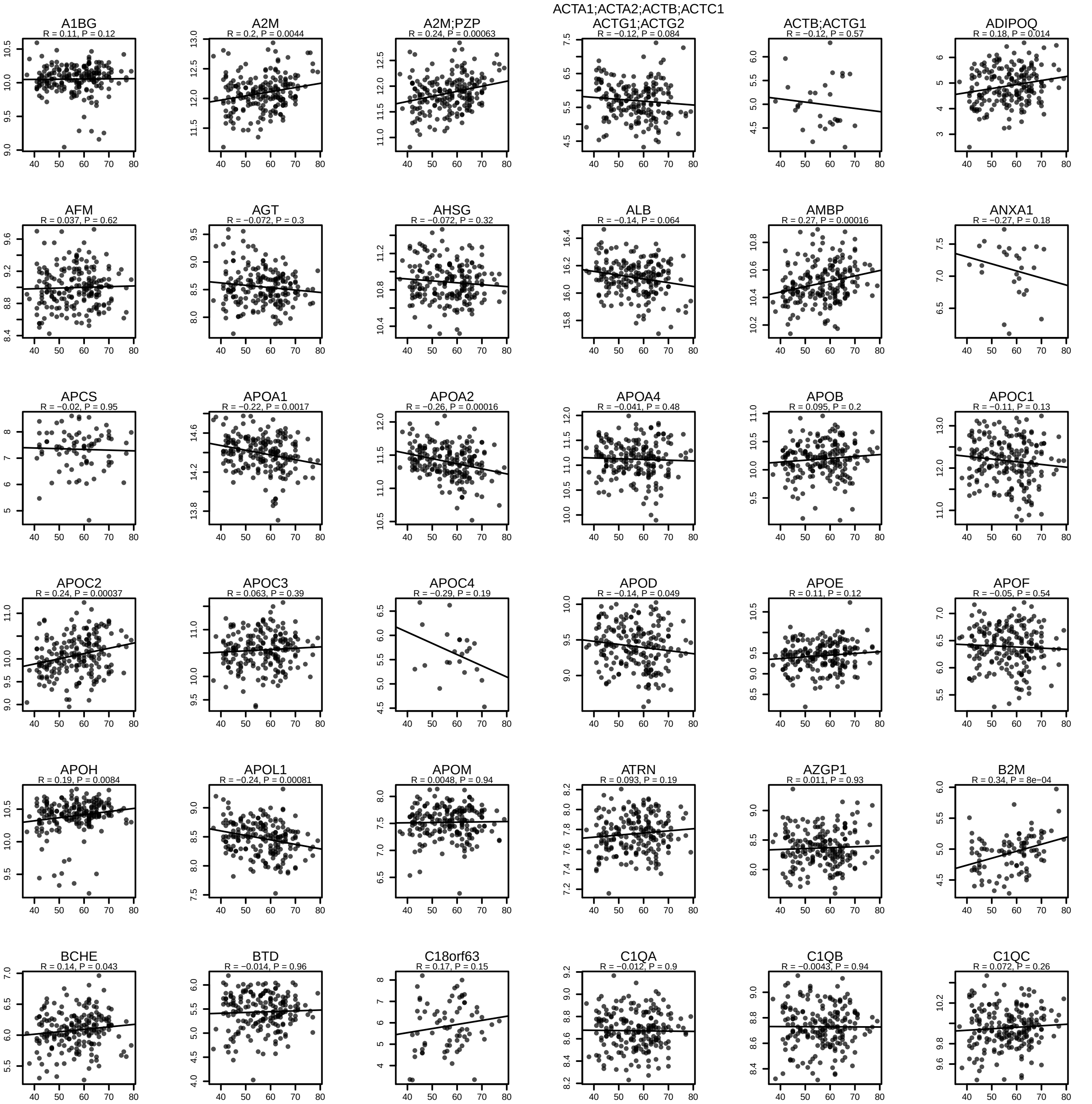

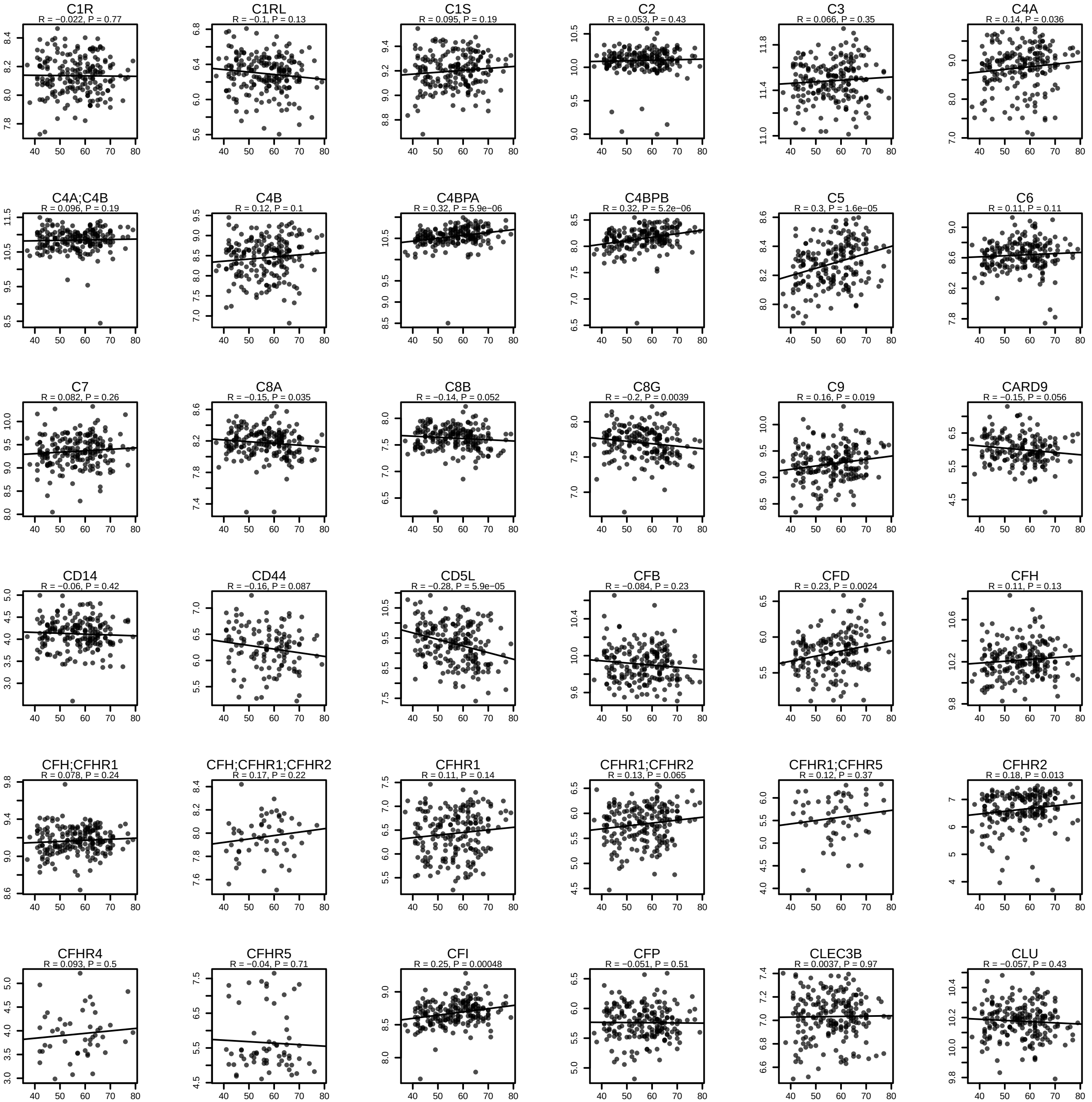

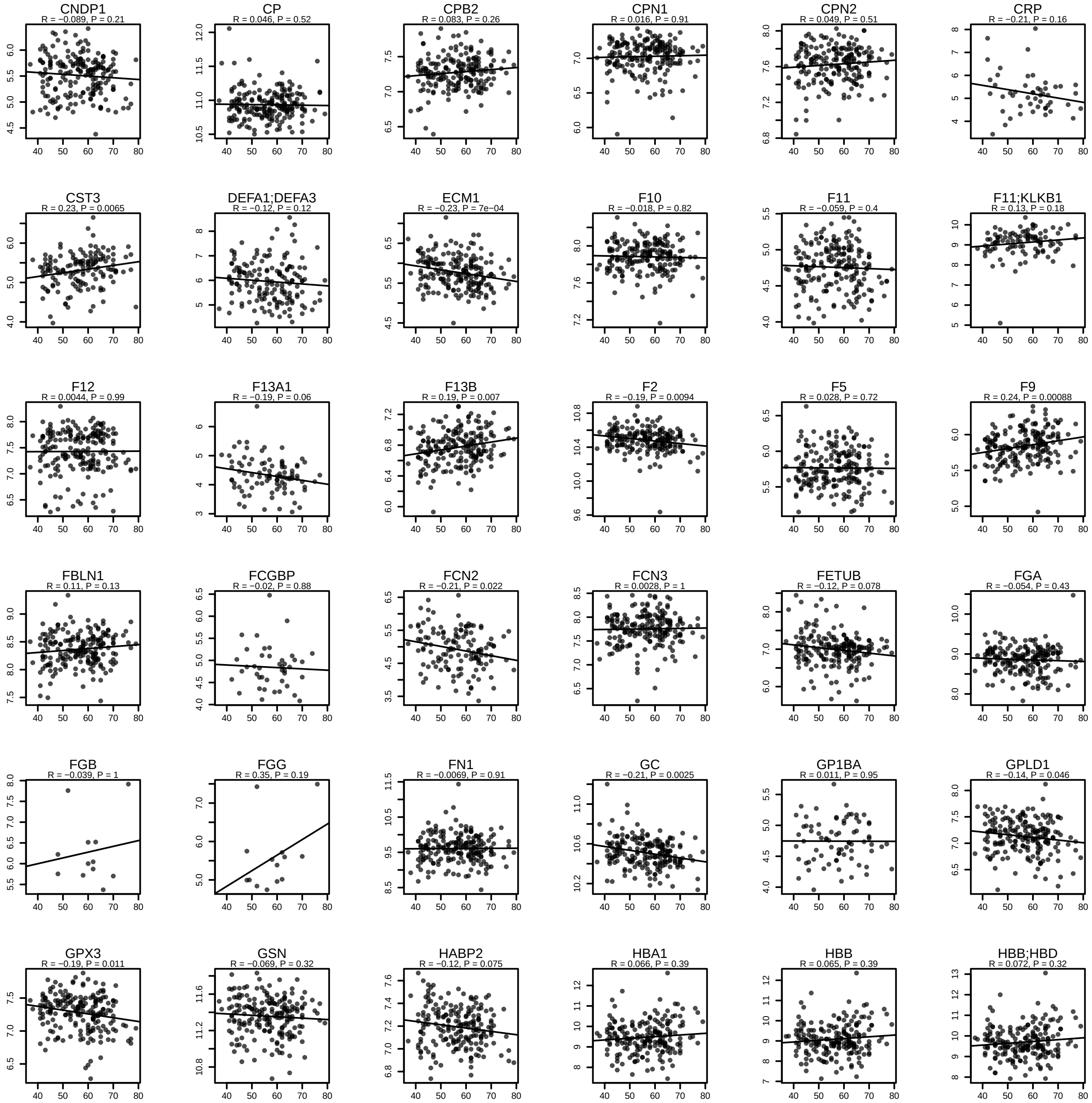

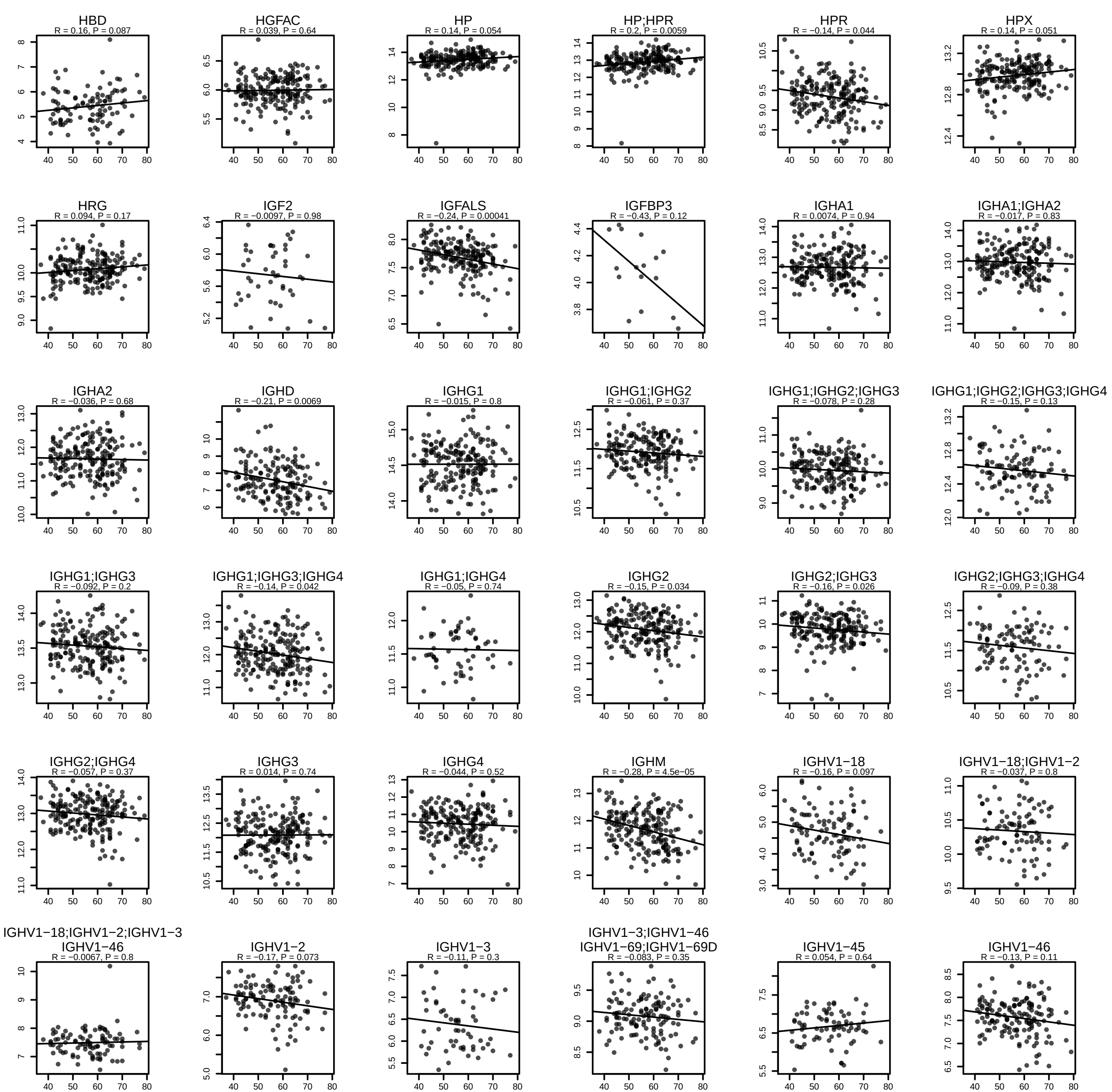

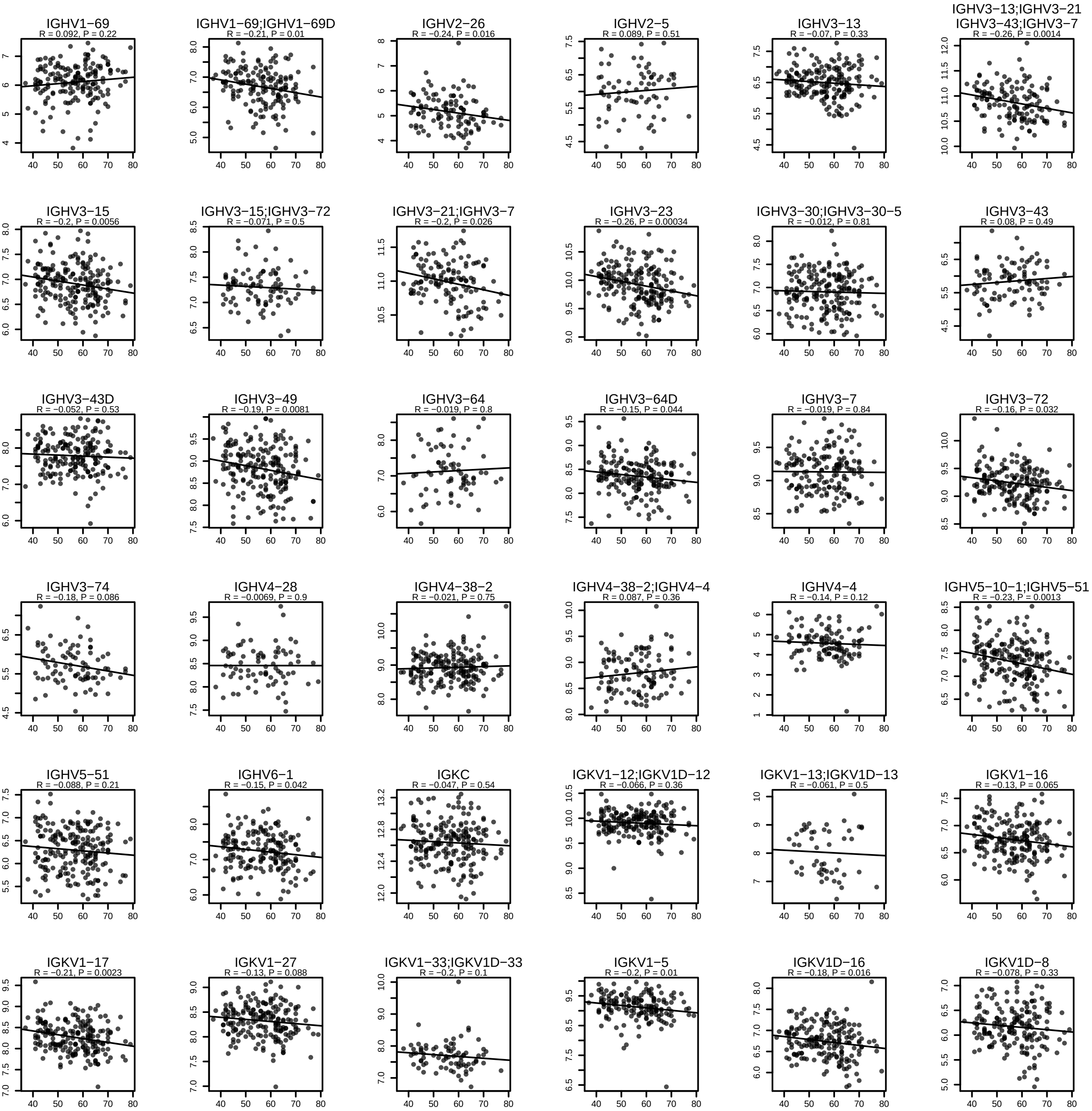

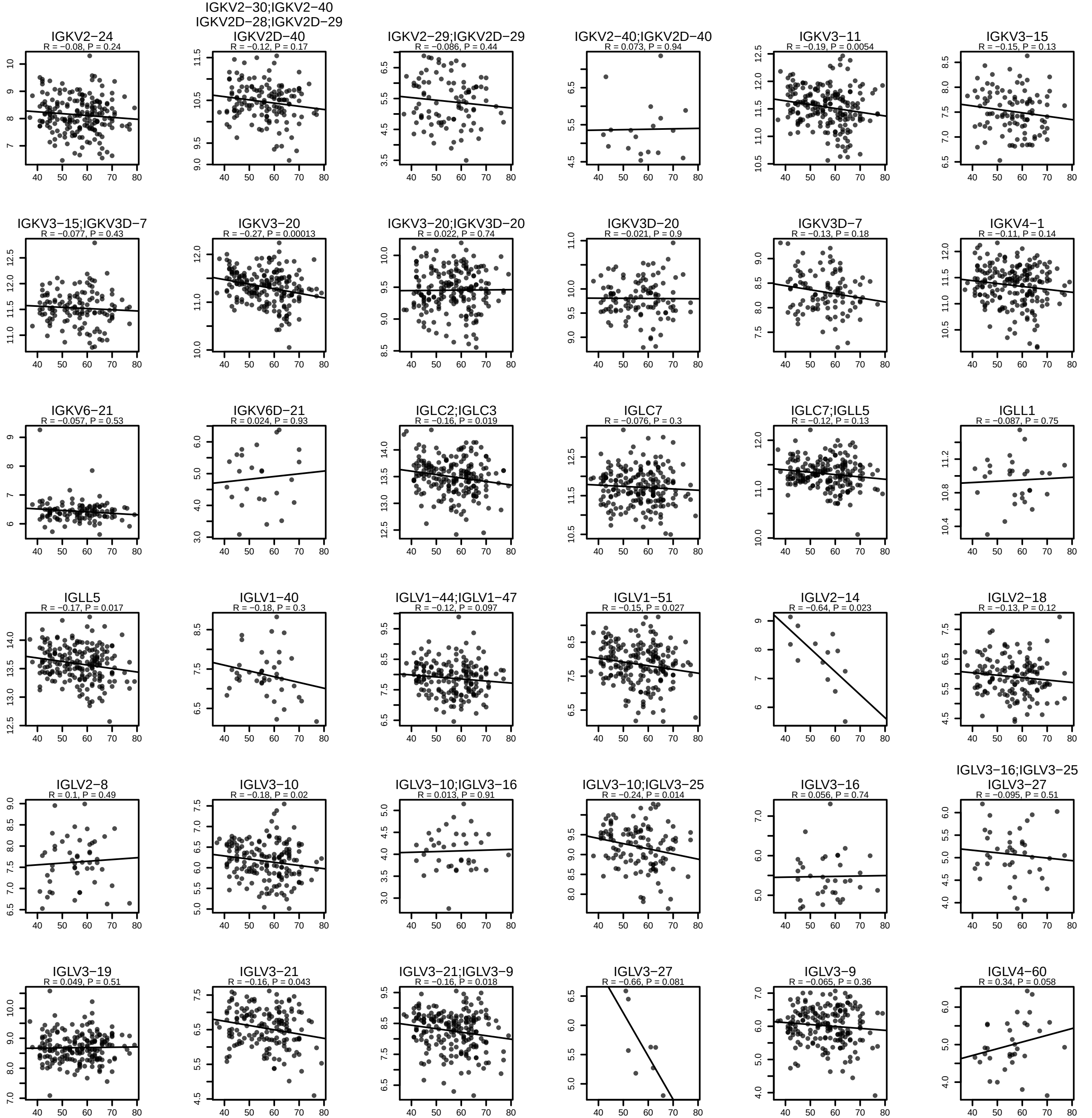

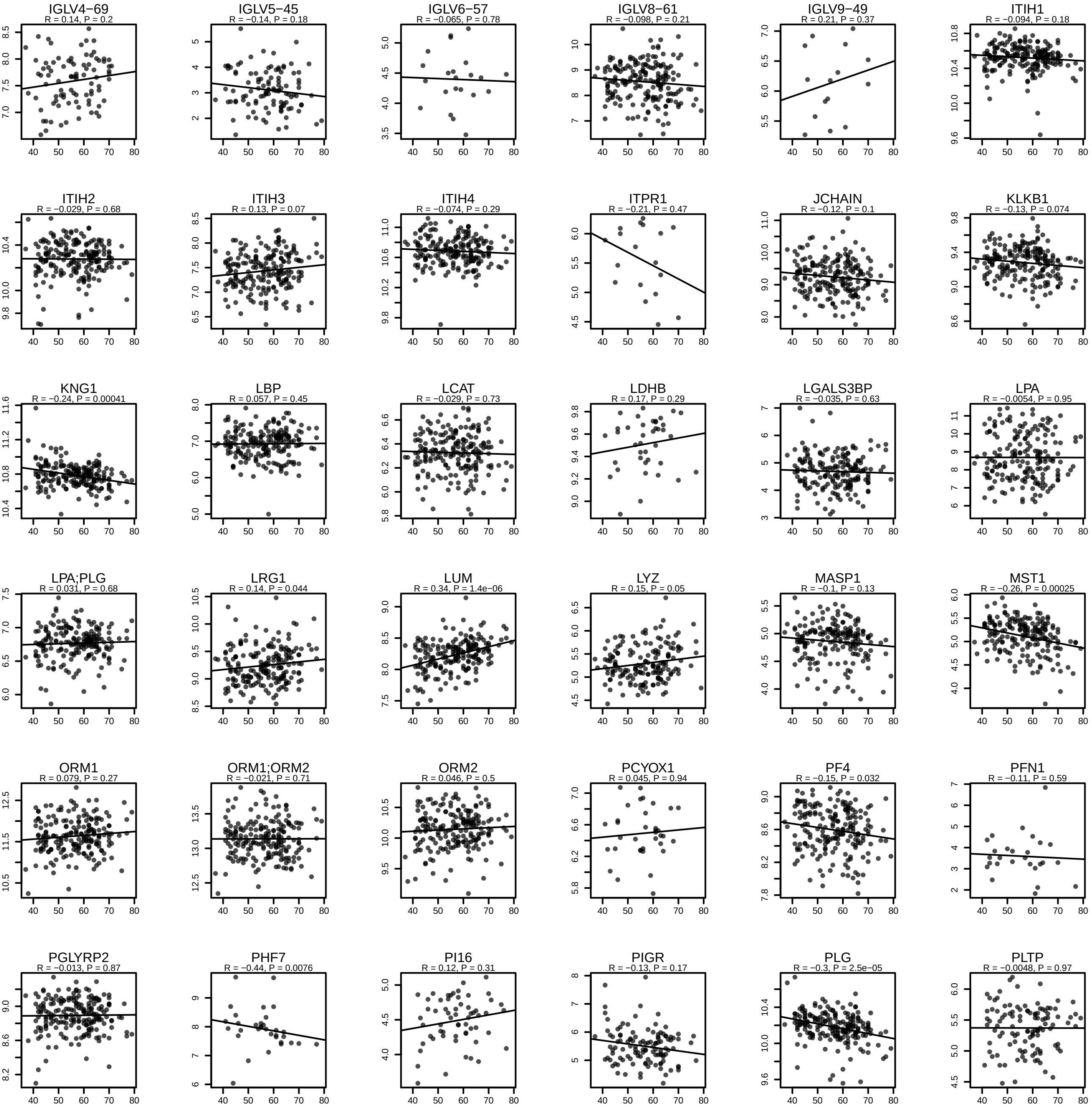

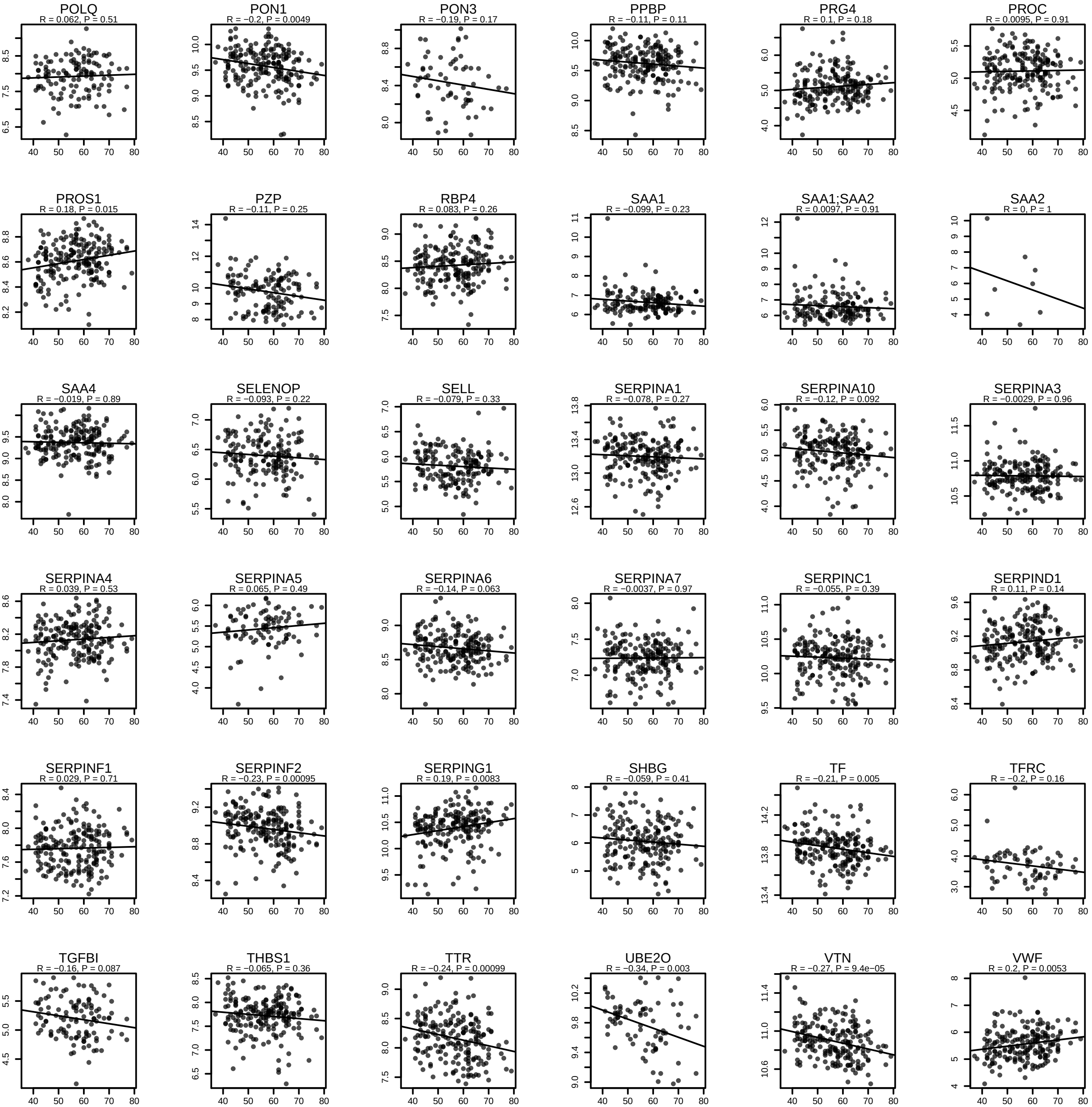

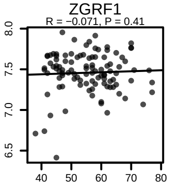

### SF10 - omics features significantly regulated depending on age.pdf

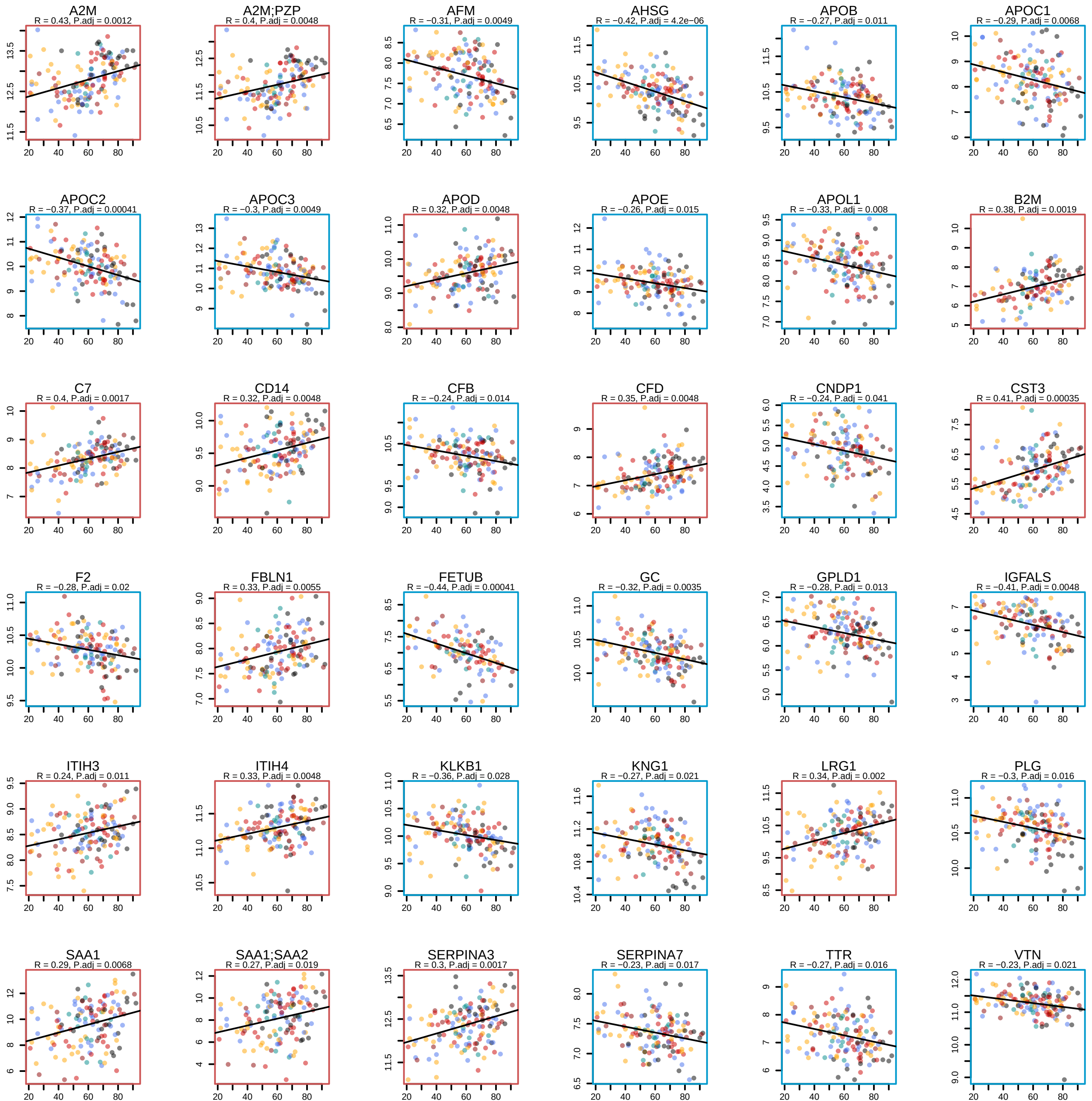

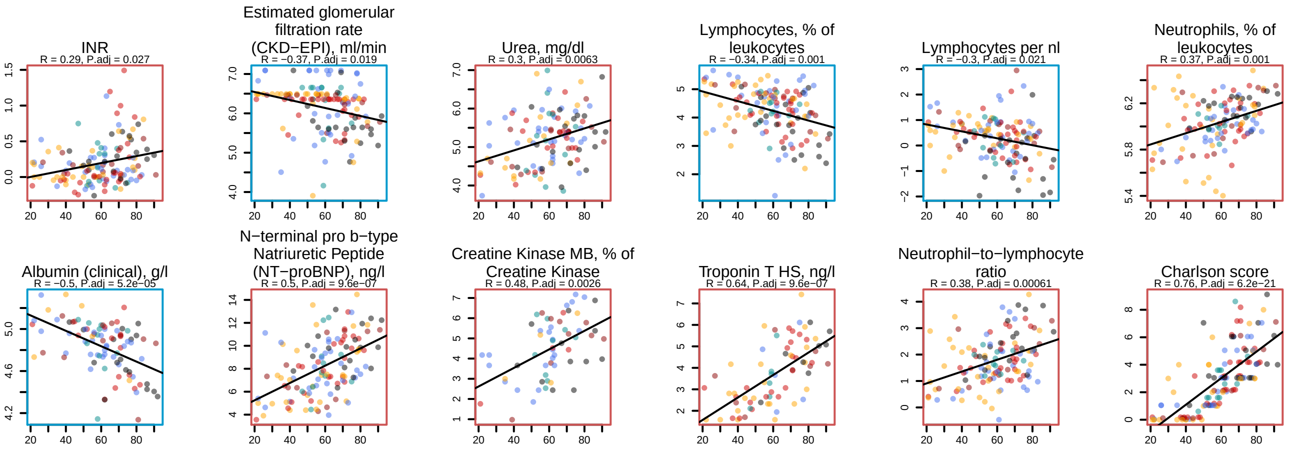

### SF11 - map of correlations between dynamic changes in omics measurements during the peak period of the disease.pdf

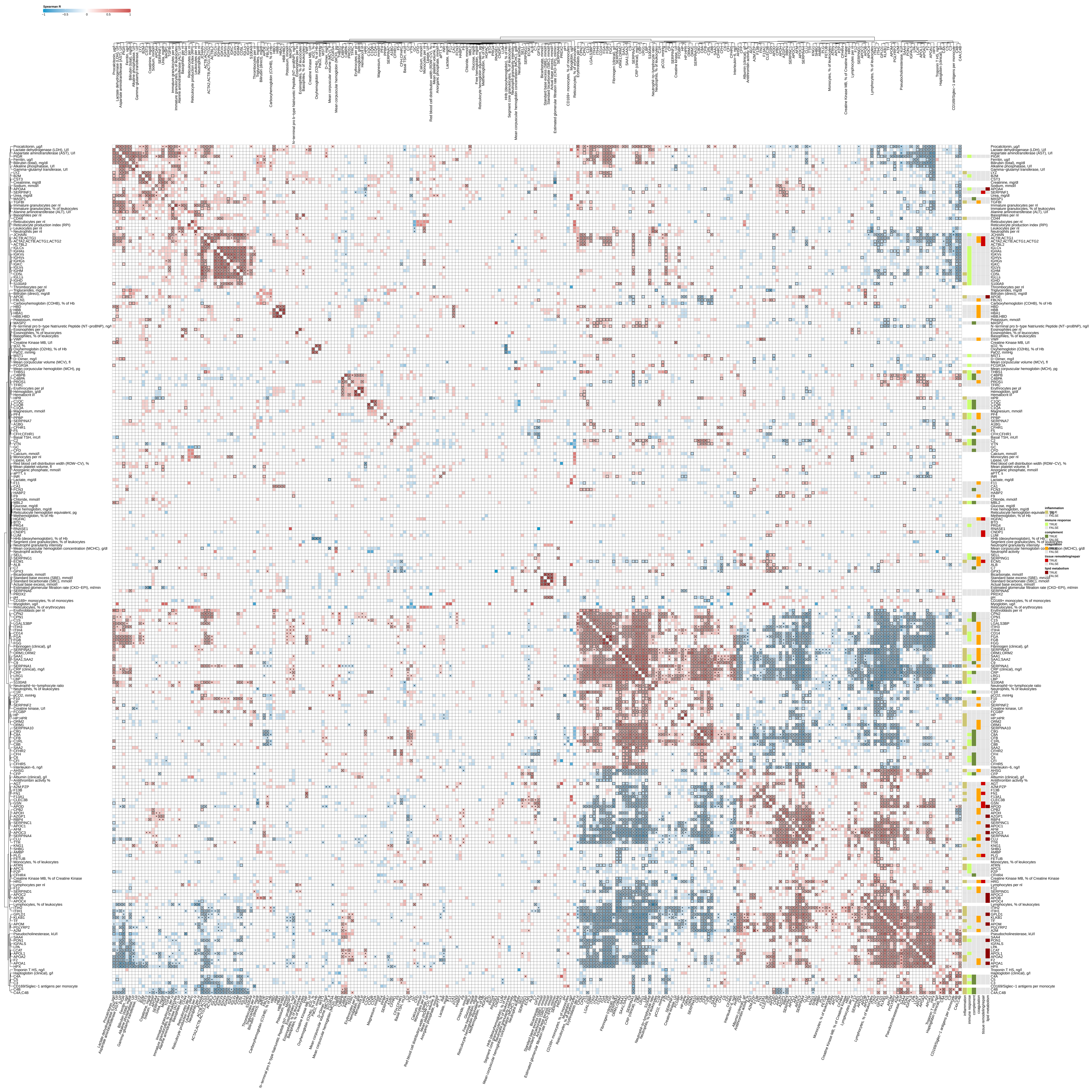

### SF12 - covariation of organ function markers and plasma proteins.pdf

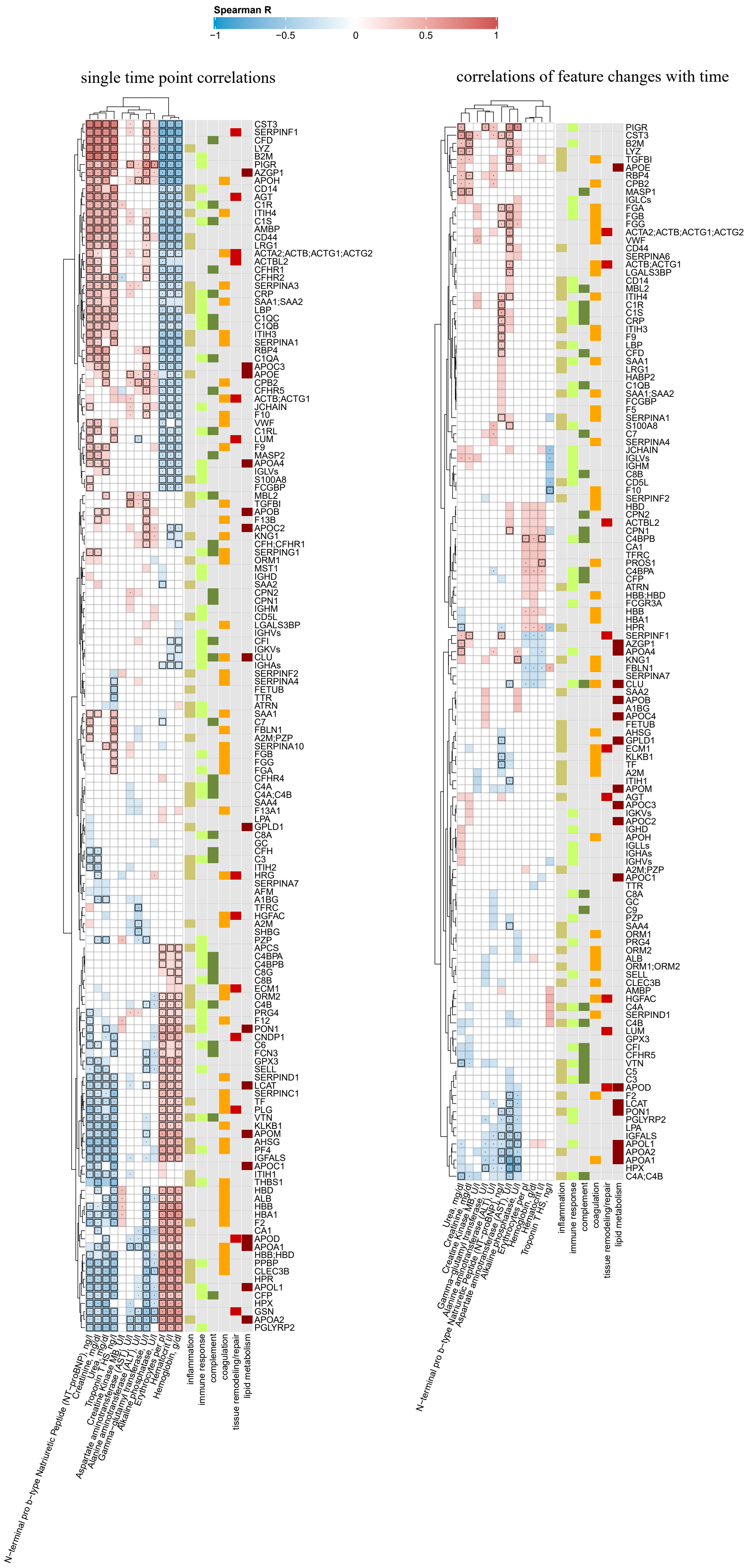

### SF13 - omics features changing during the peak of the disease - log2 fold change.pdf

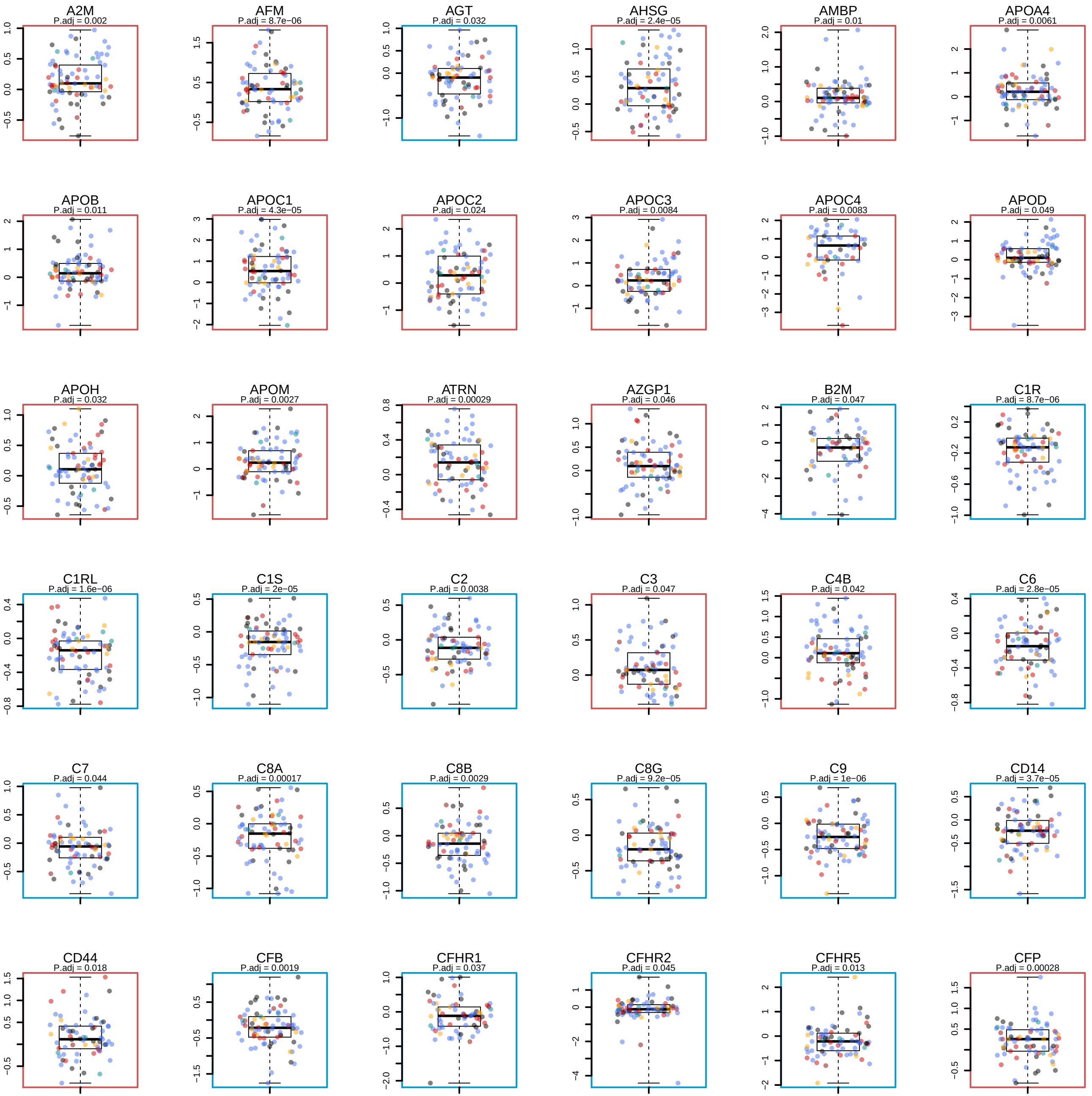

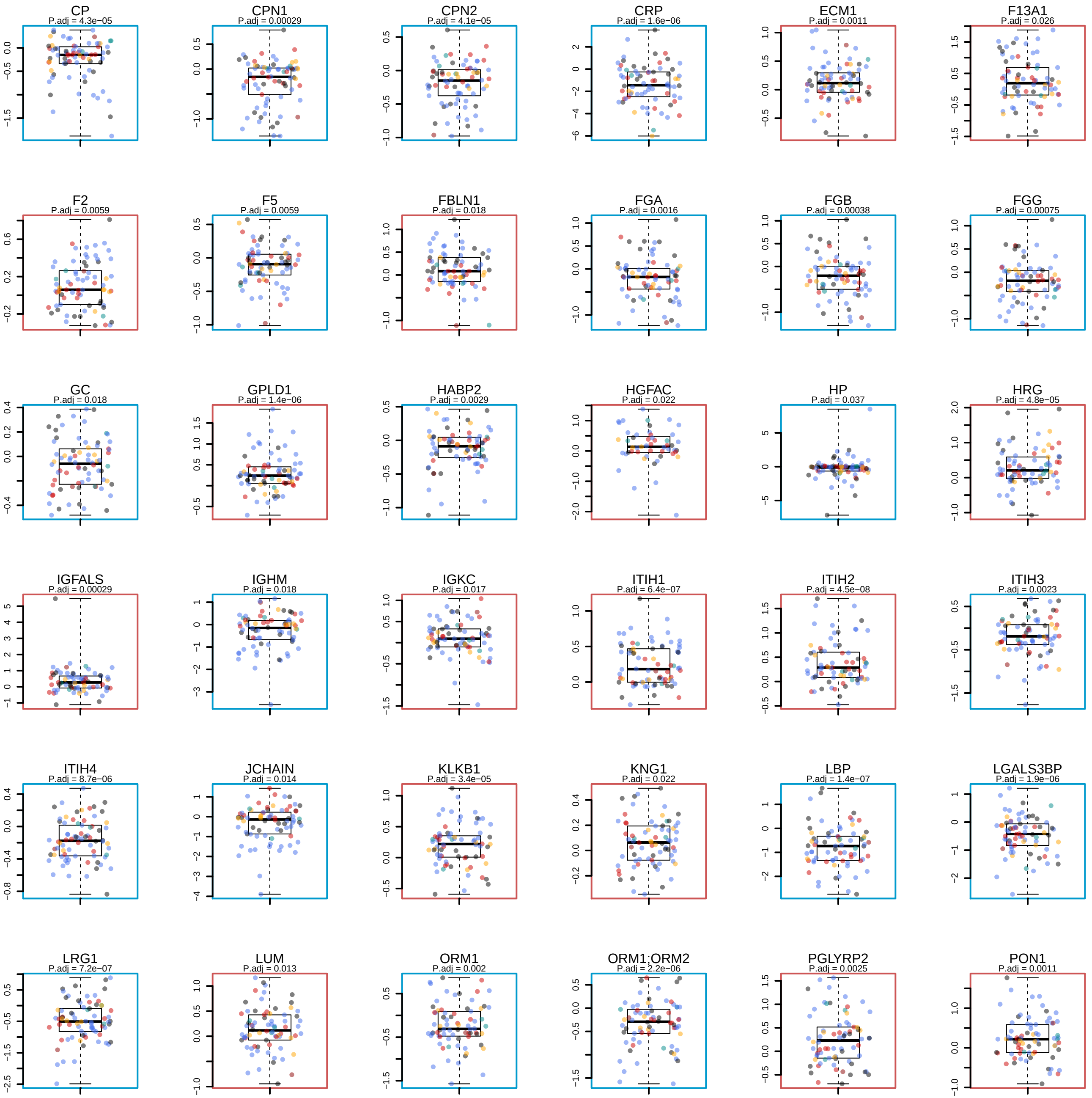

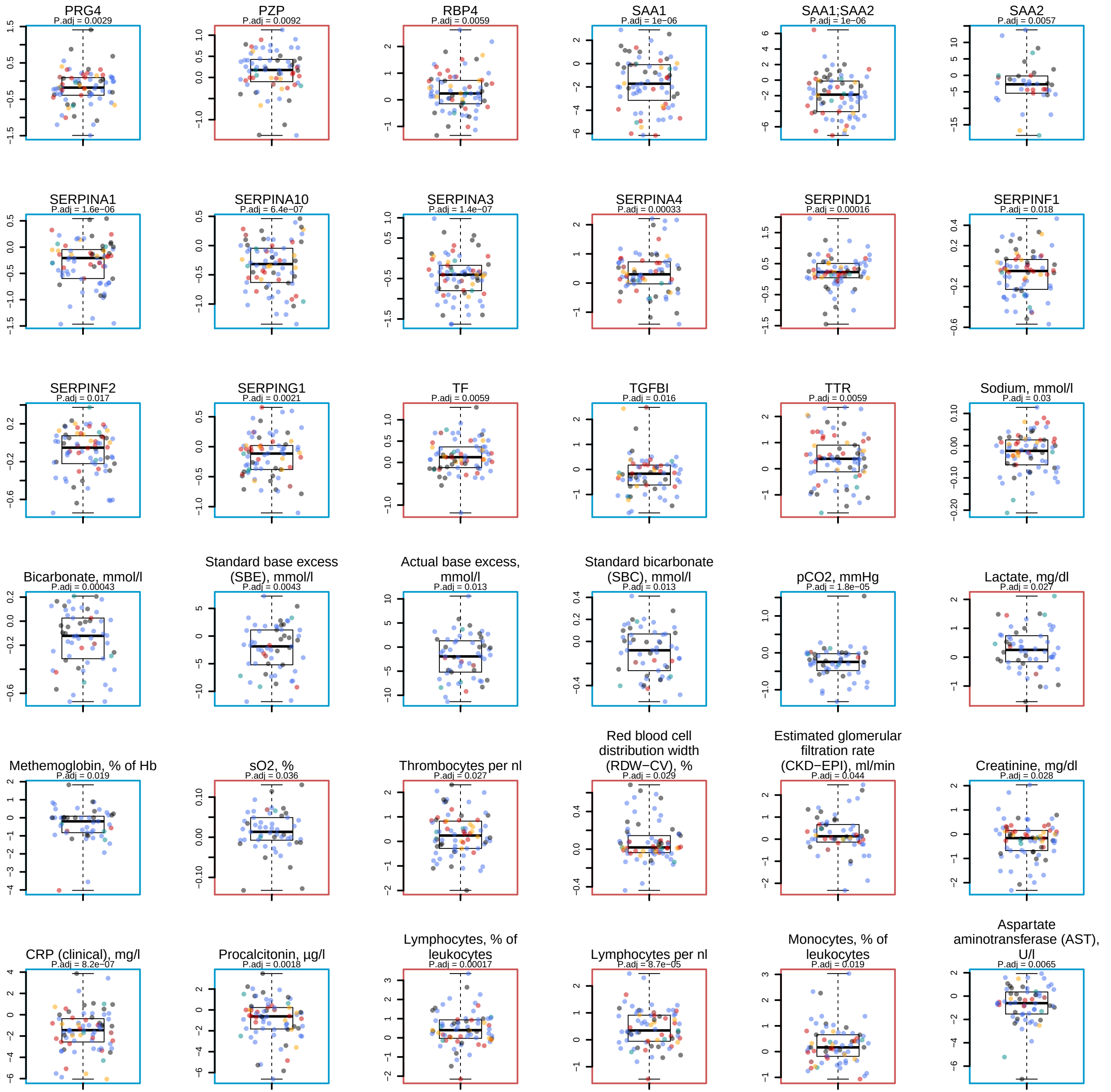

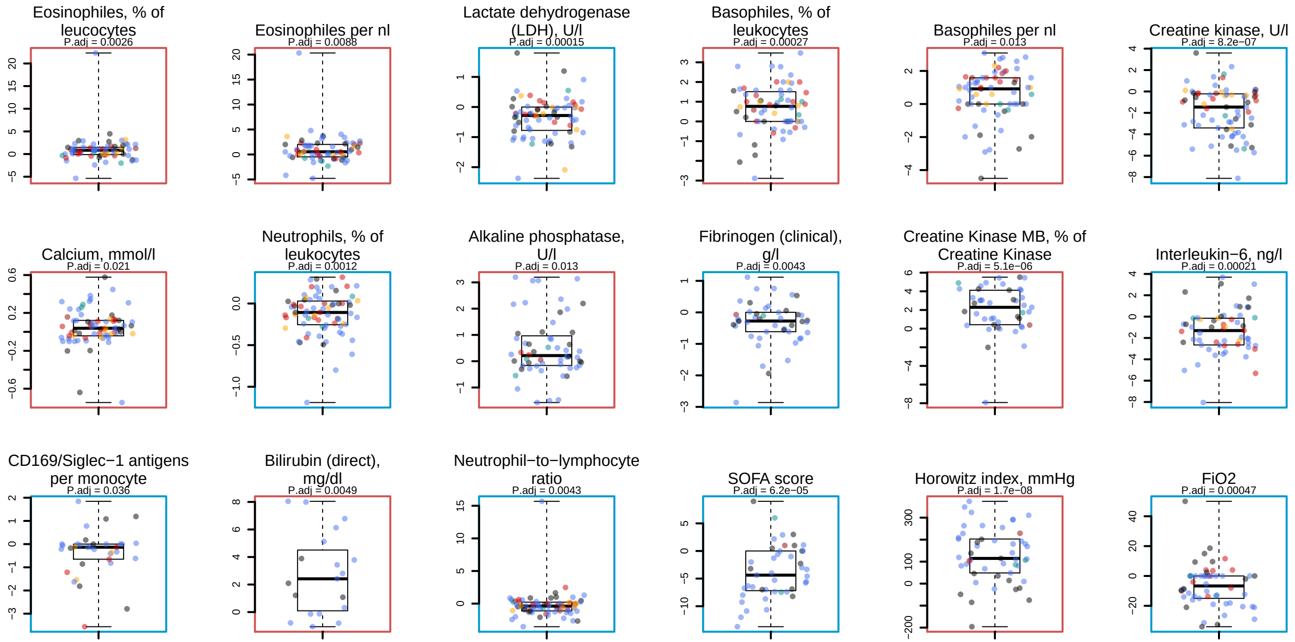

### SF14 - omics features differentially changing during the peak of the disease depending on the severity.pdf

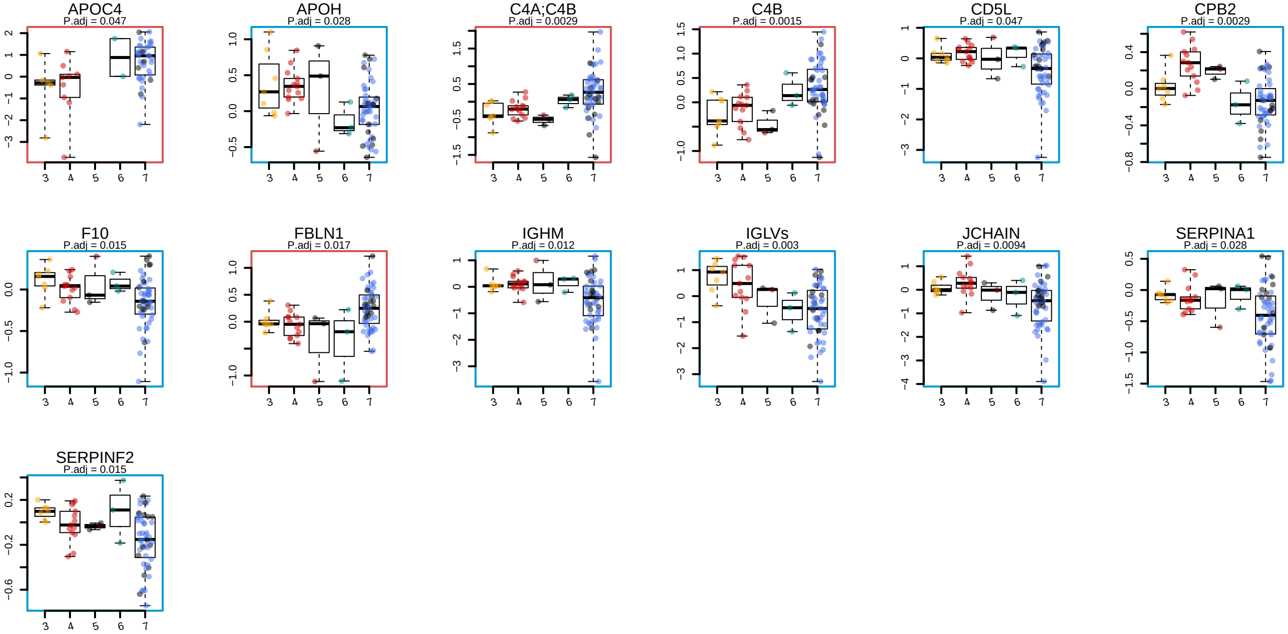

### SF15 - trajectories at the disease peak for max WHO = 3.pdf

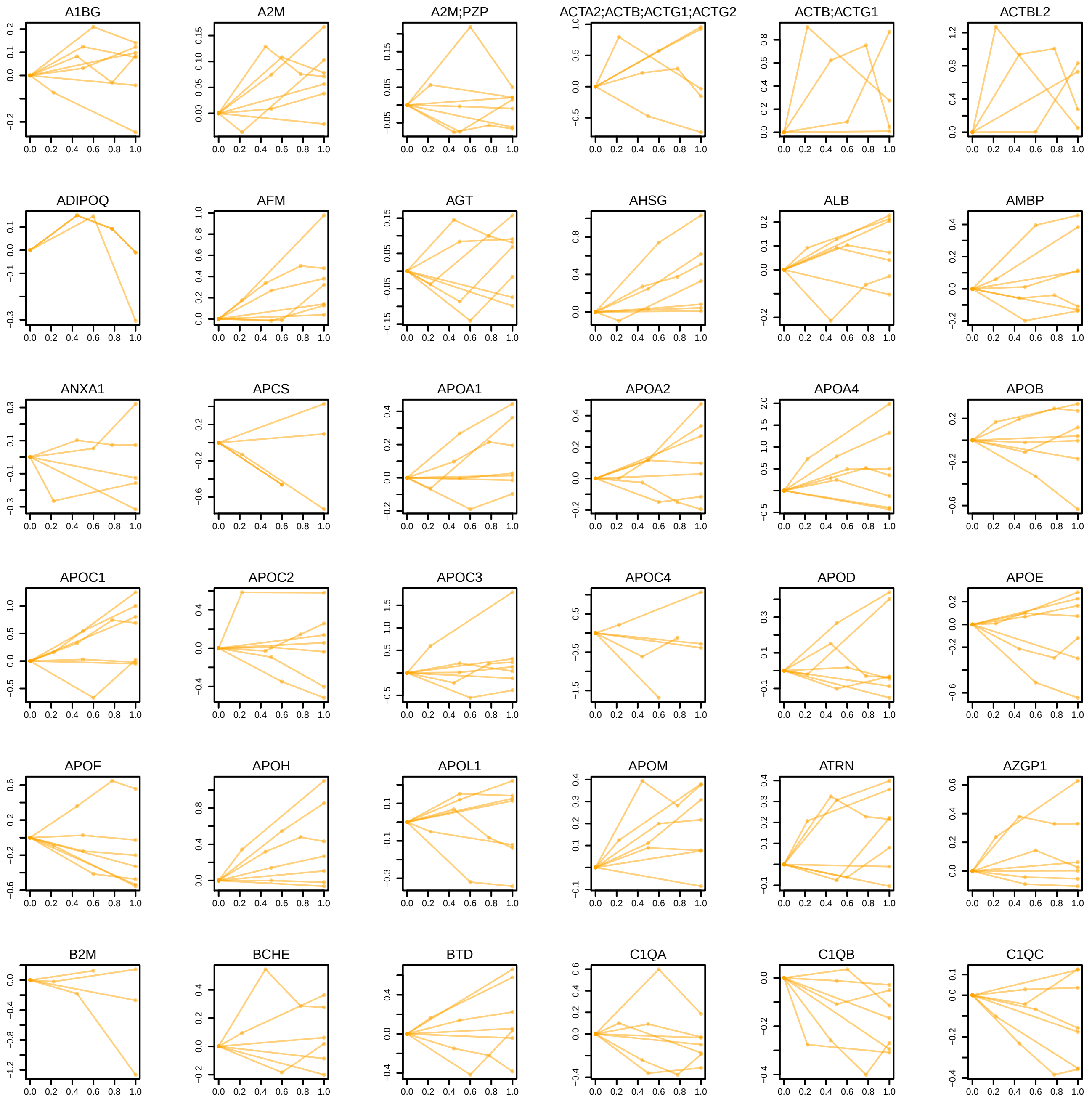

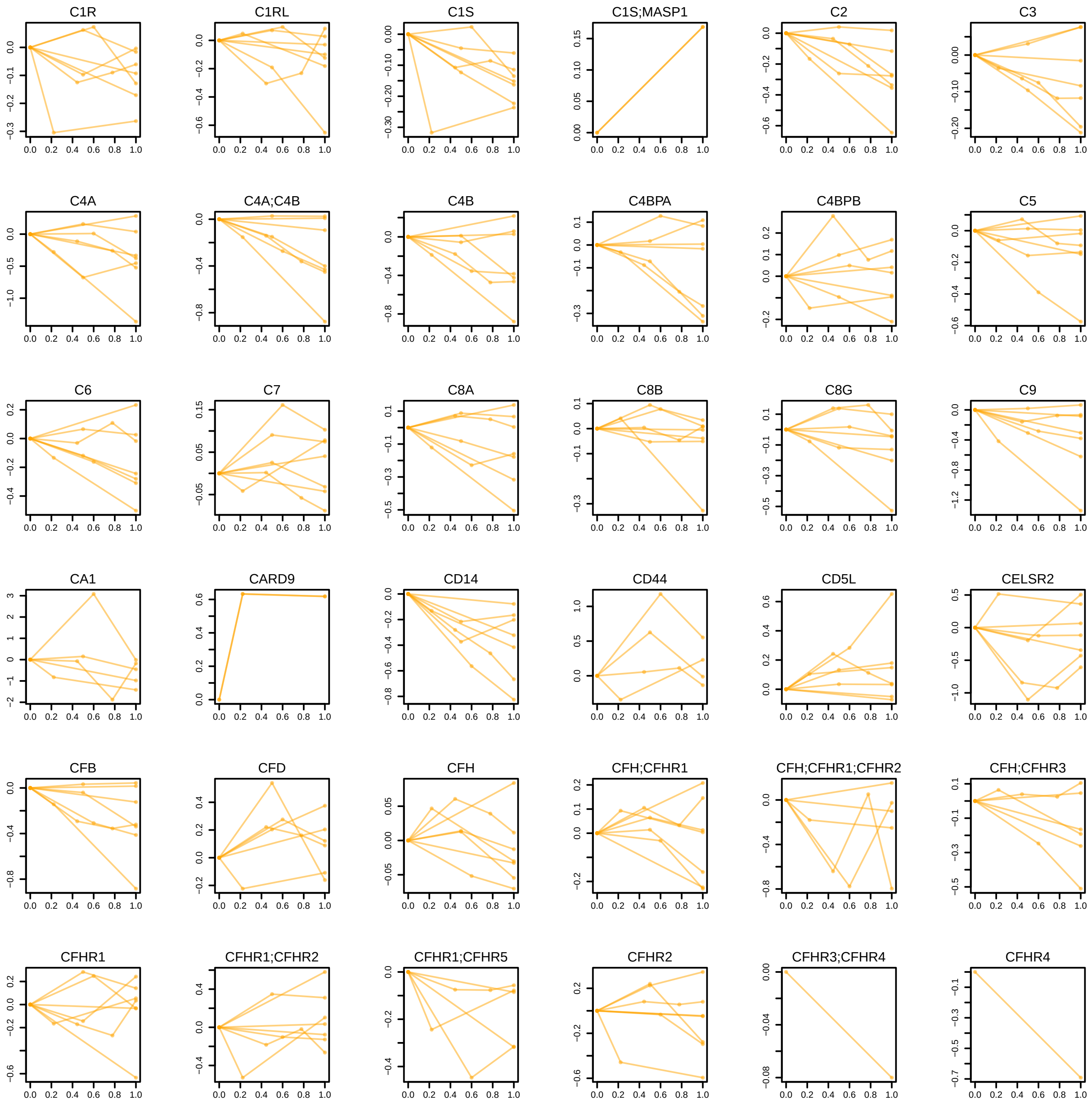

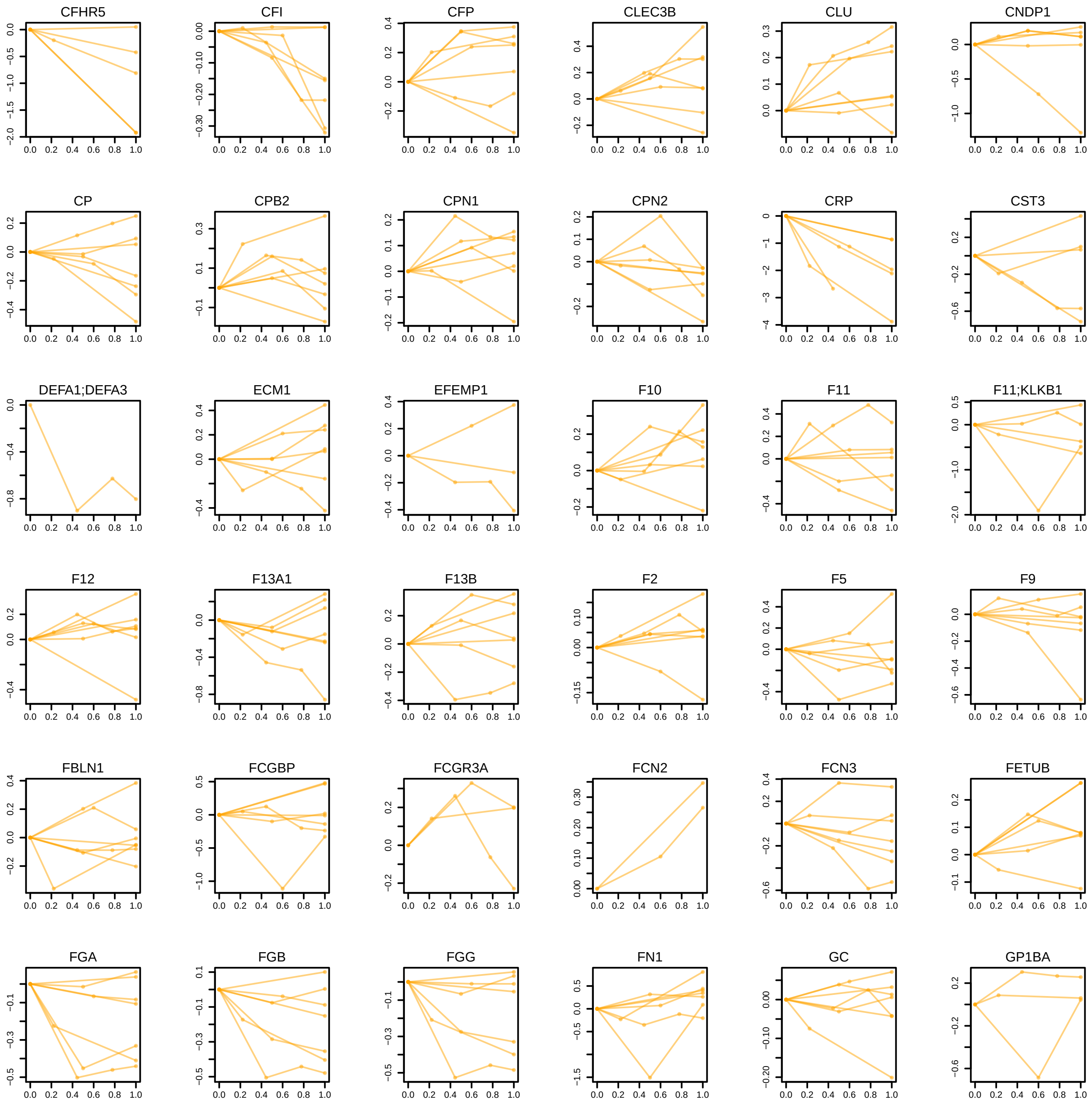

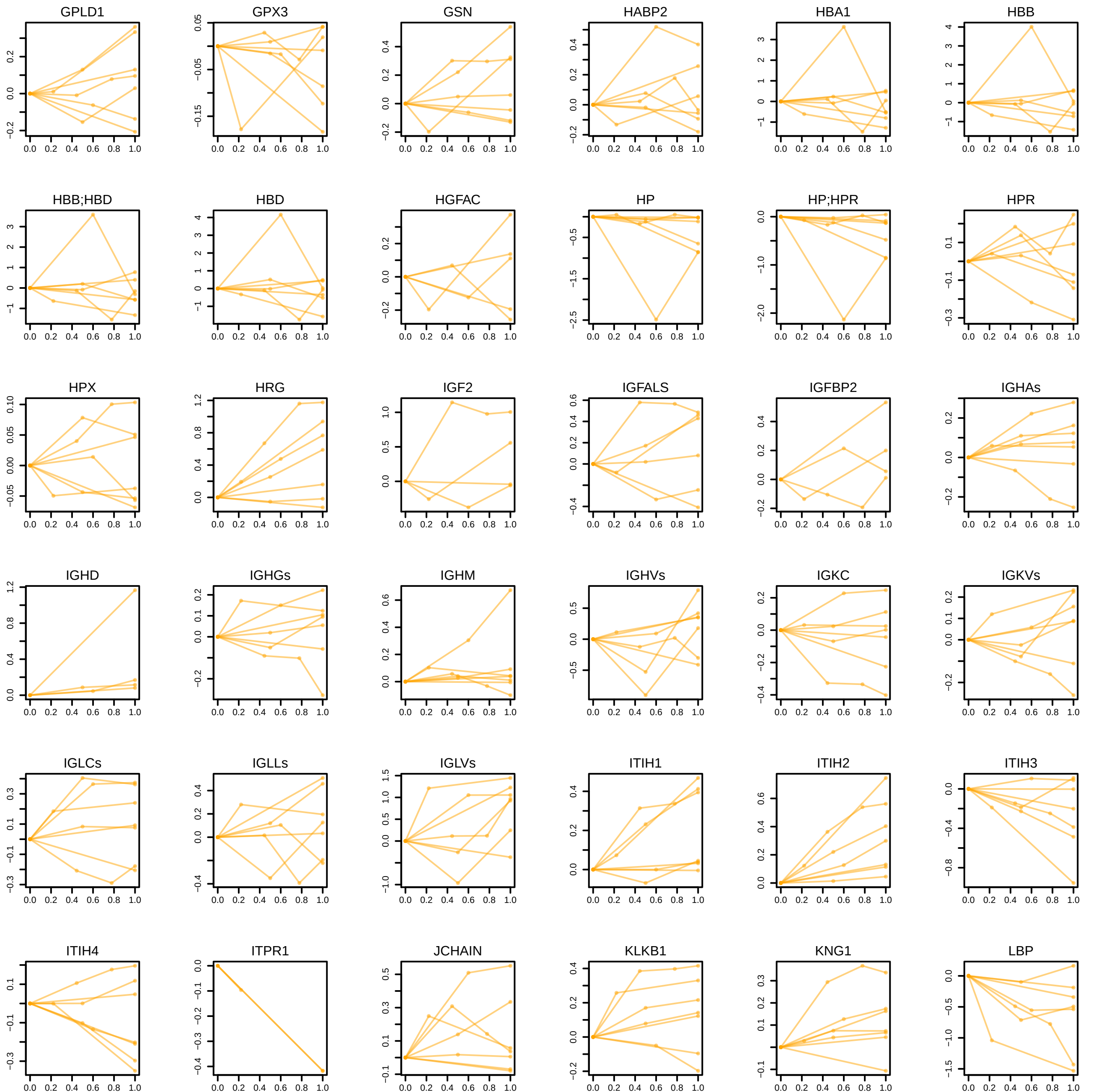

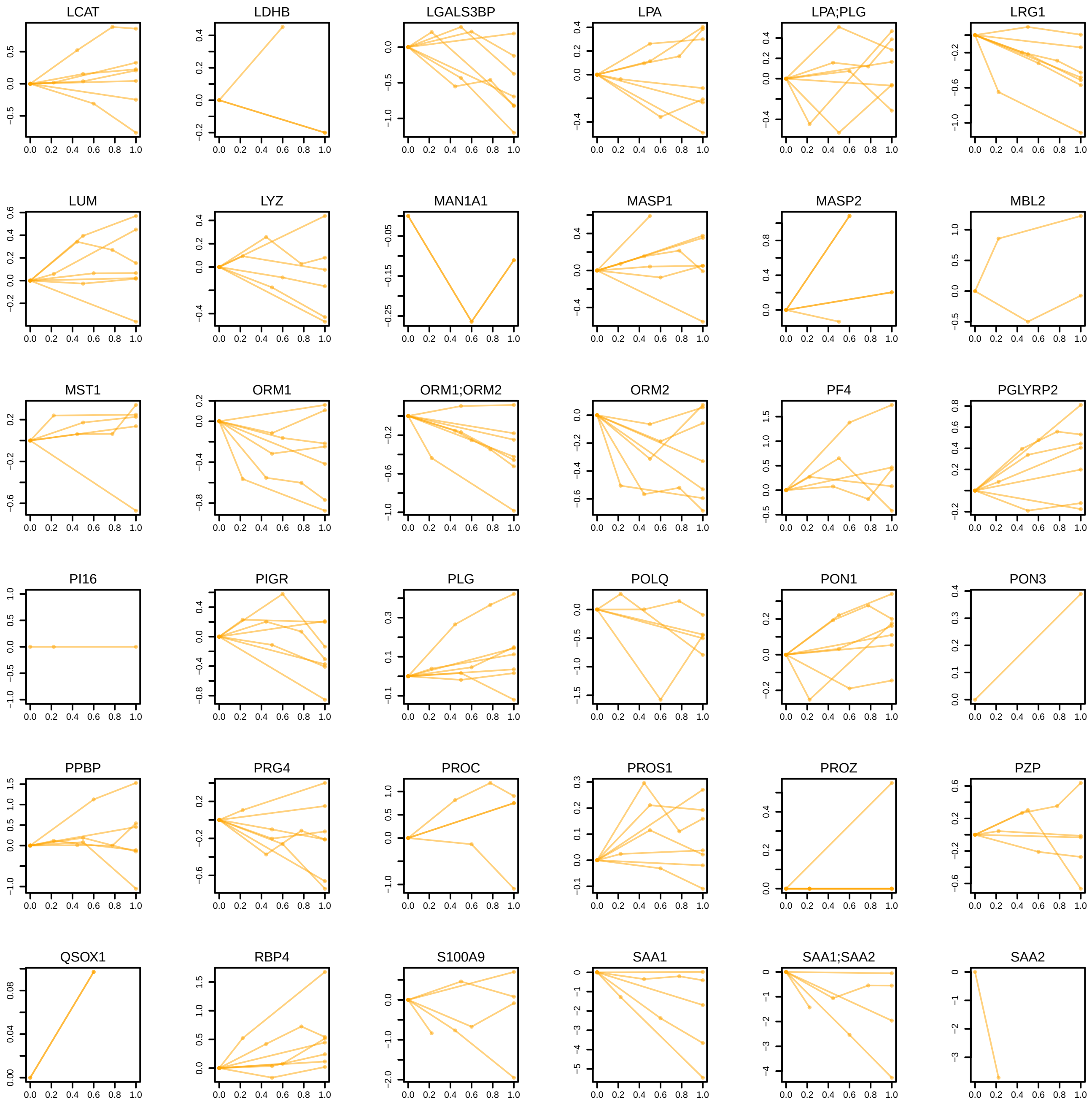

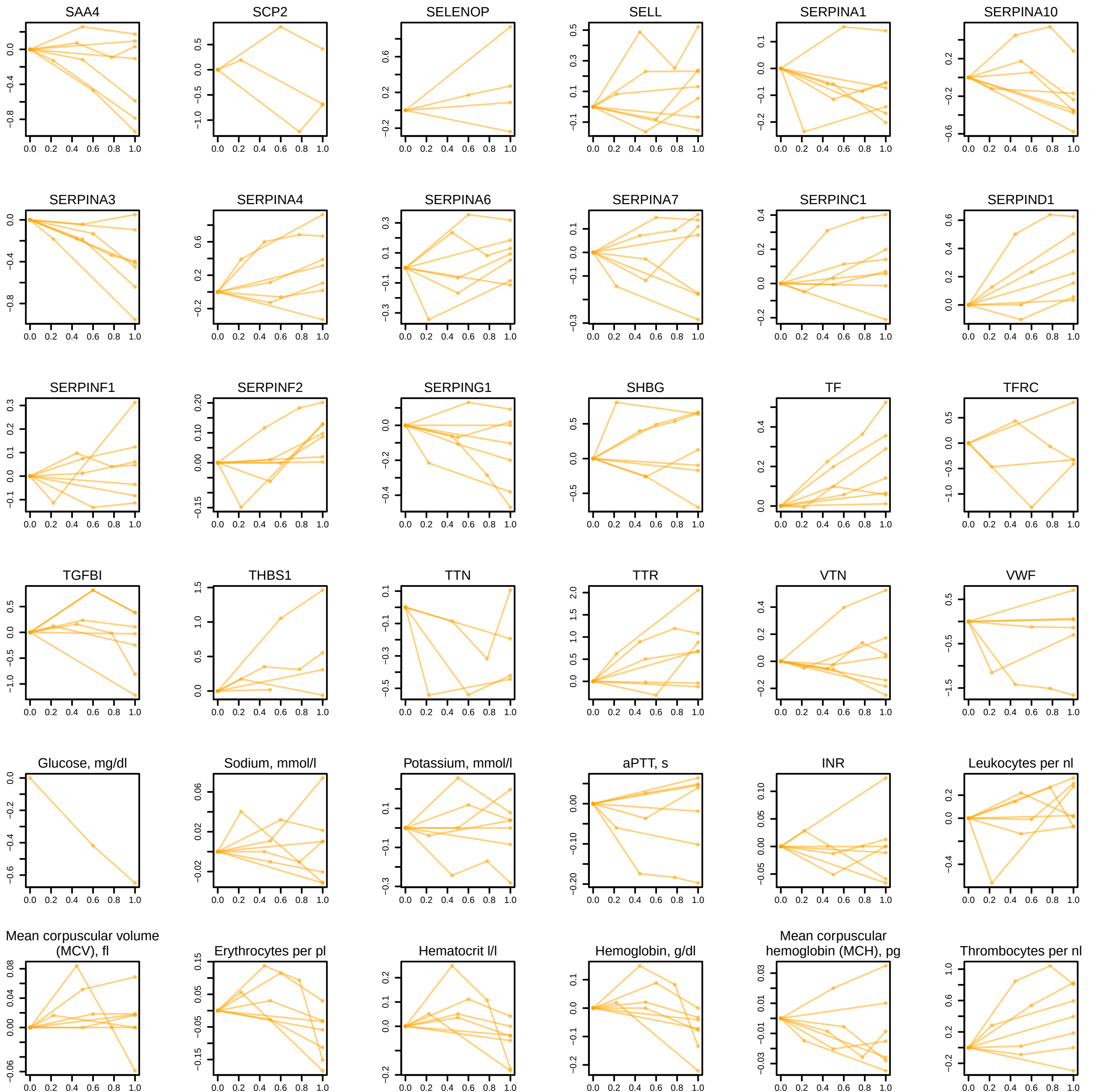

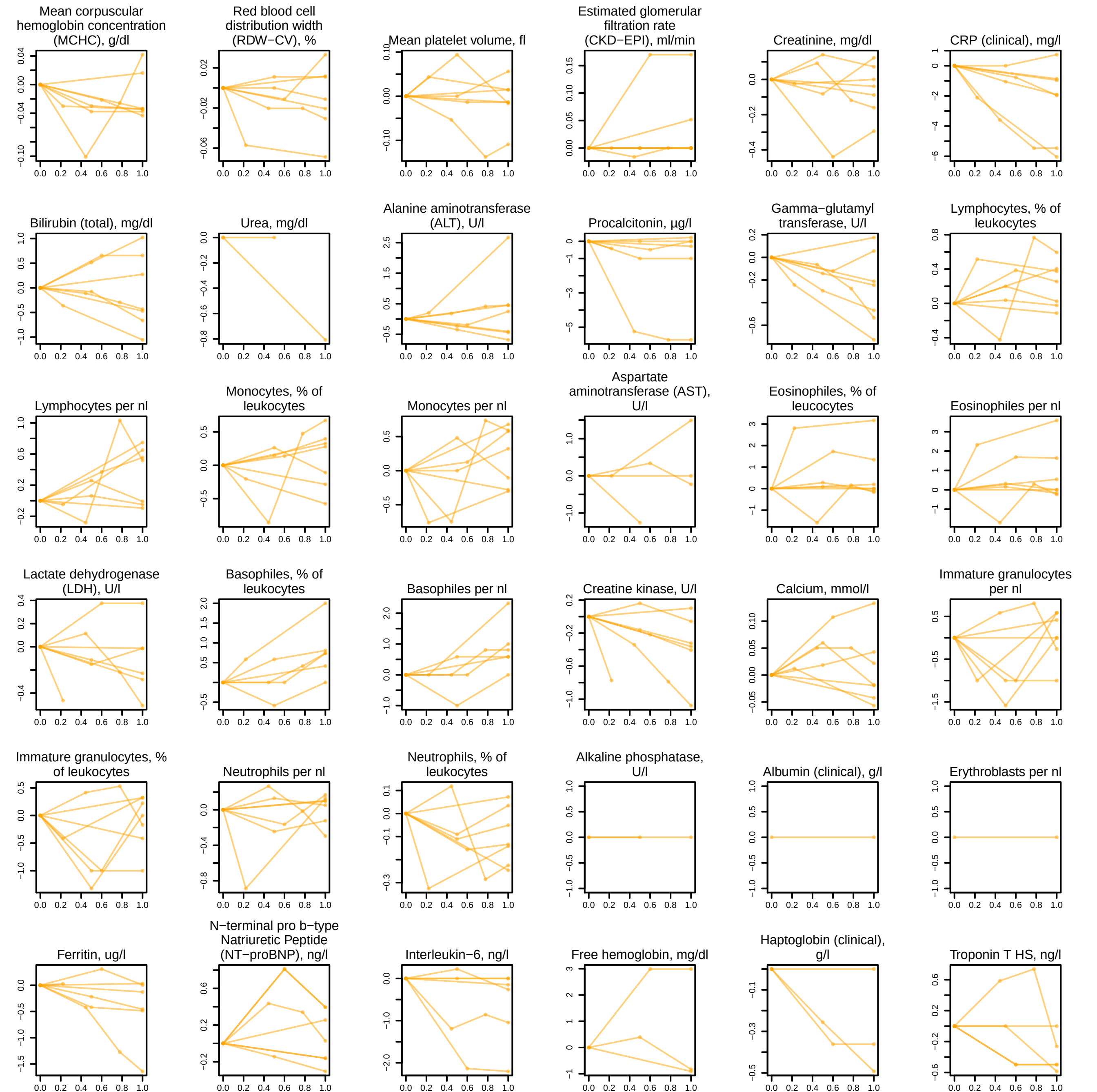

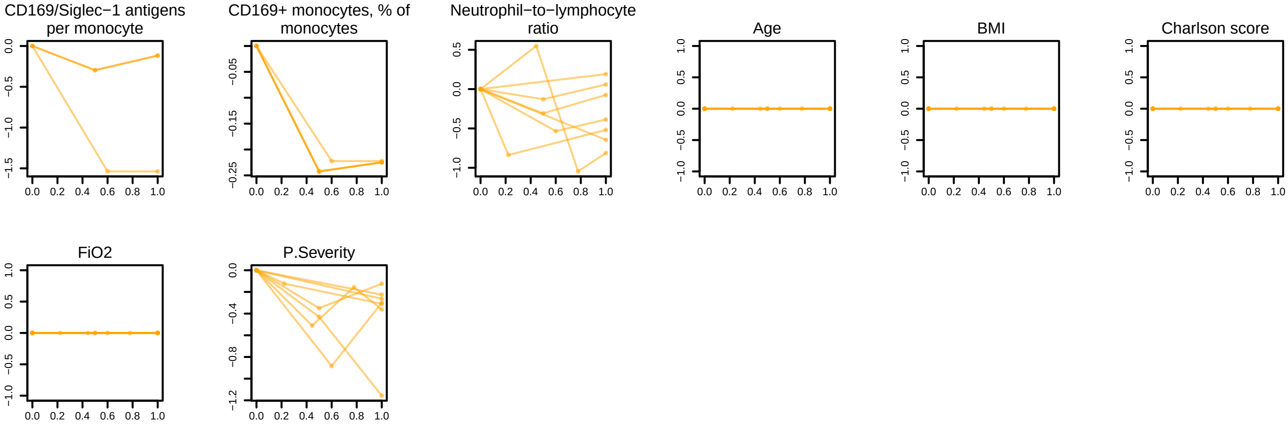

### SF16 - trajectories at the disease peak for max WHO = 4.pdf

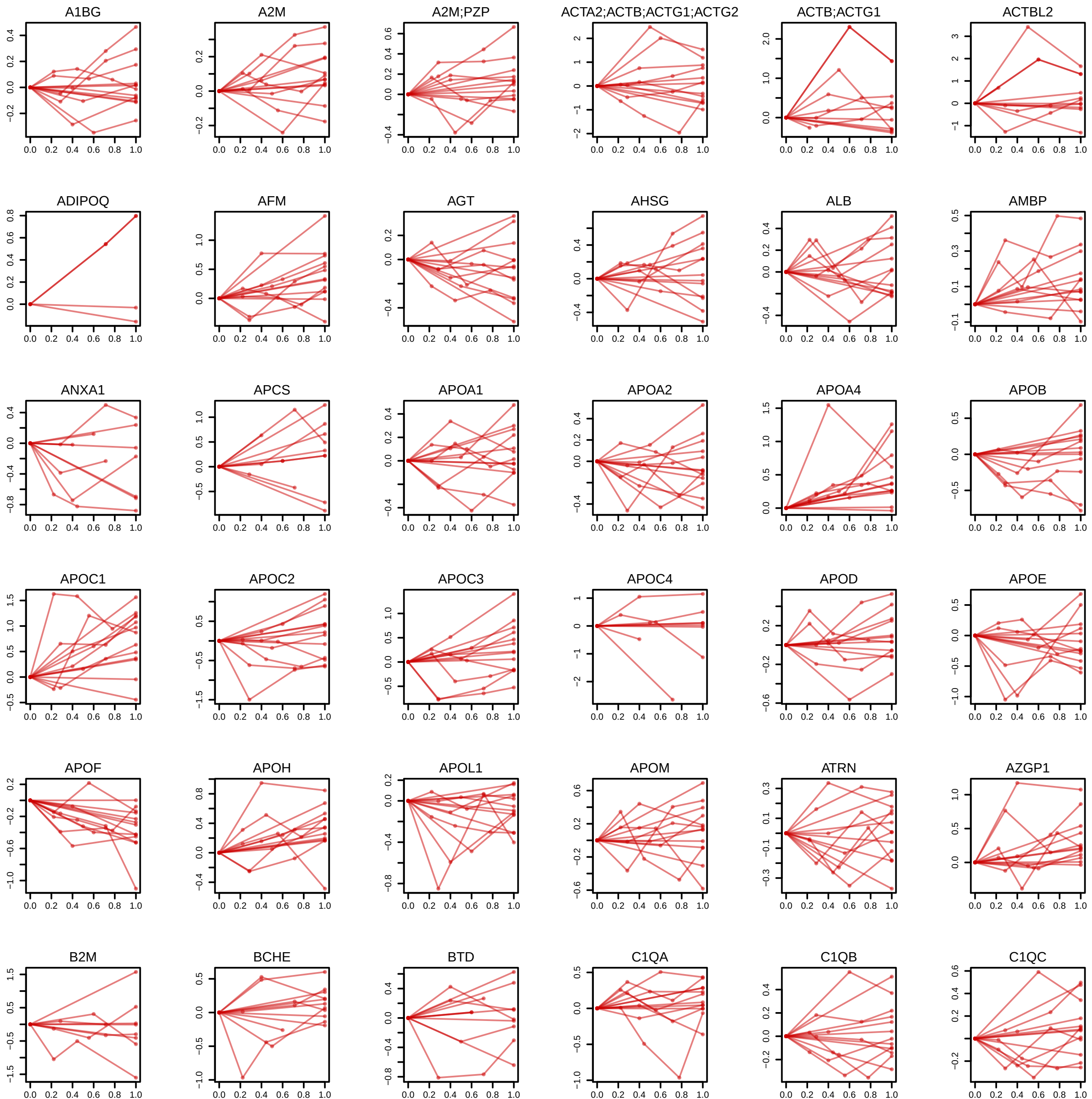

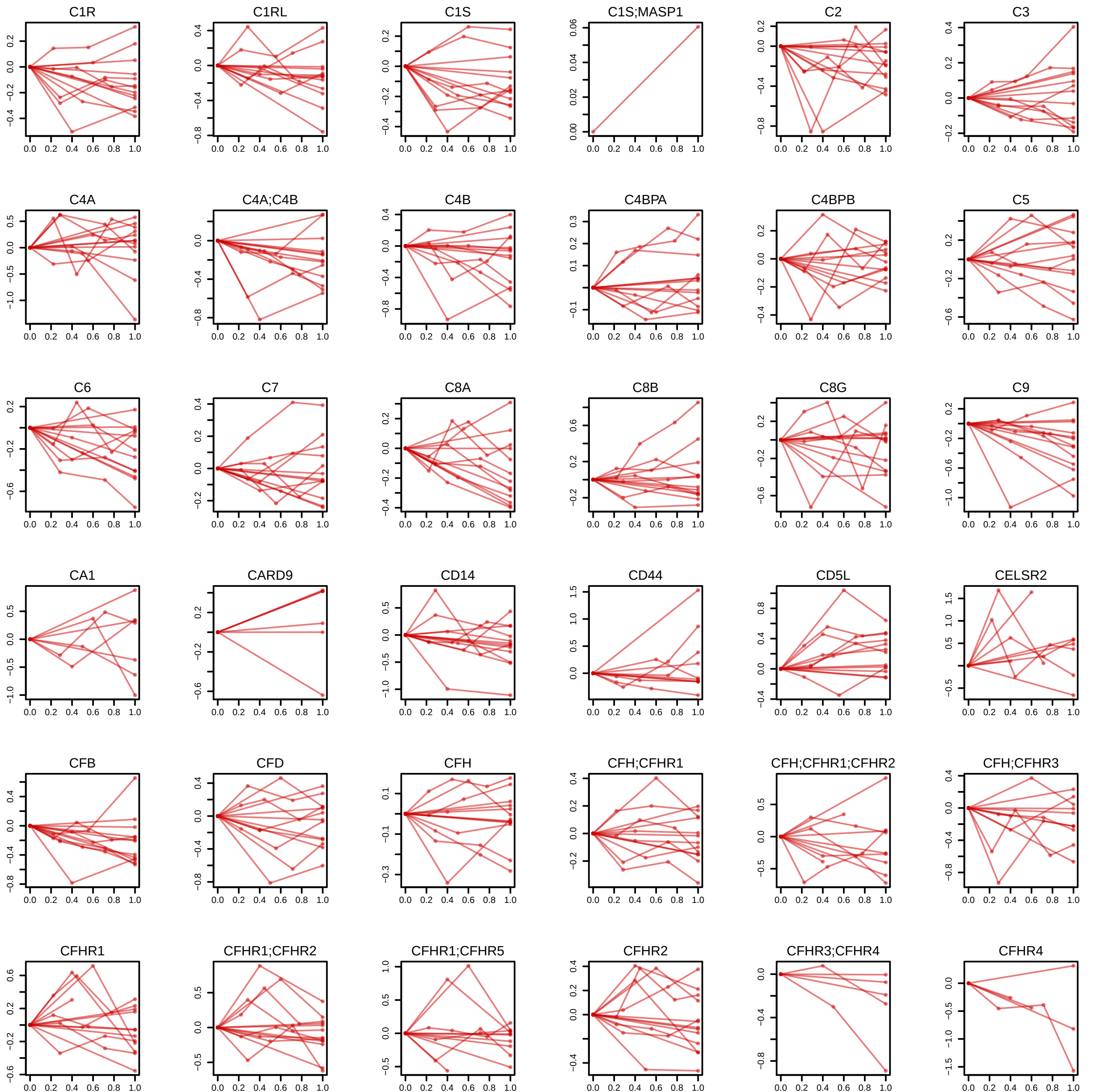

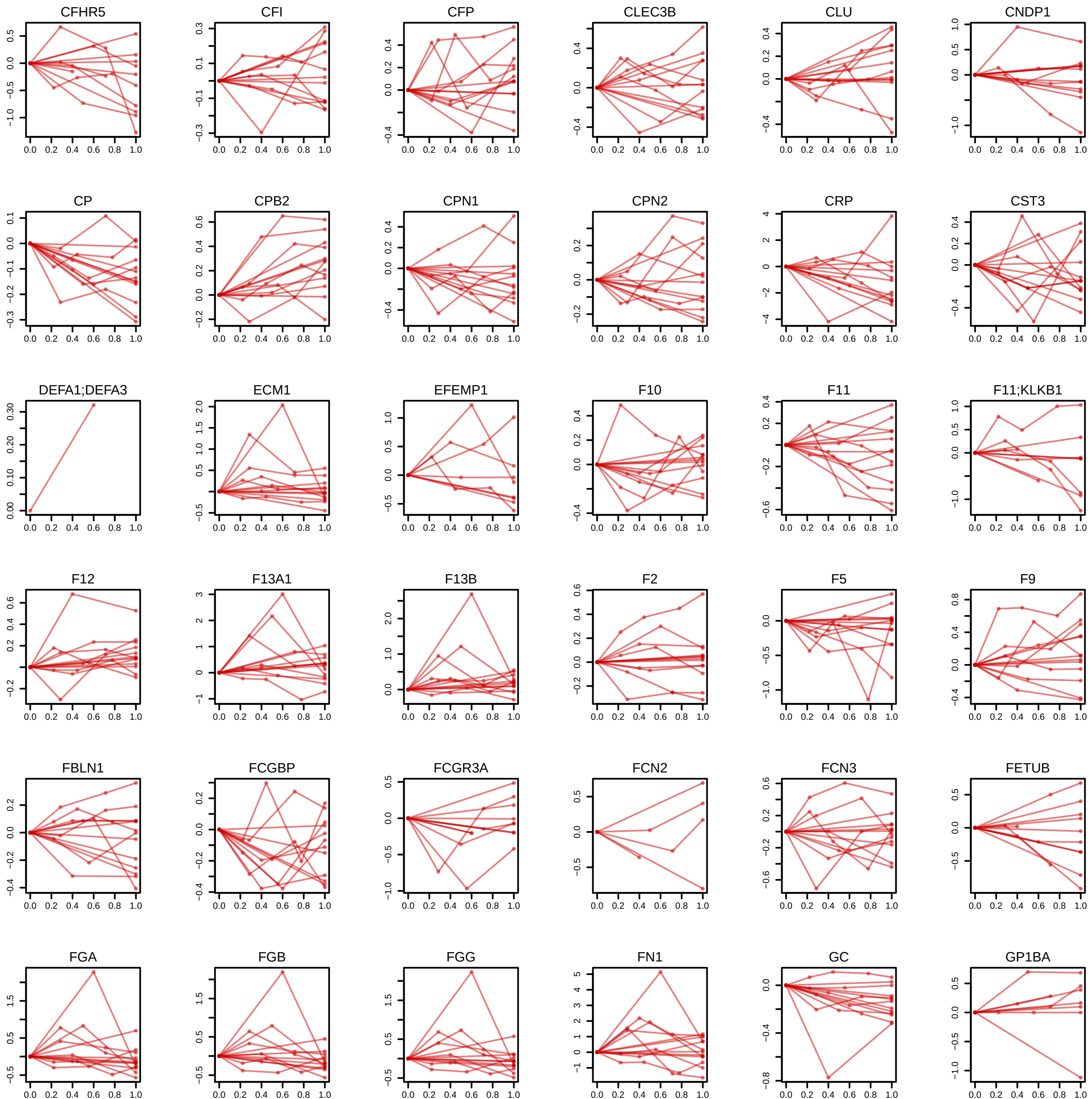

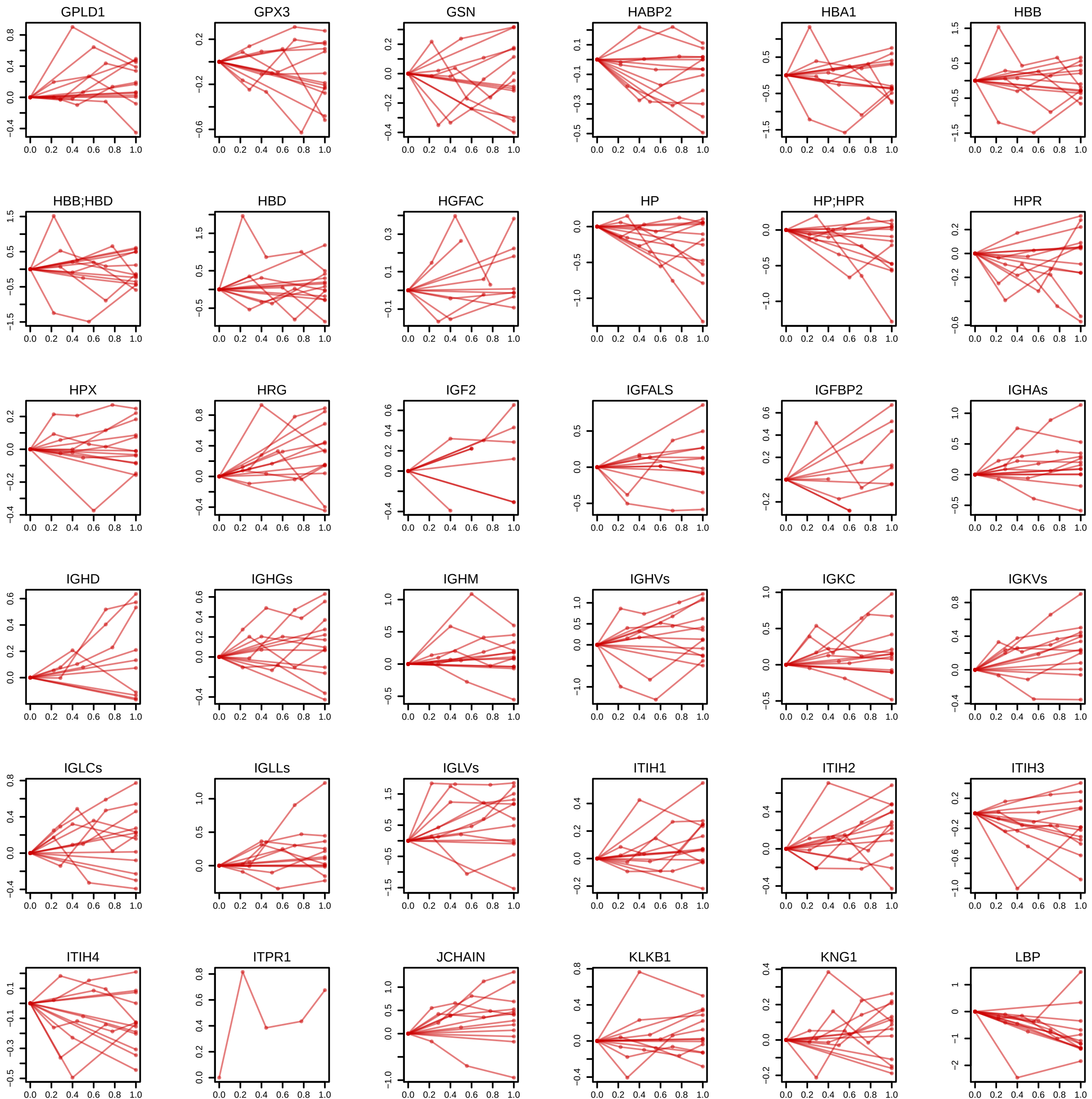

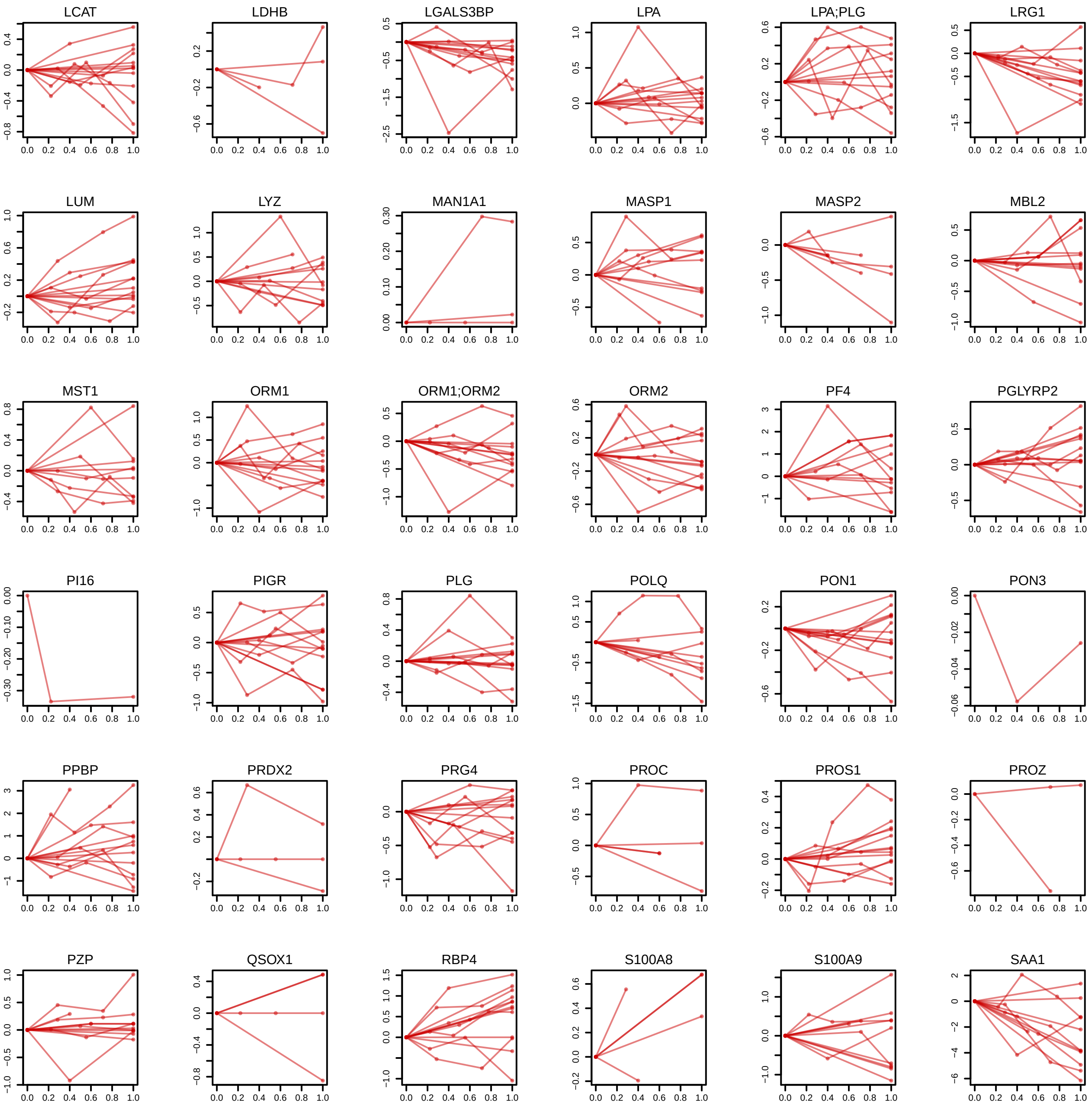

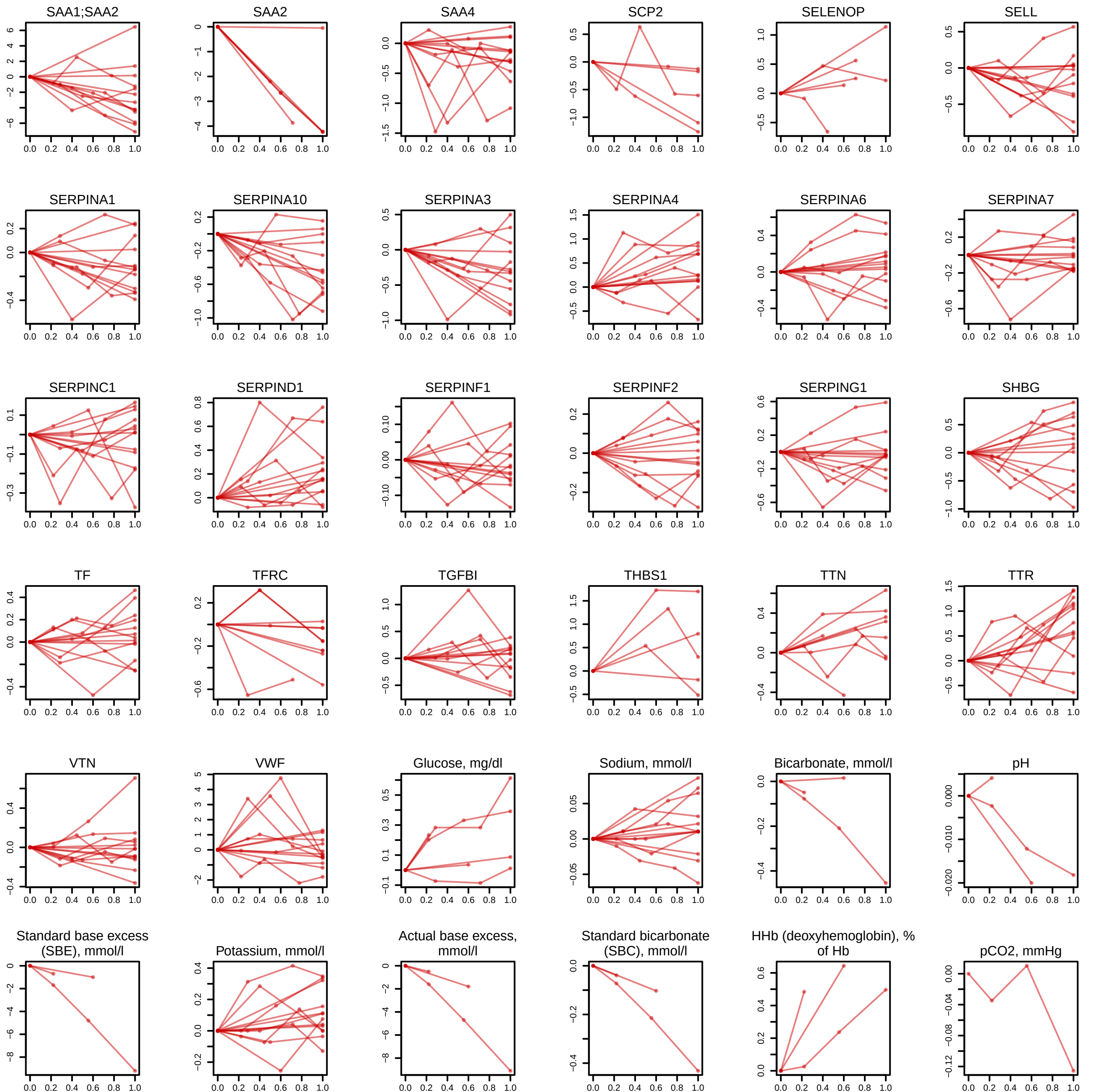

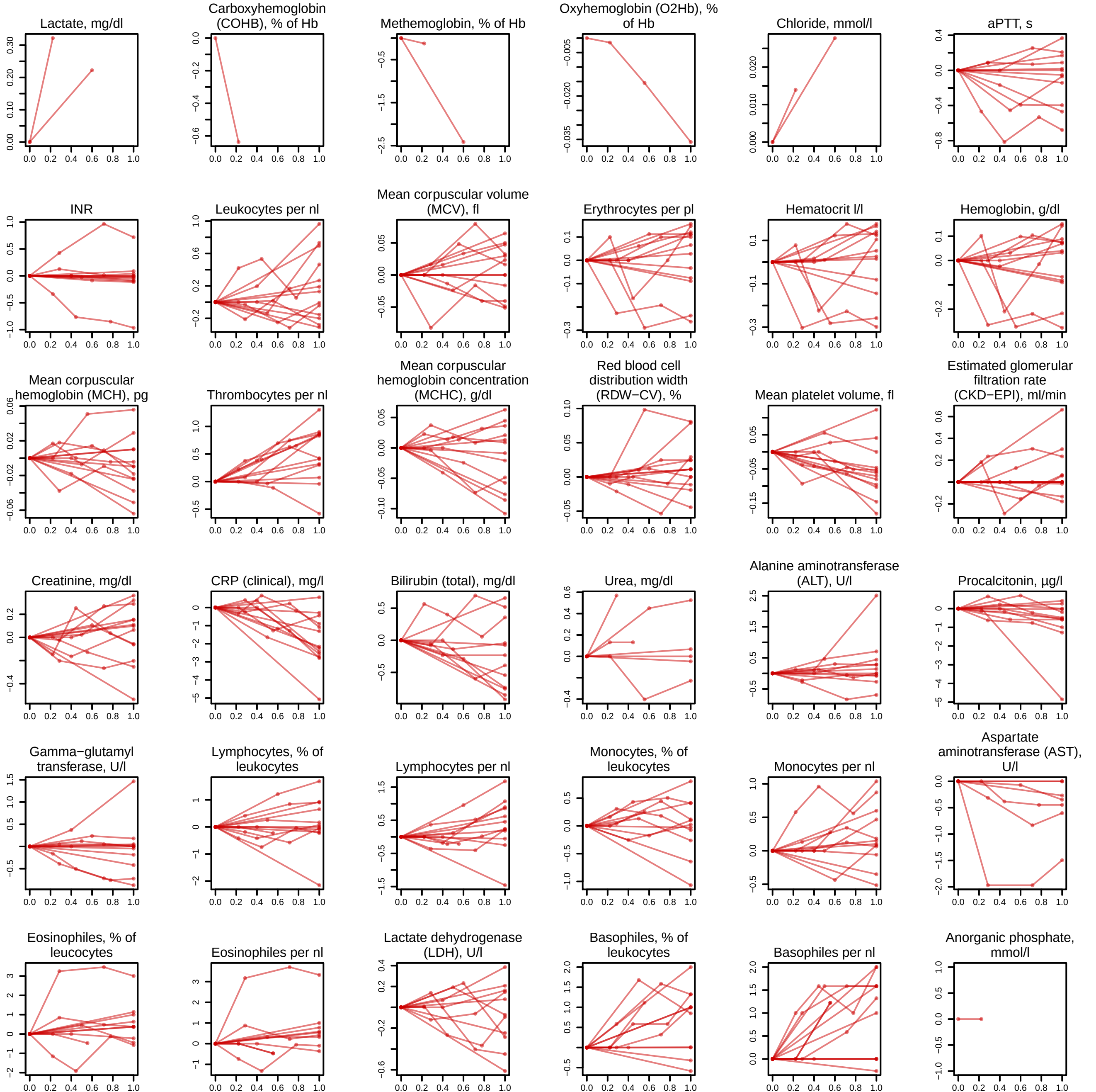

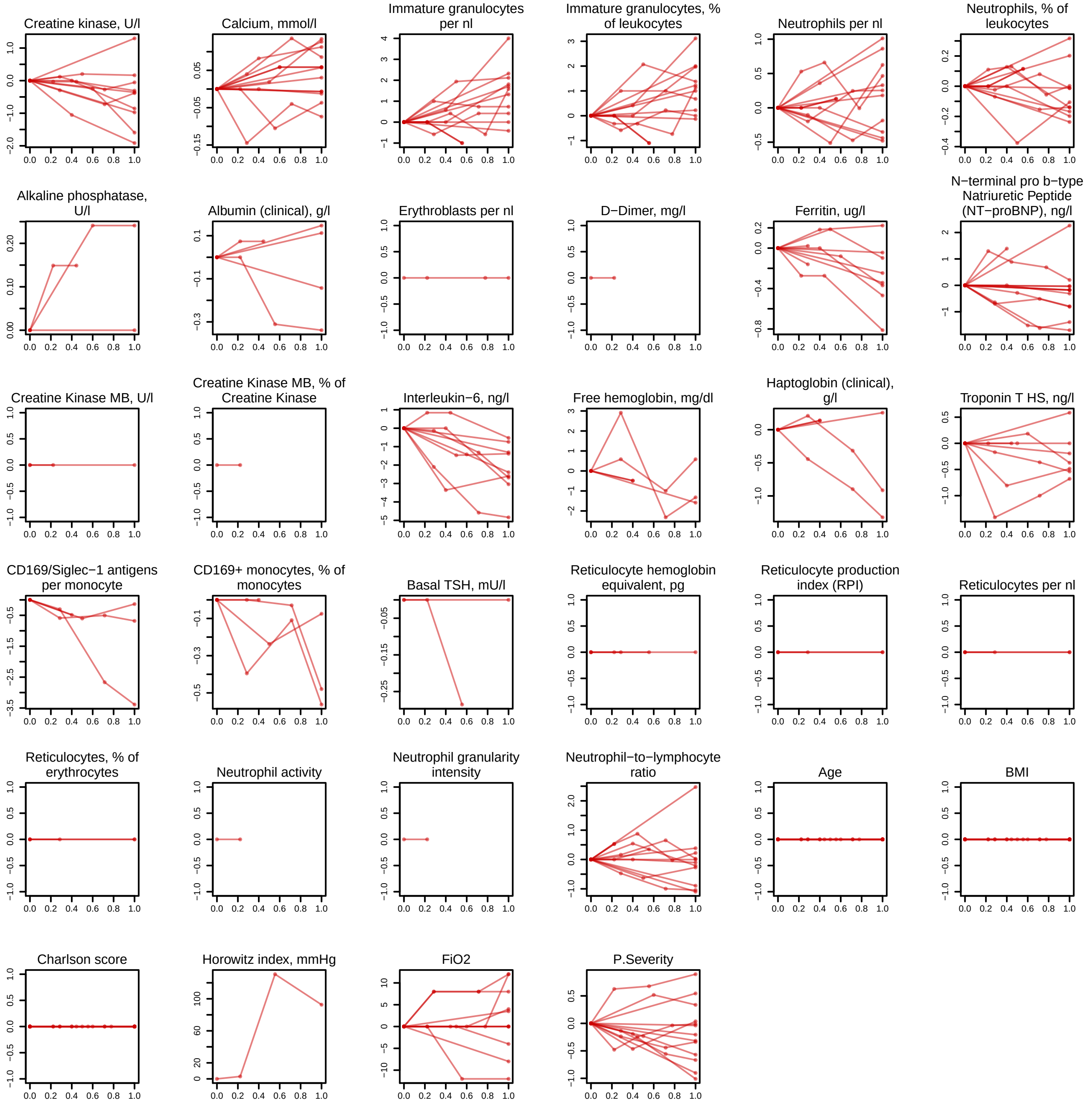

### SF17 - trajectories at the disease peak for max WHO = 5.pdf

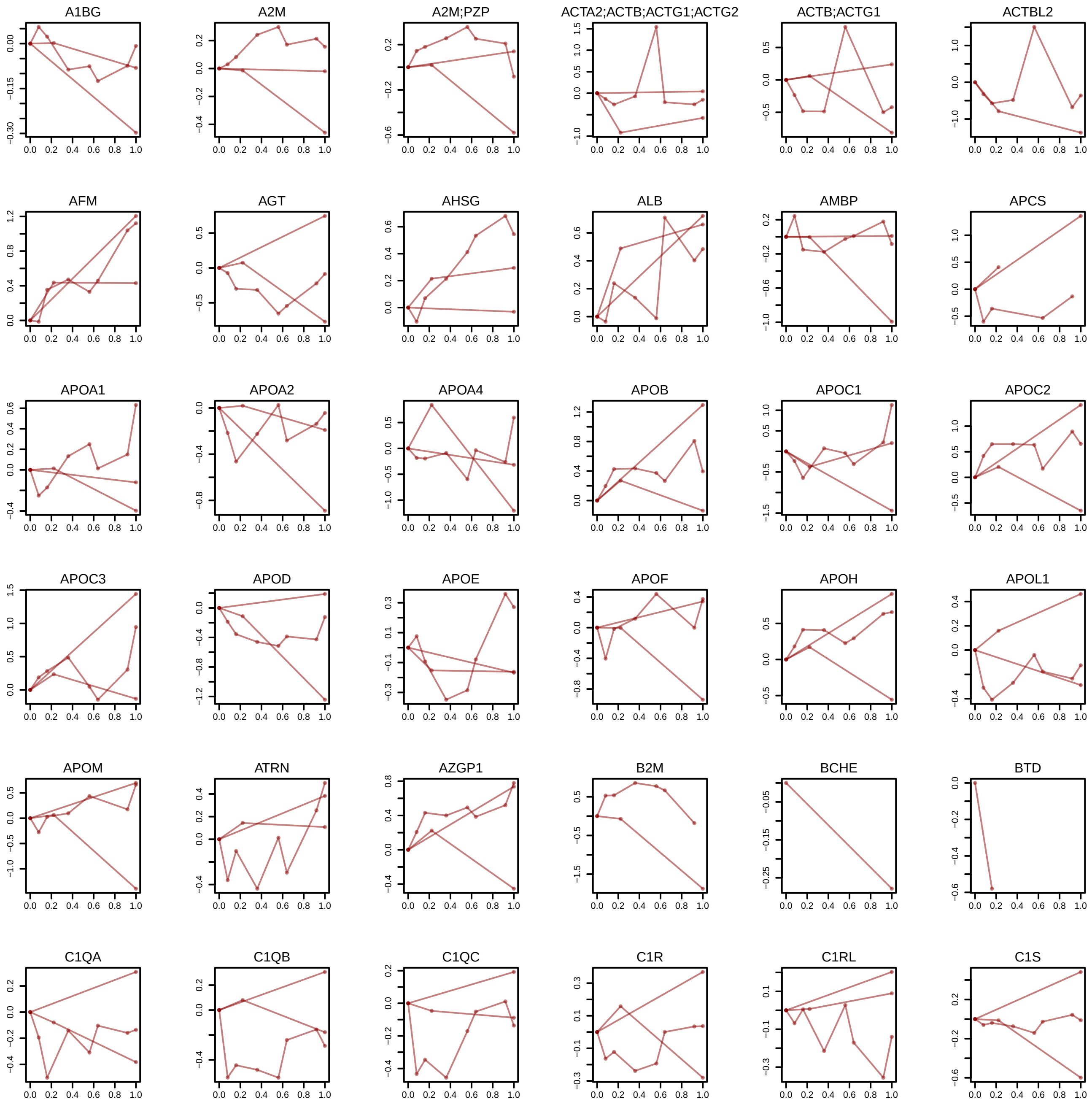

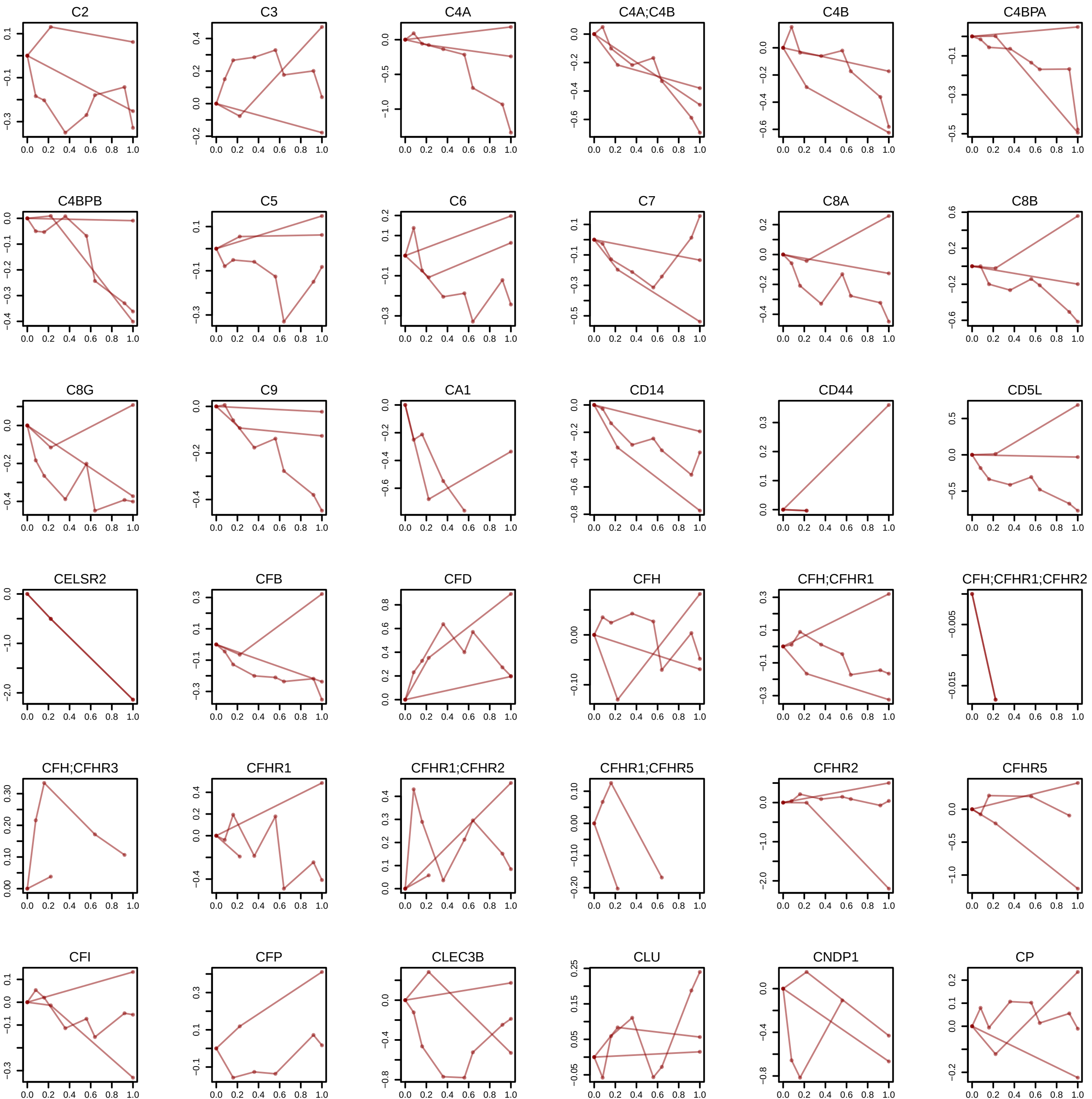

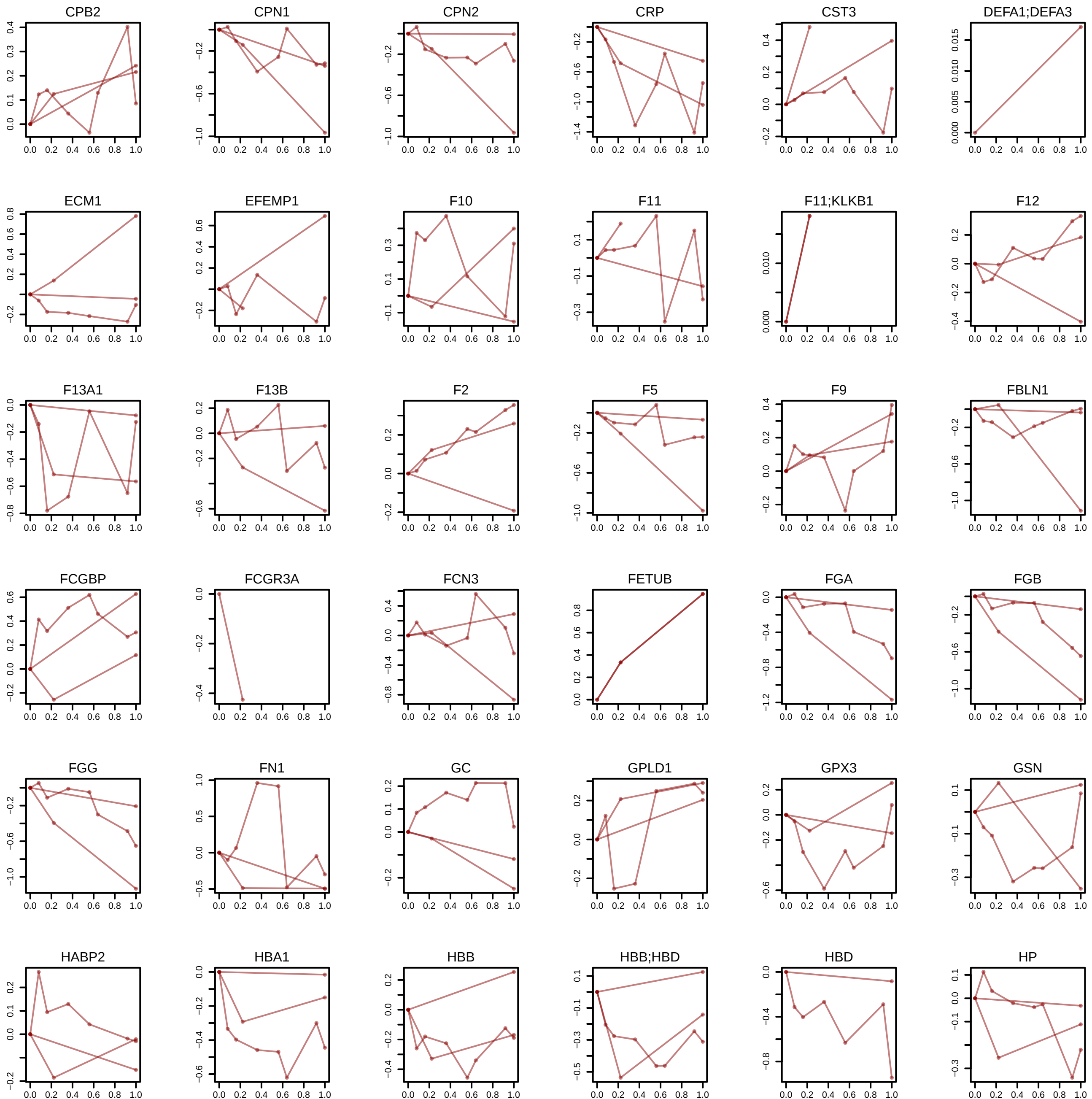

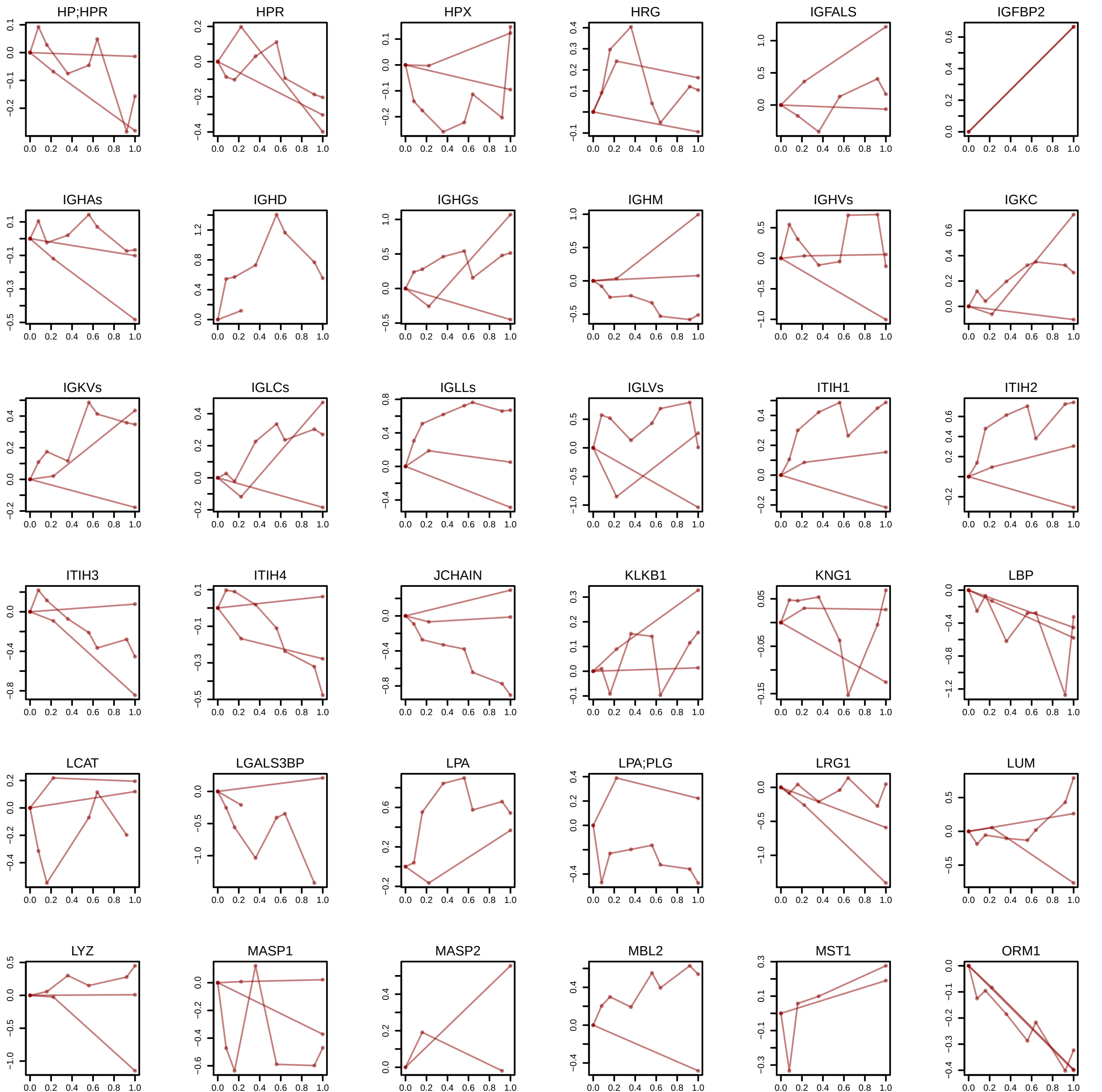

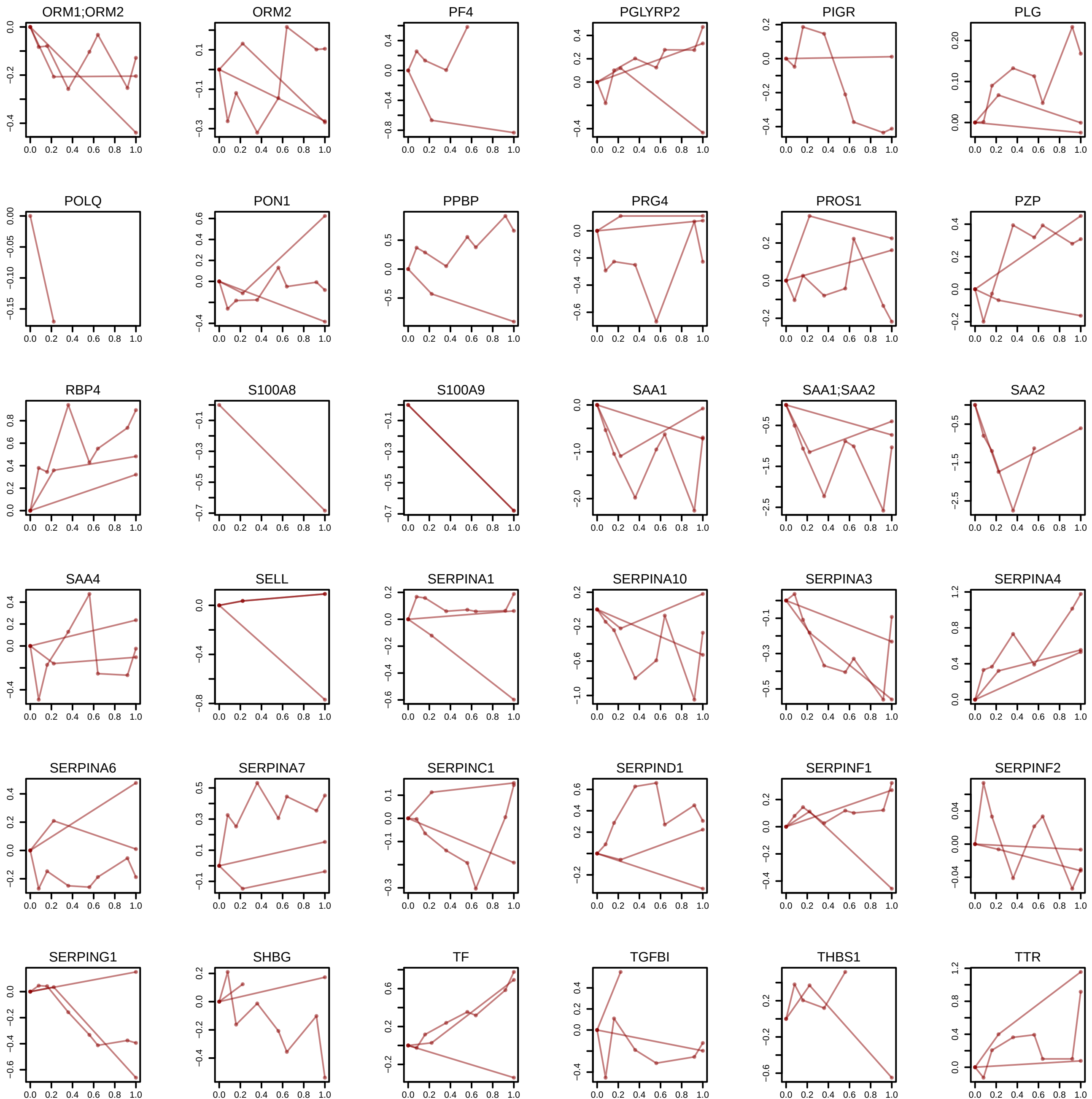

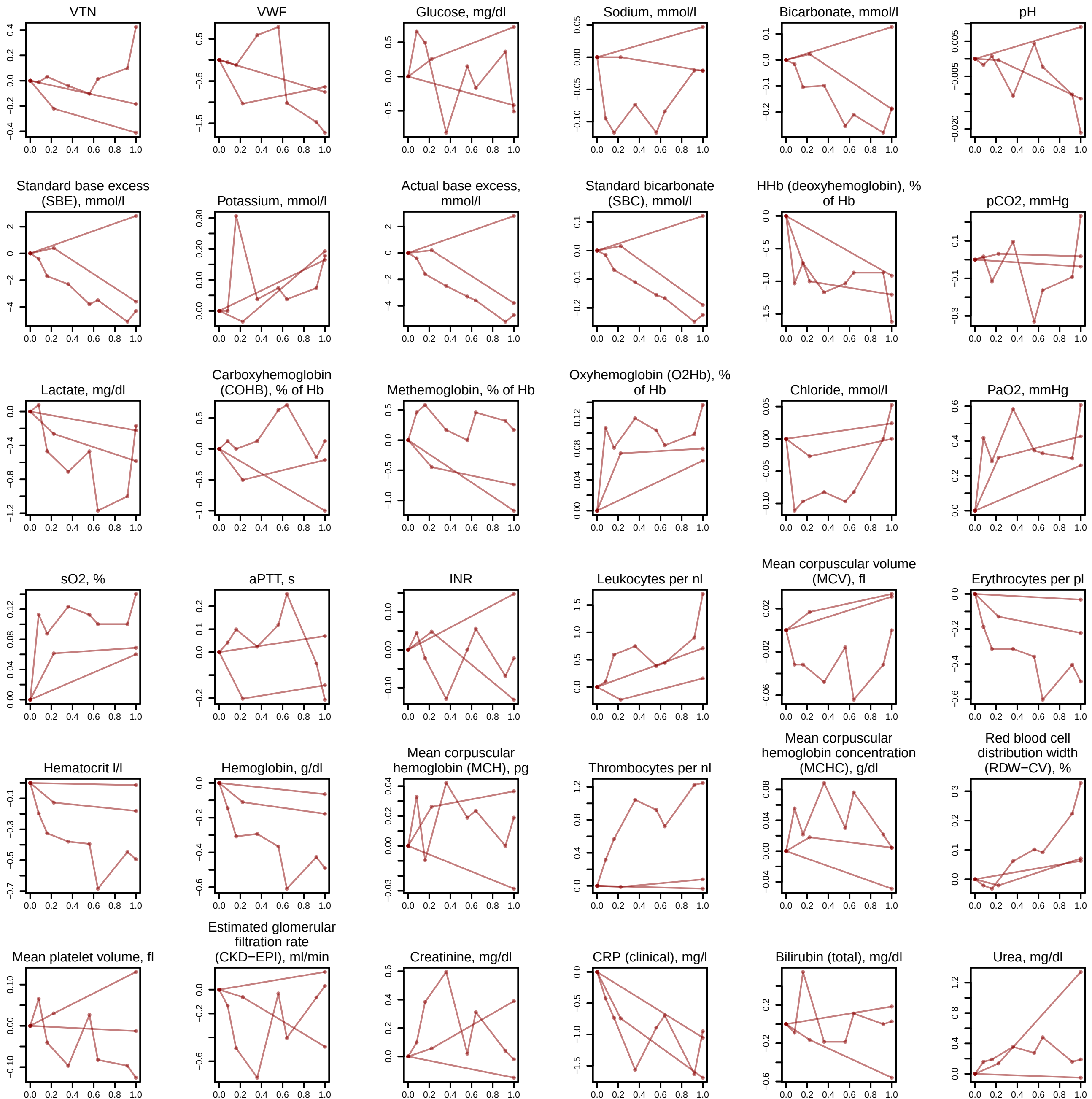

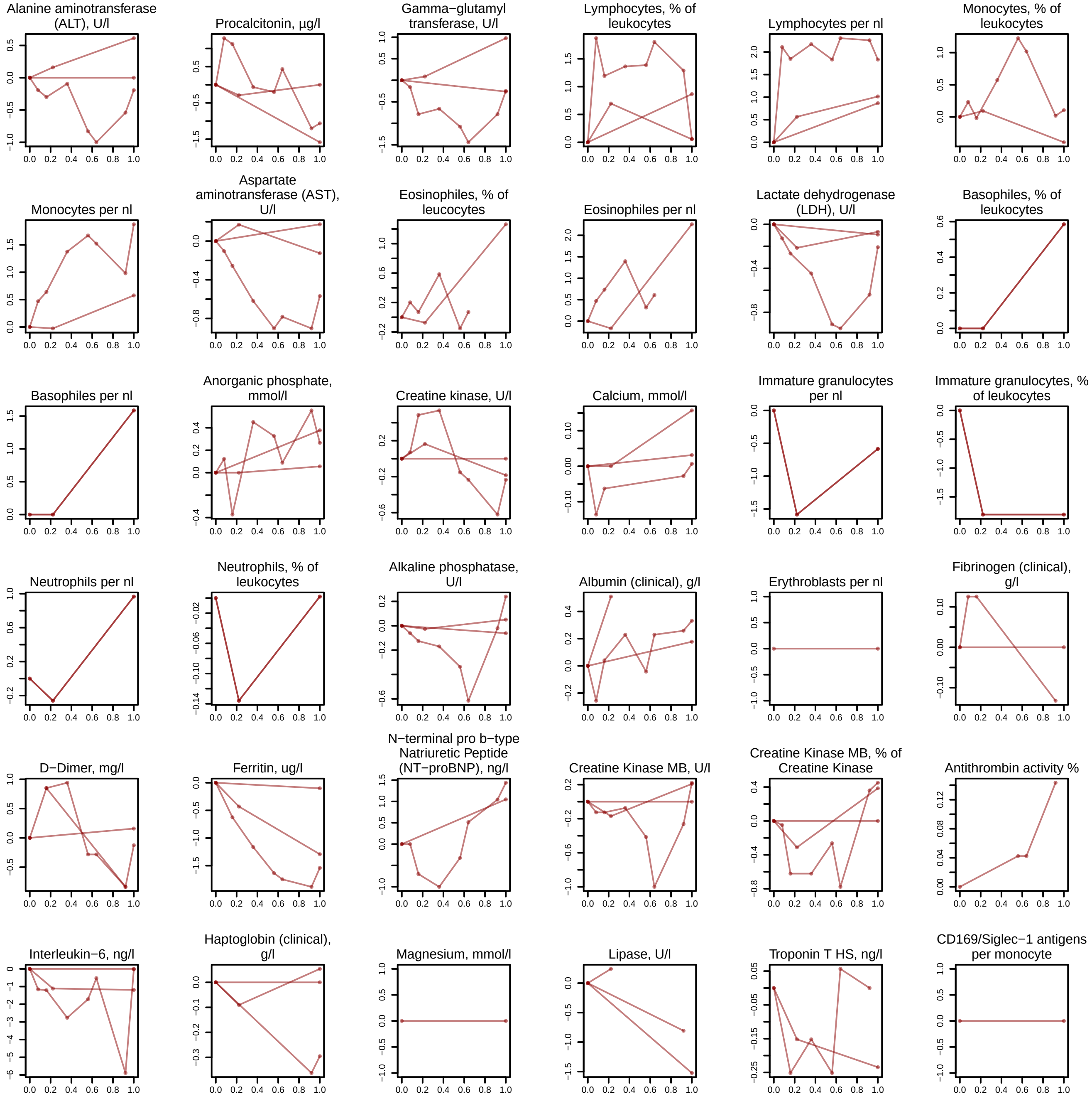

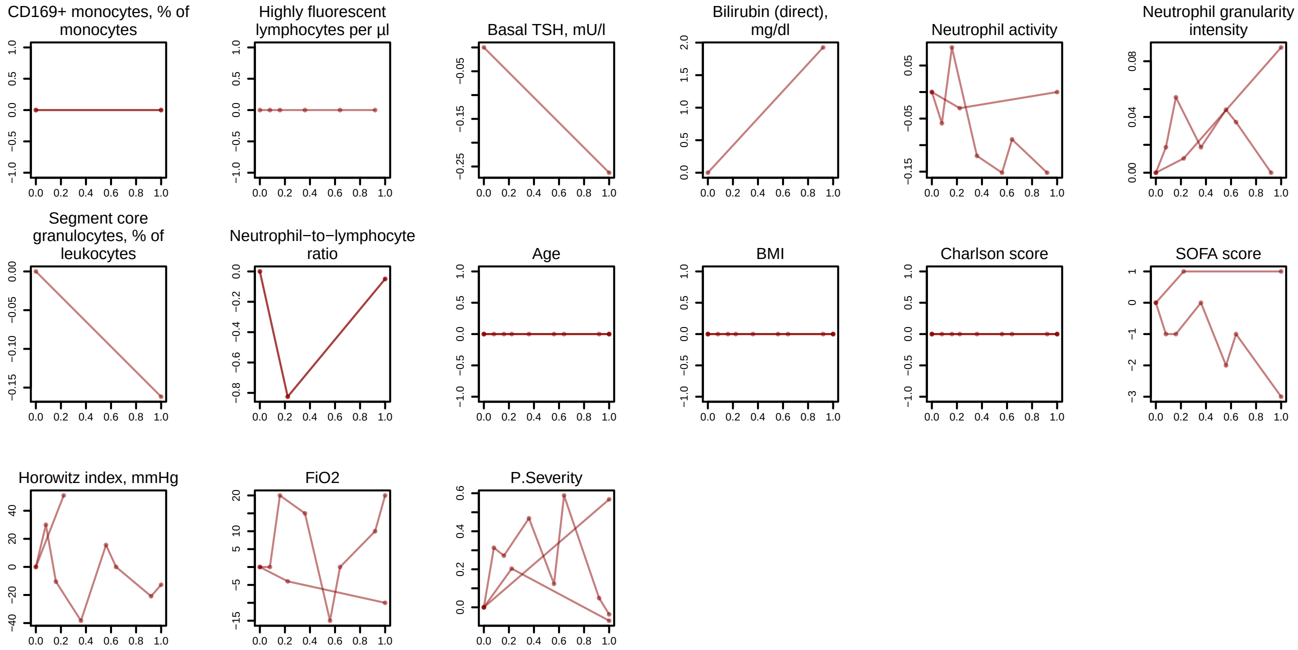

### SF18 - trajectories at the disease peak for max WHO = 6.pdf

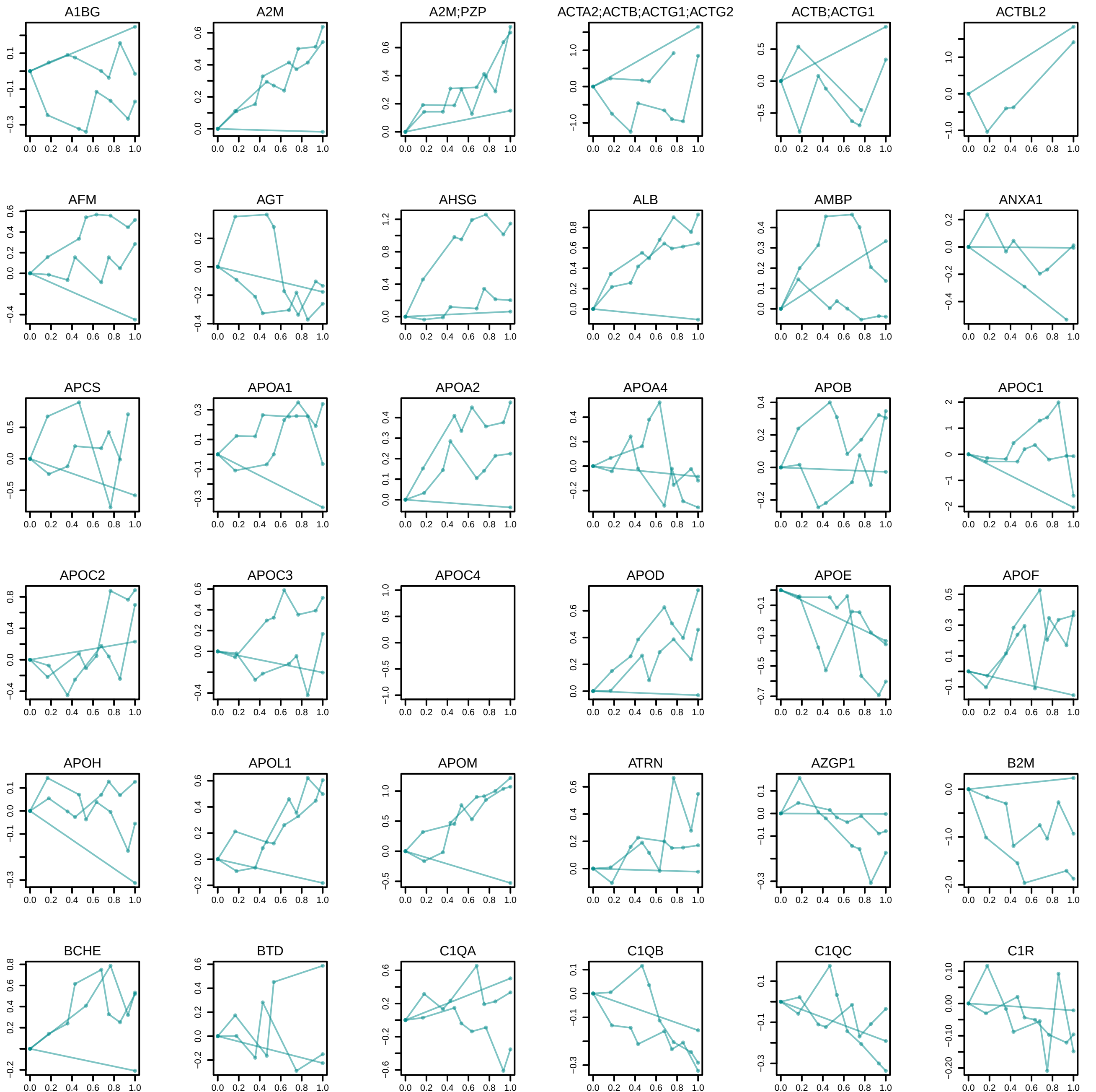

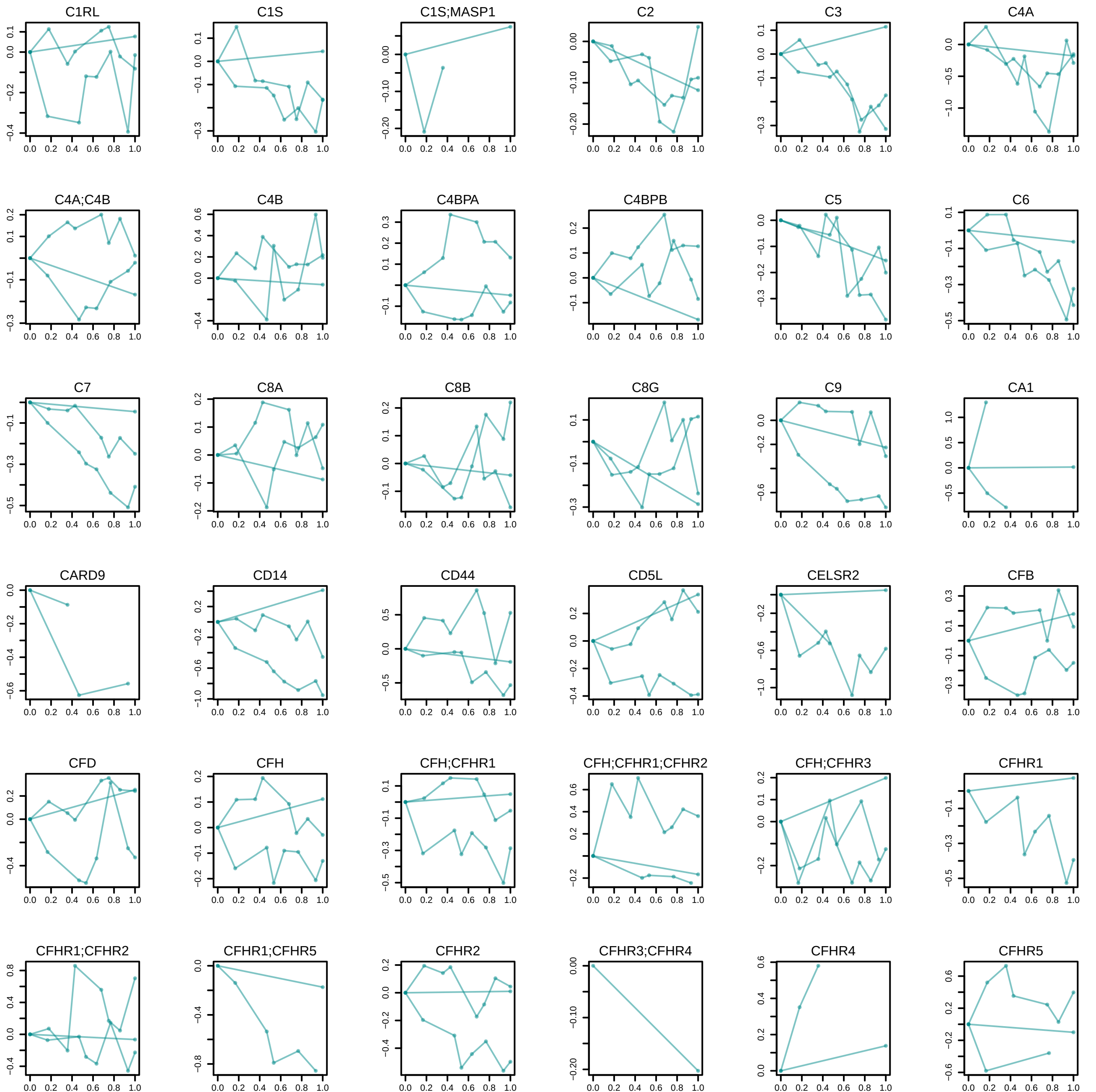

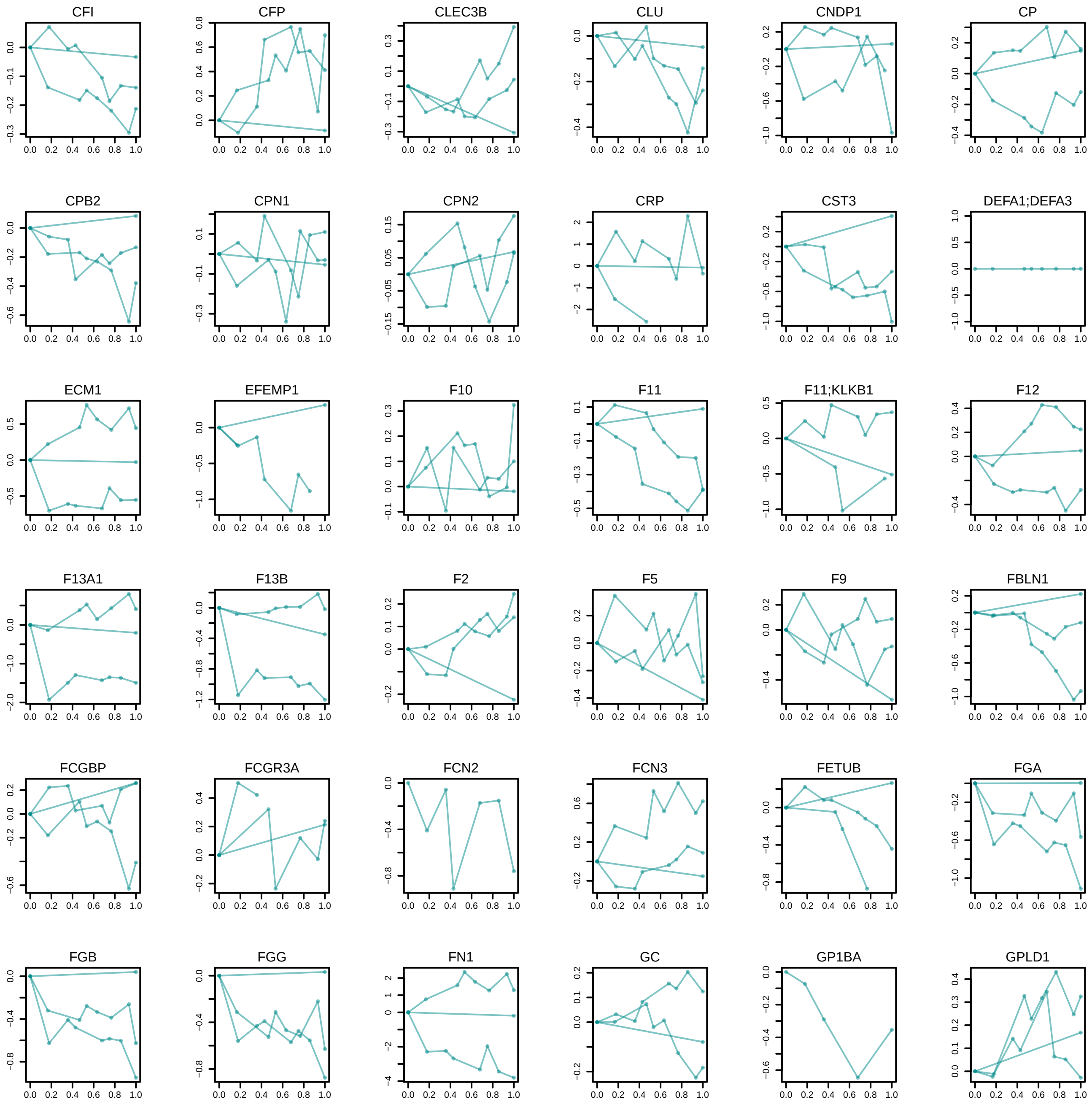

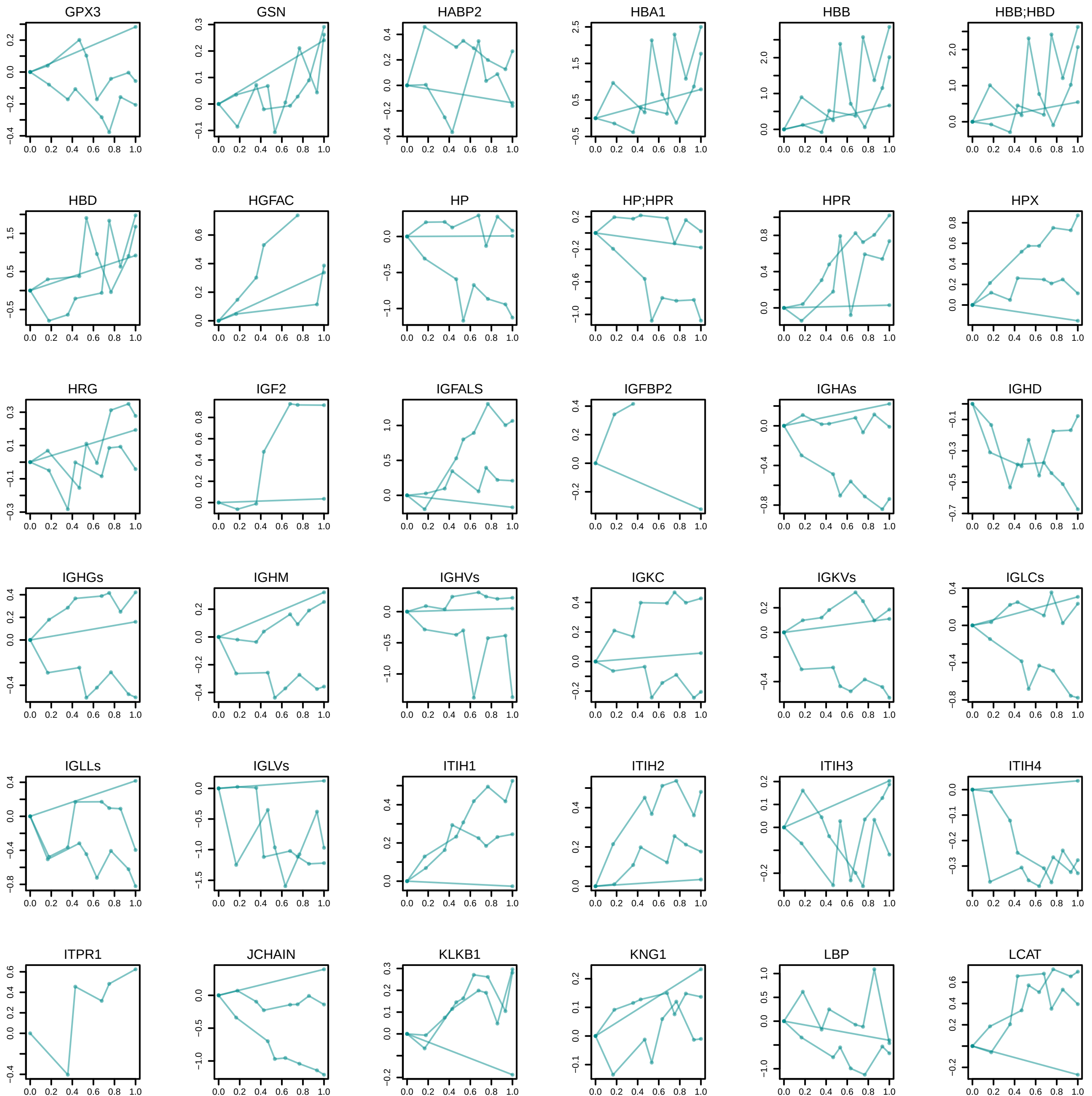

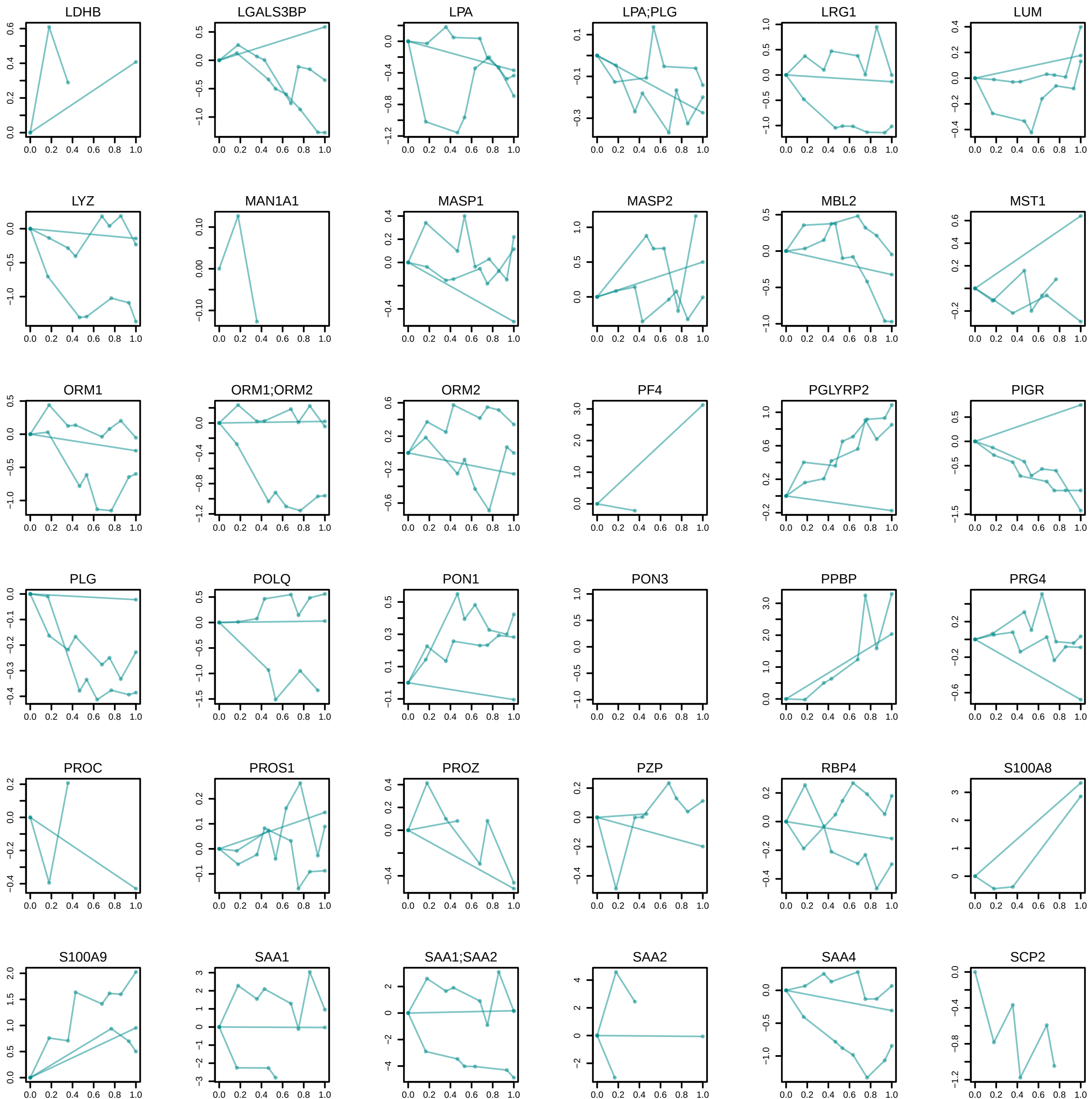

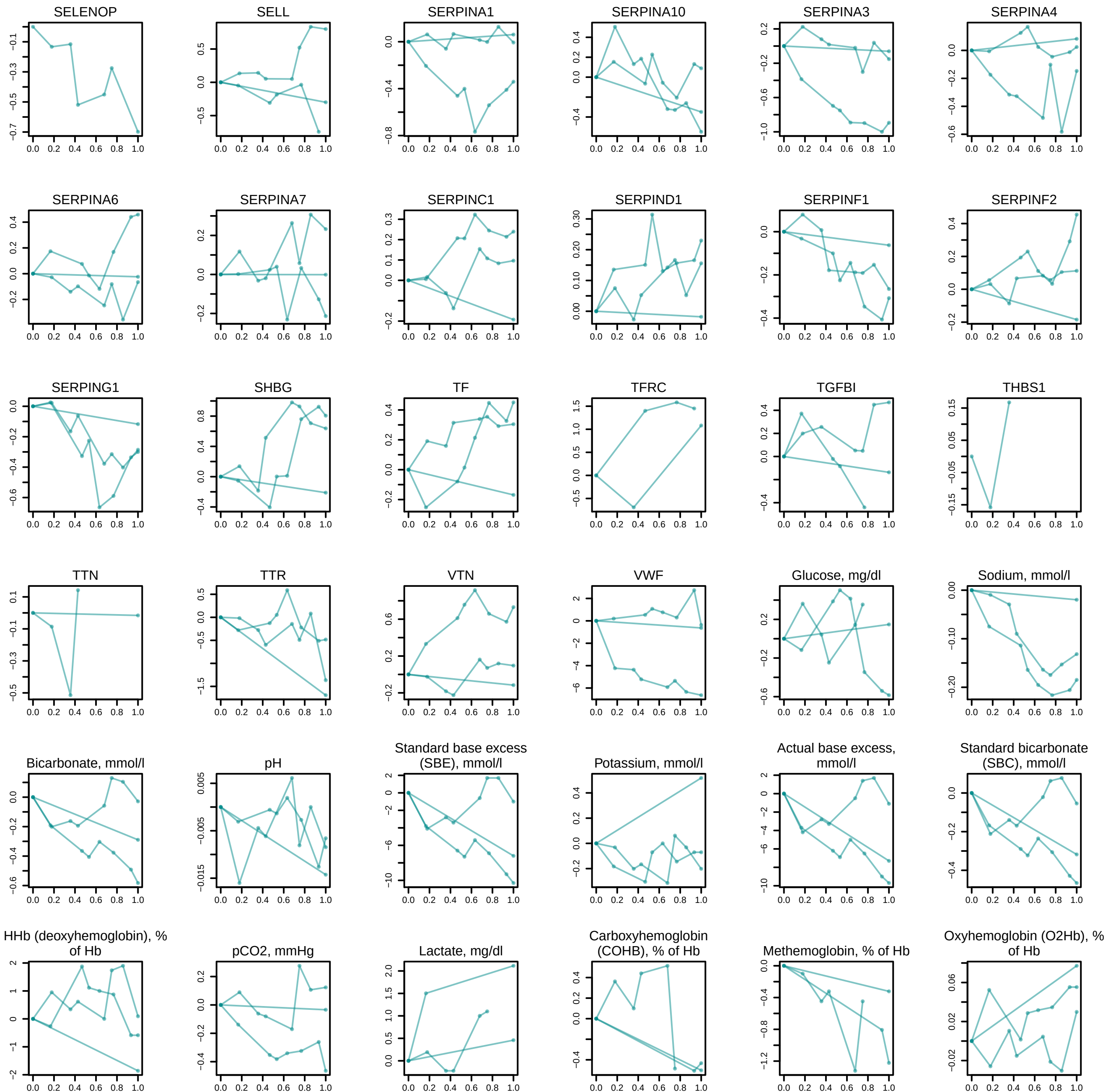

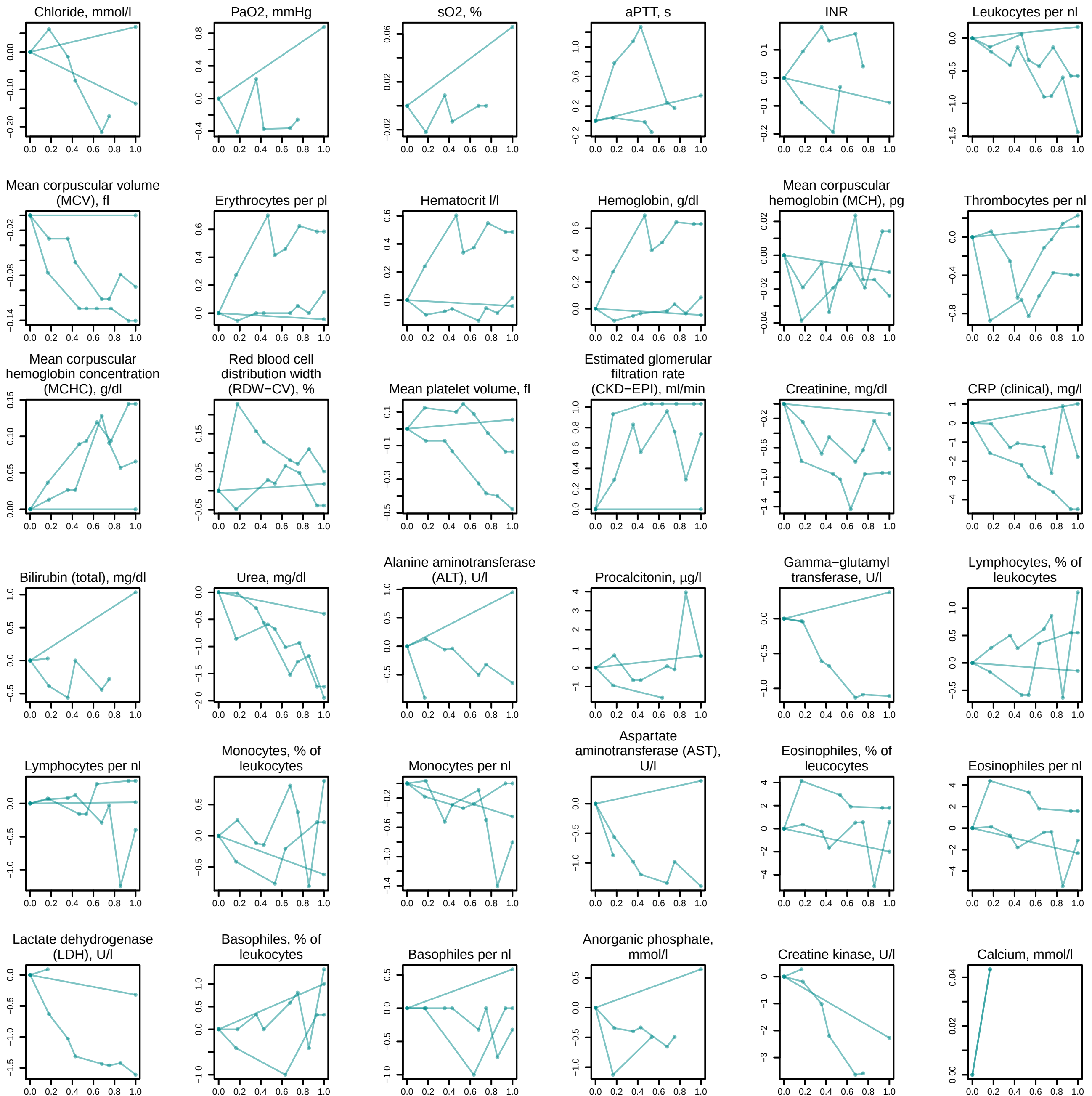

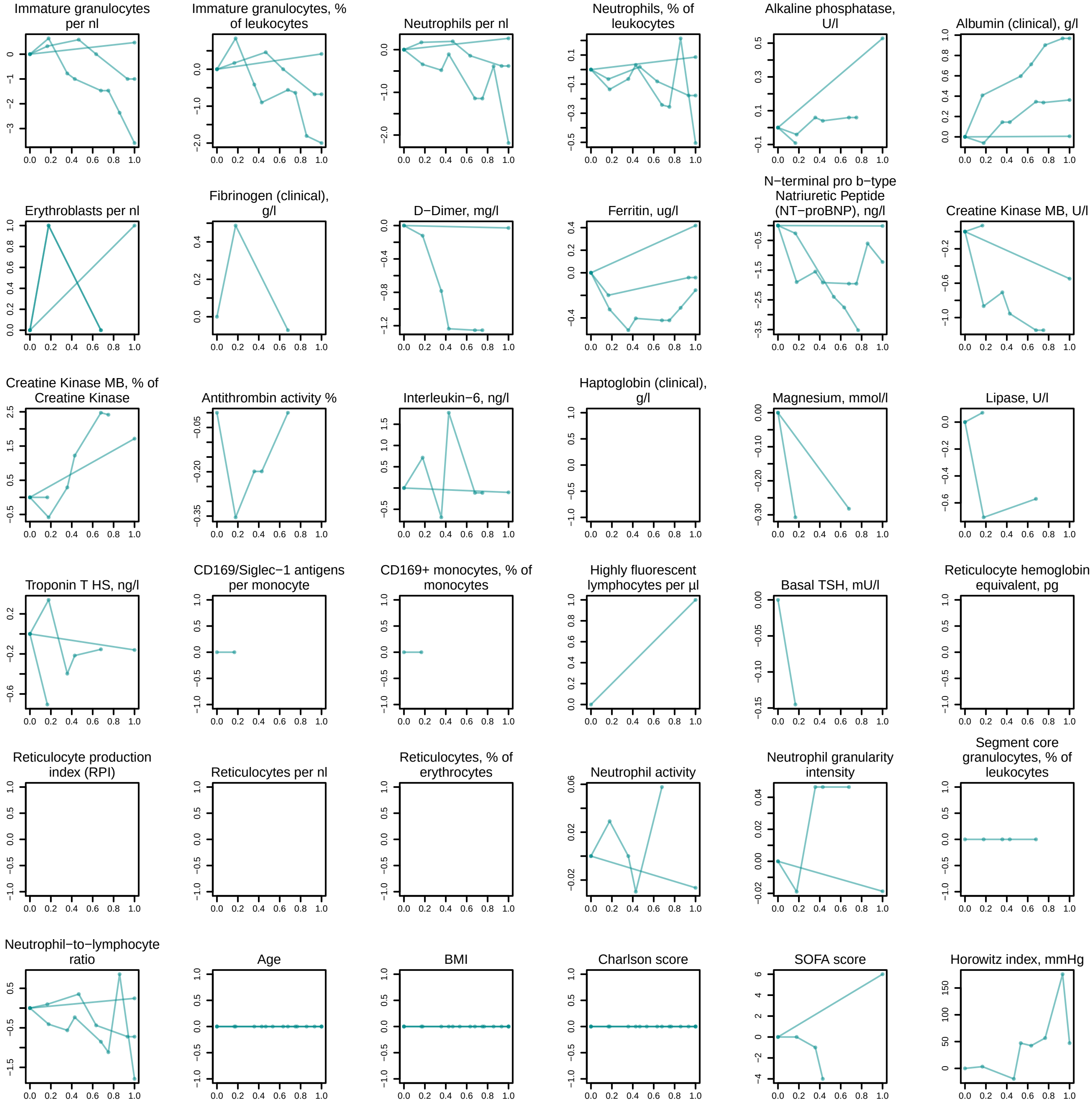

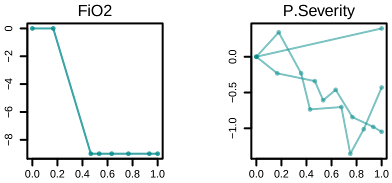

### SF19 - trajectories at the disease peak for max WHO = 7 - patients who died highlighted in black.pdf

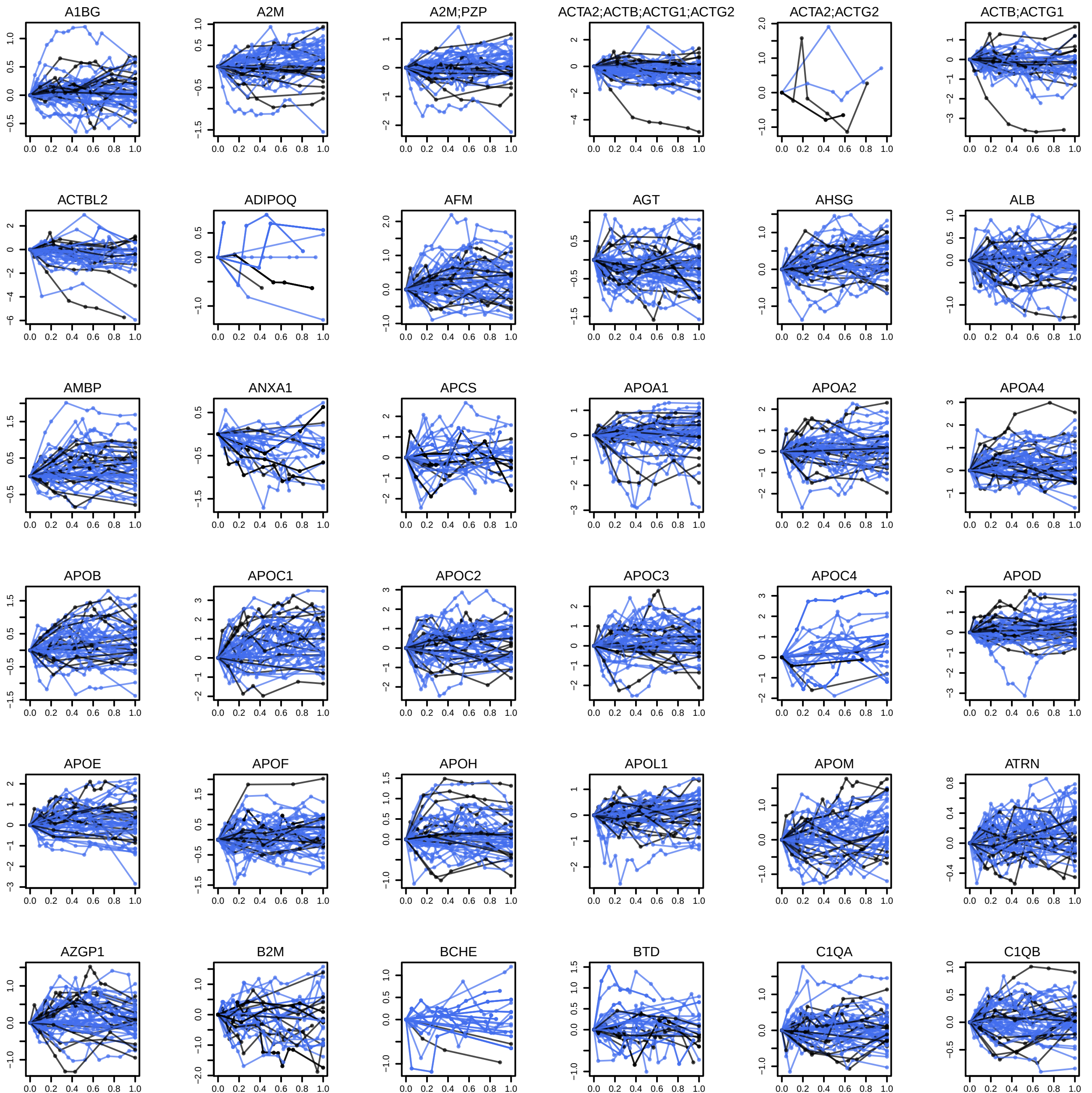

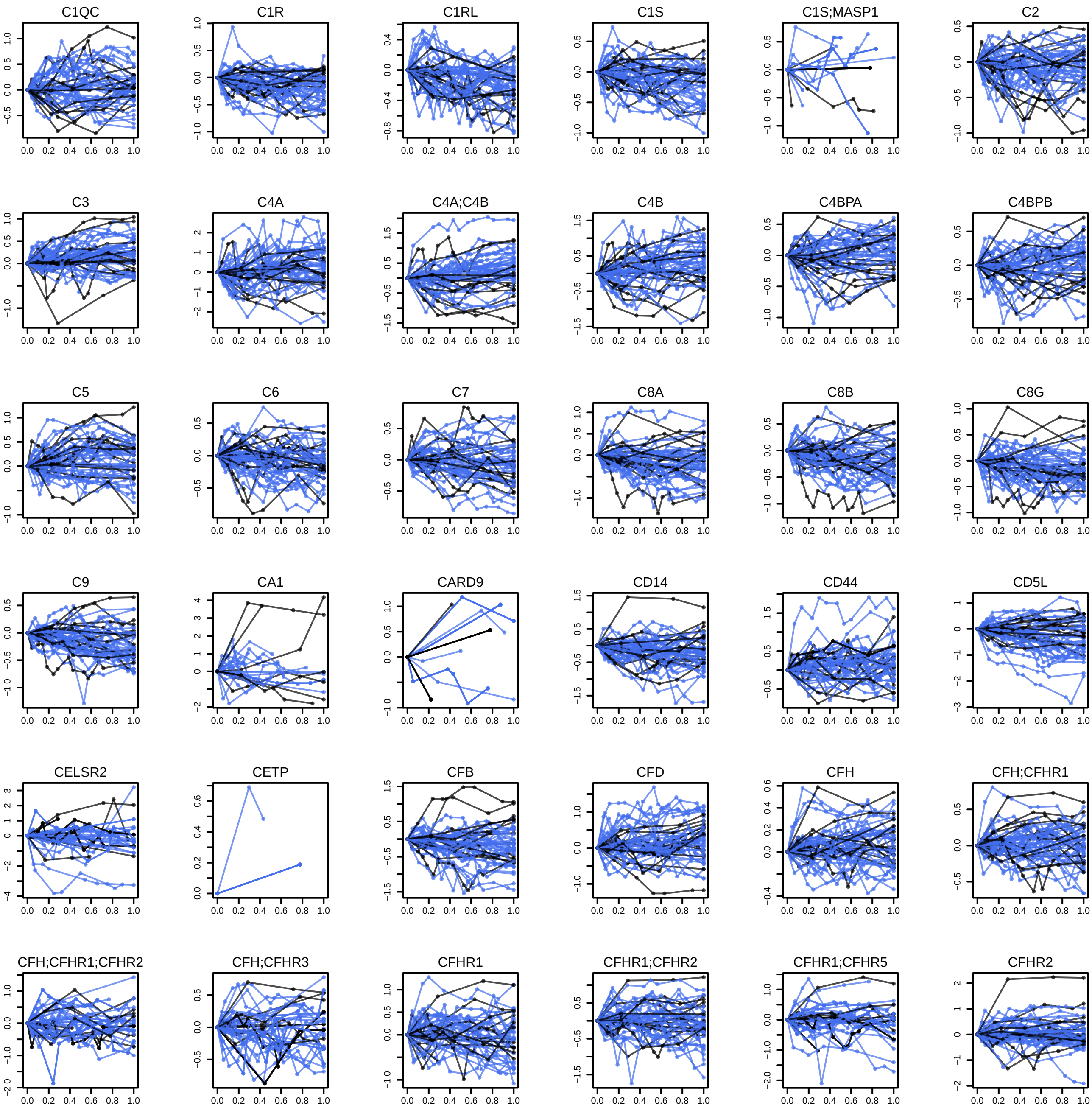

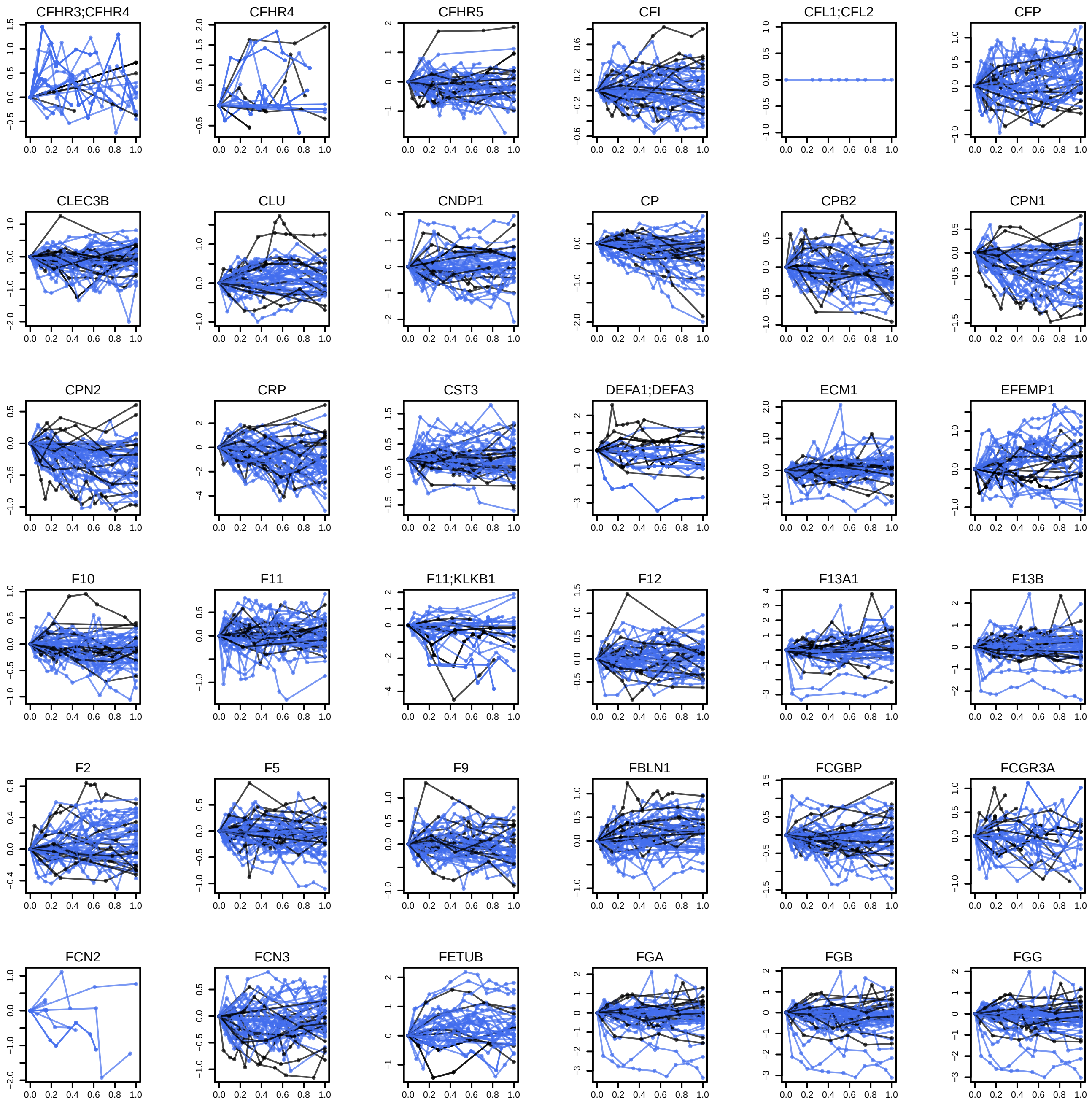

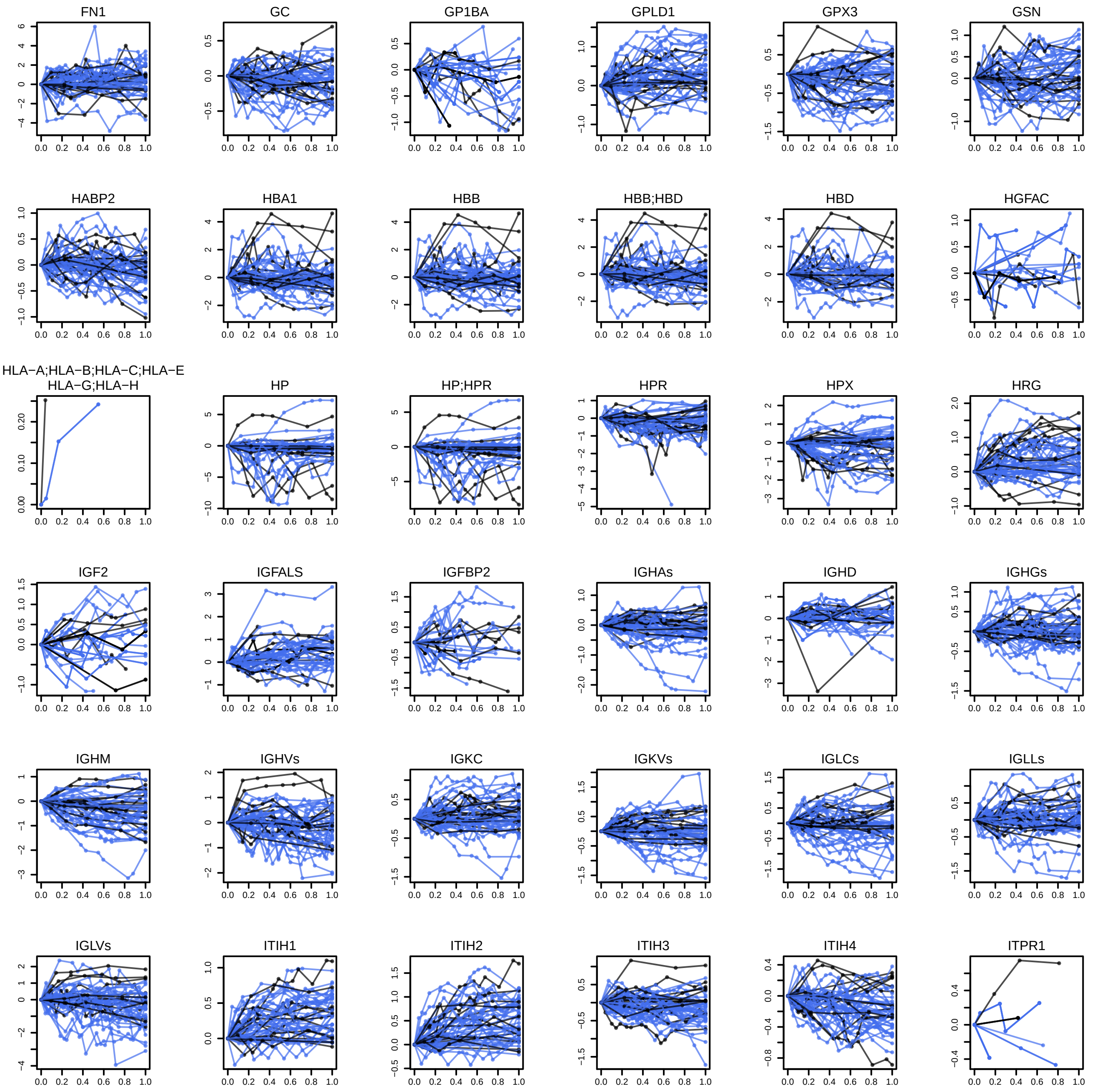

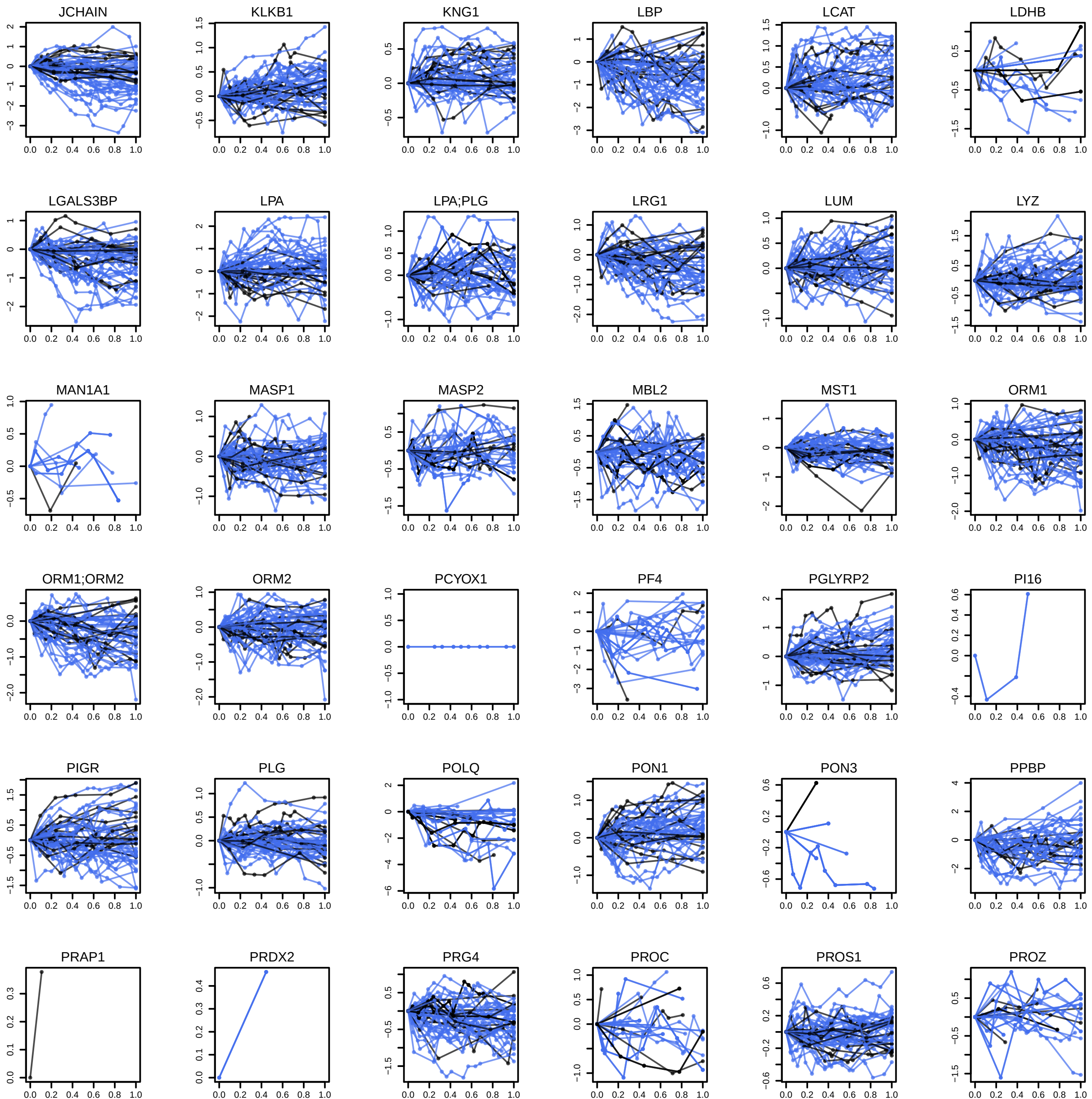

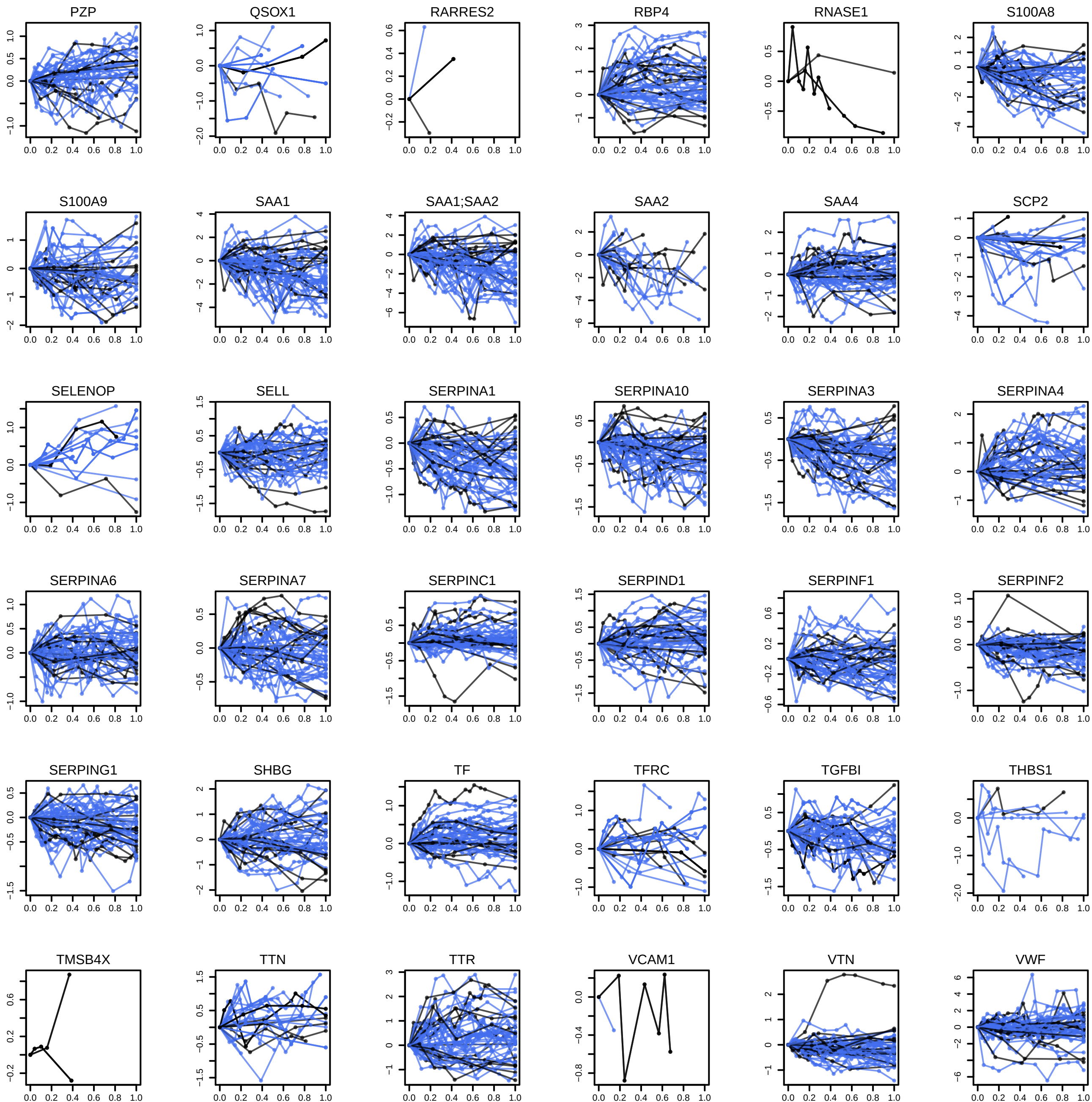

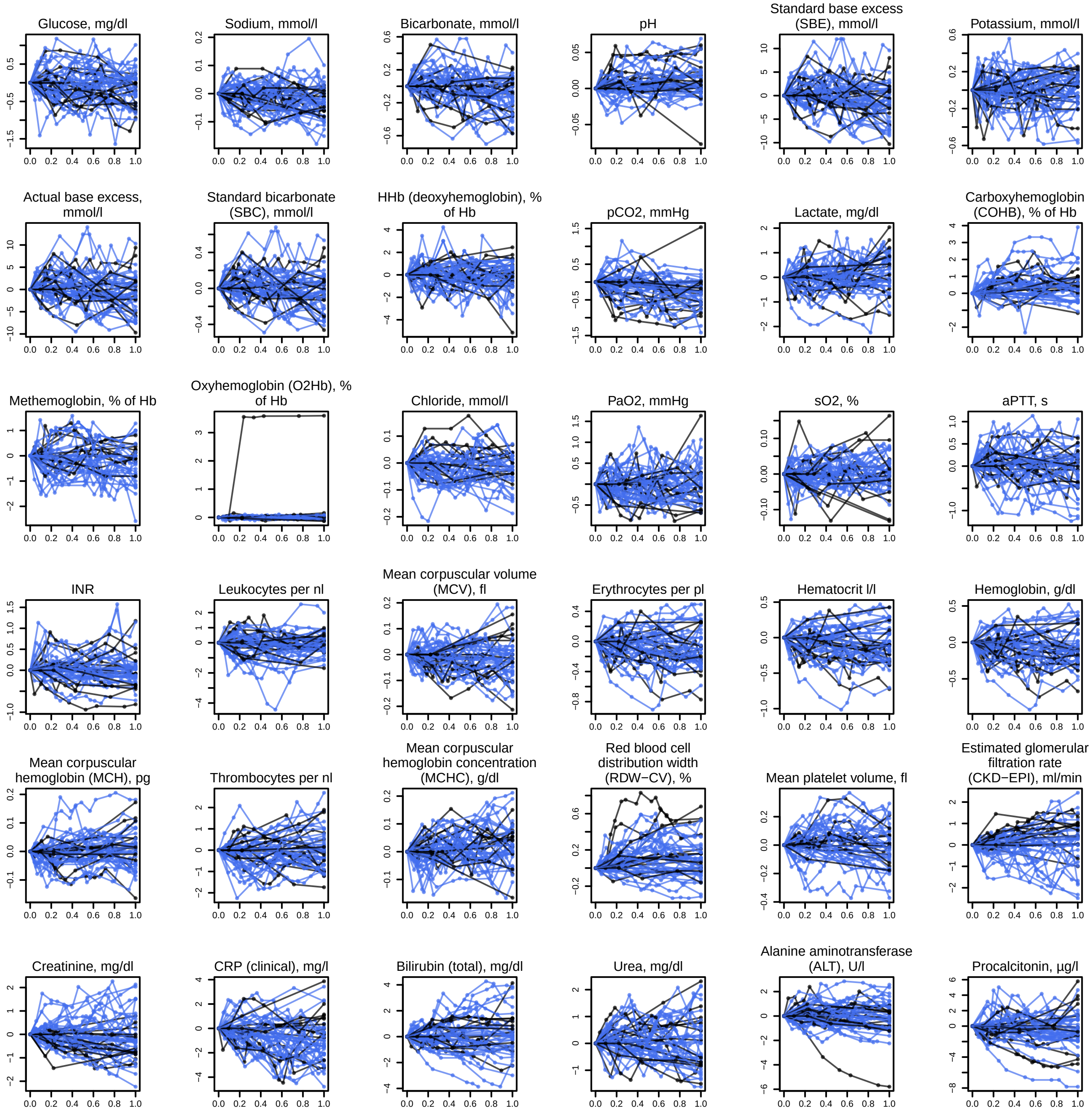

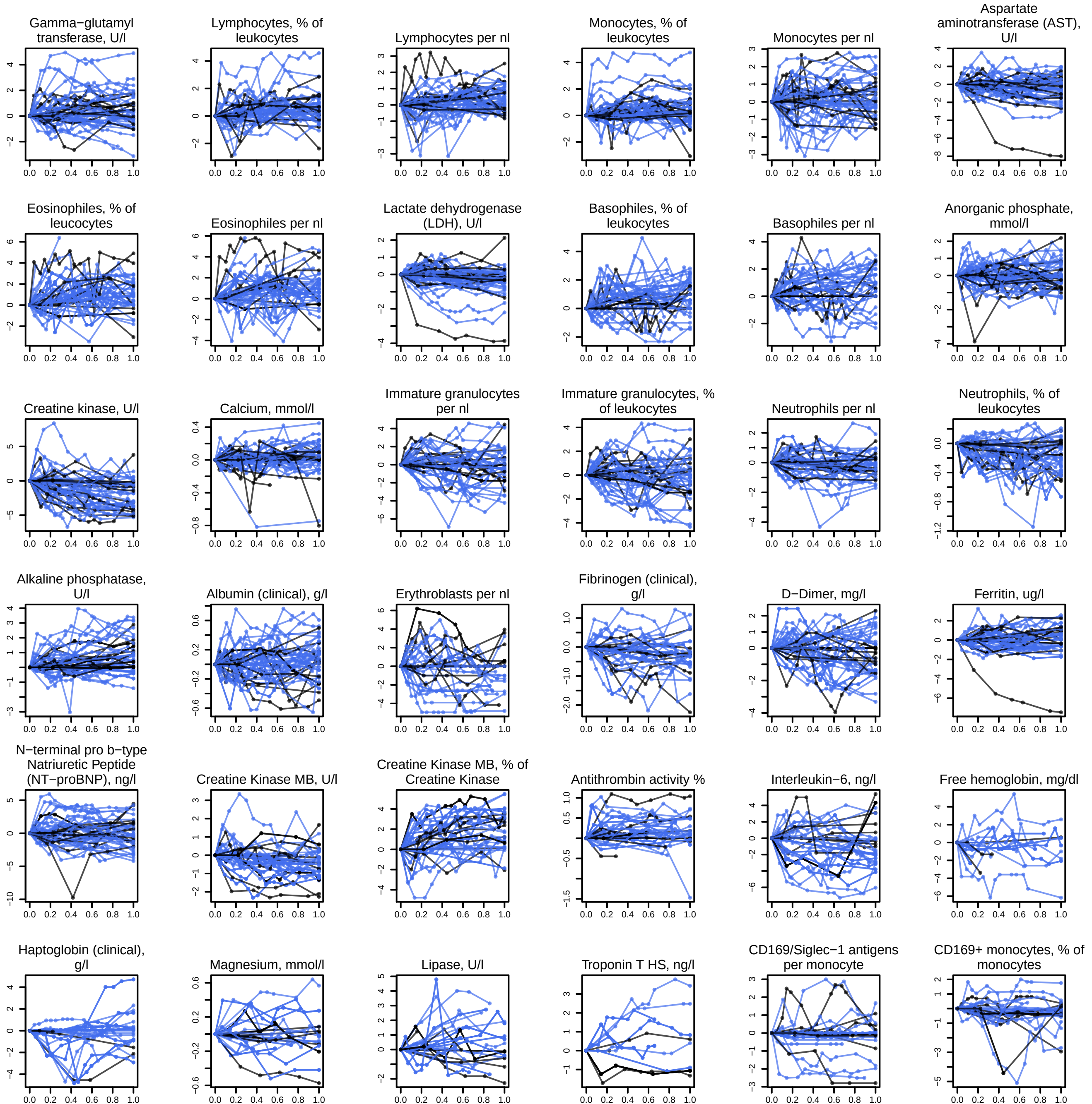

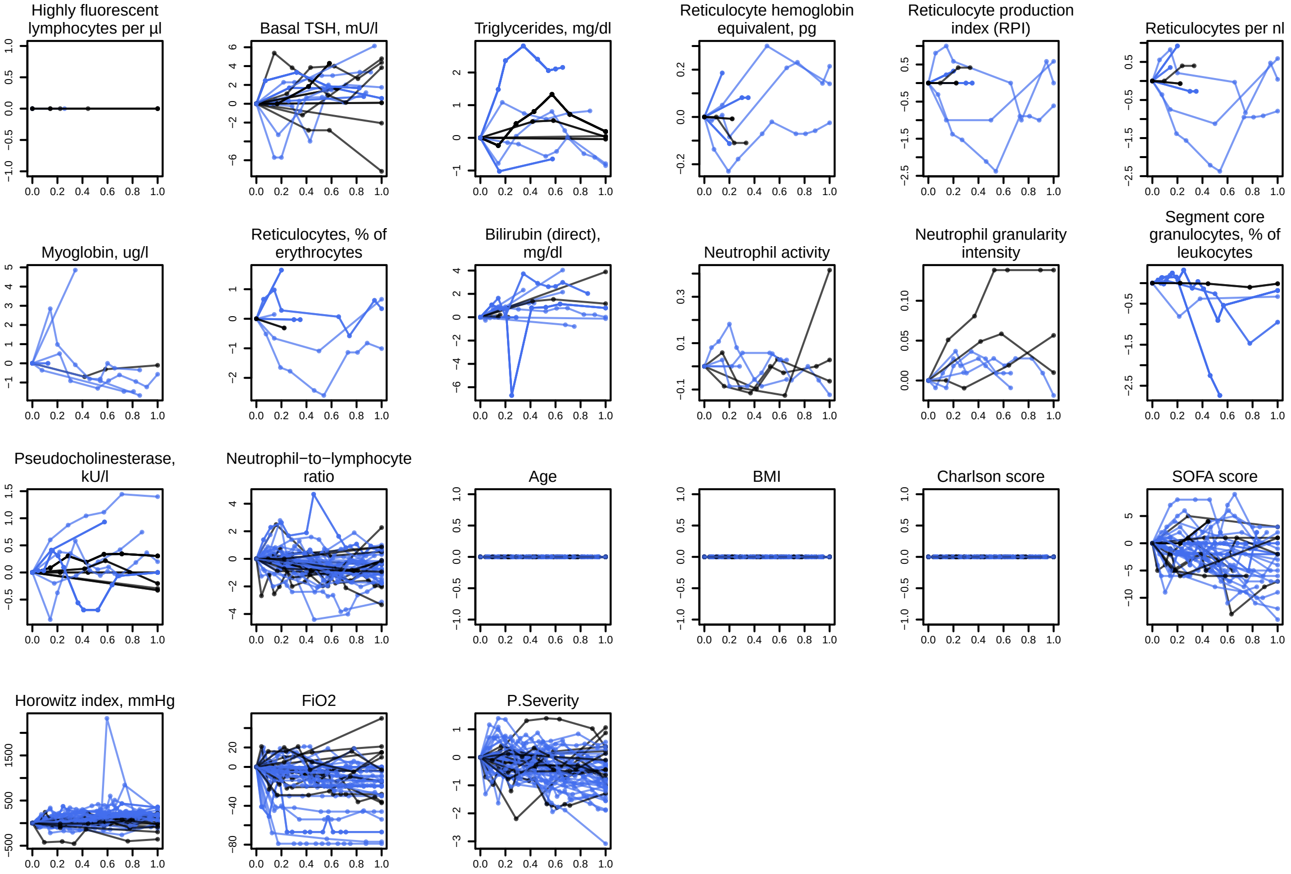

### SF20 - omics features predictive of the remaining time in hospital for patients at WHO = 3.pdf

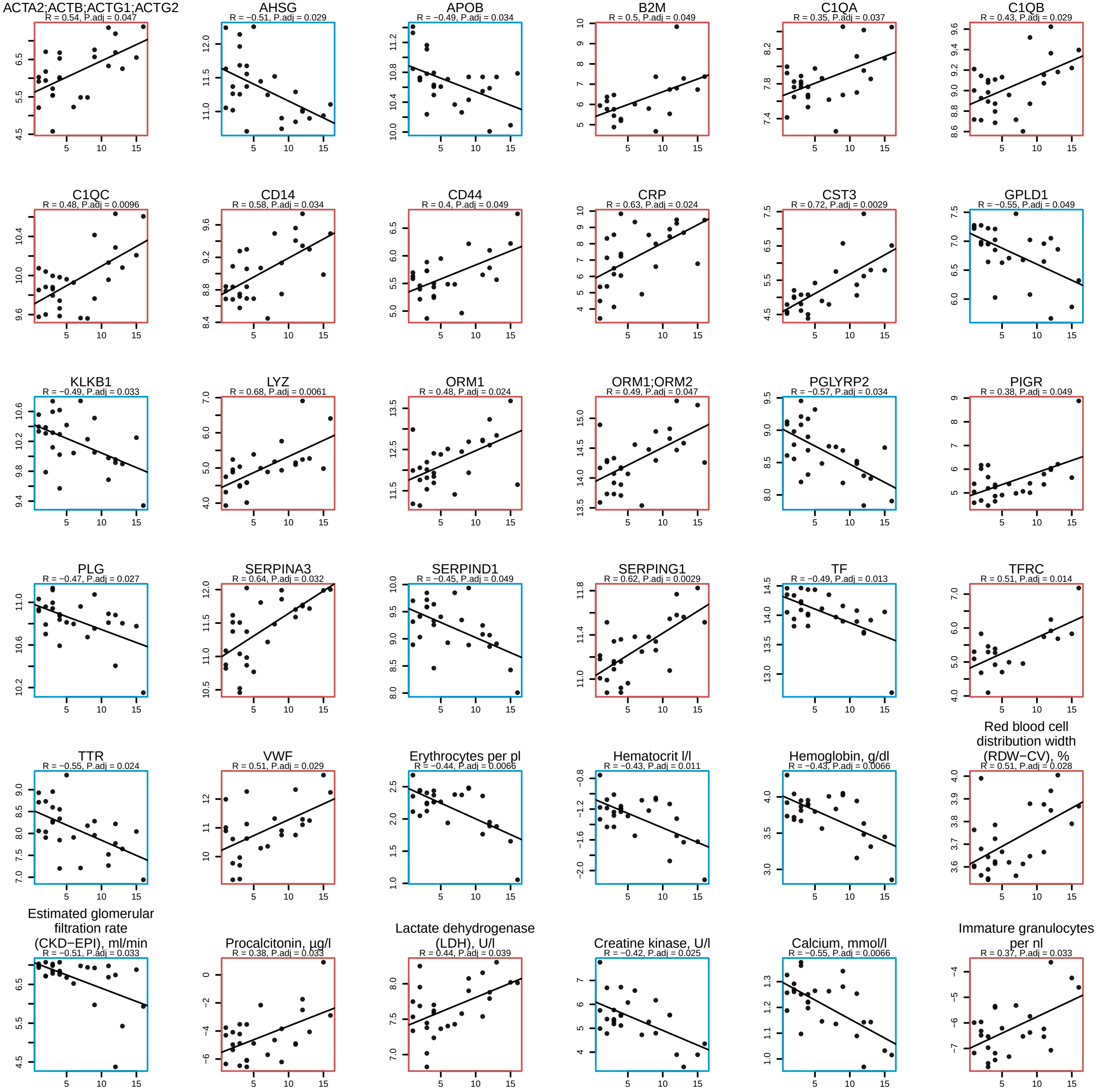

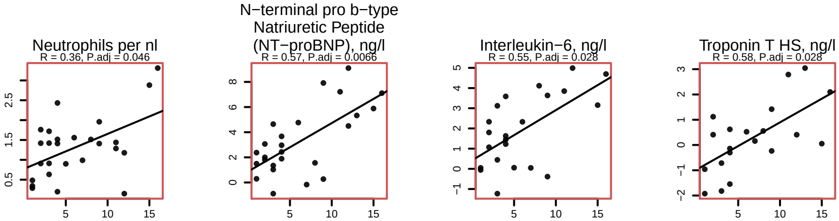

### SF21 - omics features predictive of future disease worsening.pdf

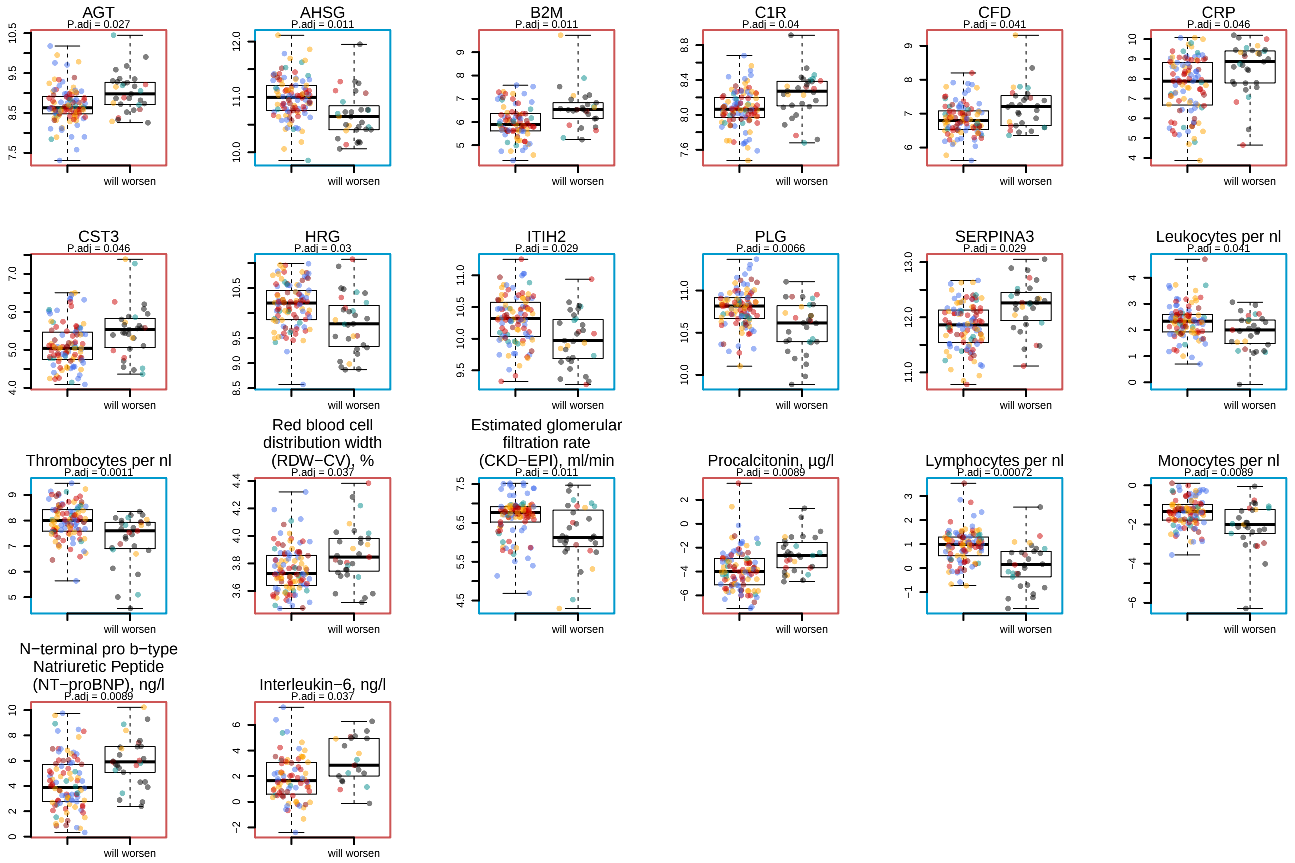

### SF22 - omics features the dynamics of which indicates poor prognosis in WHO = 6 and 7 patients.pdf

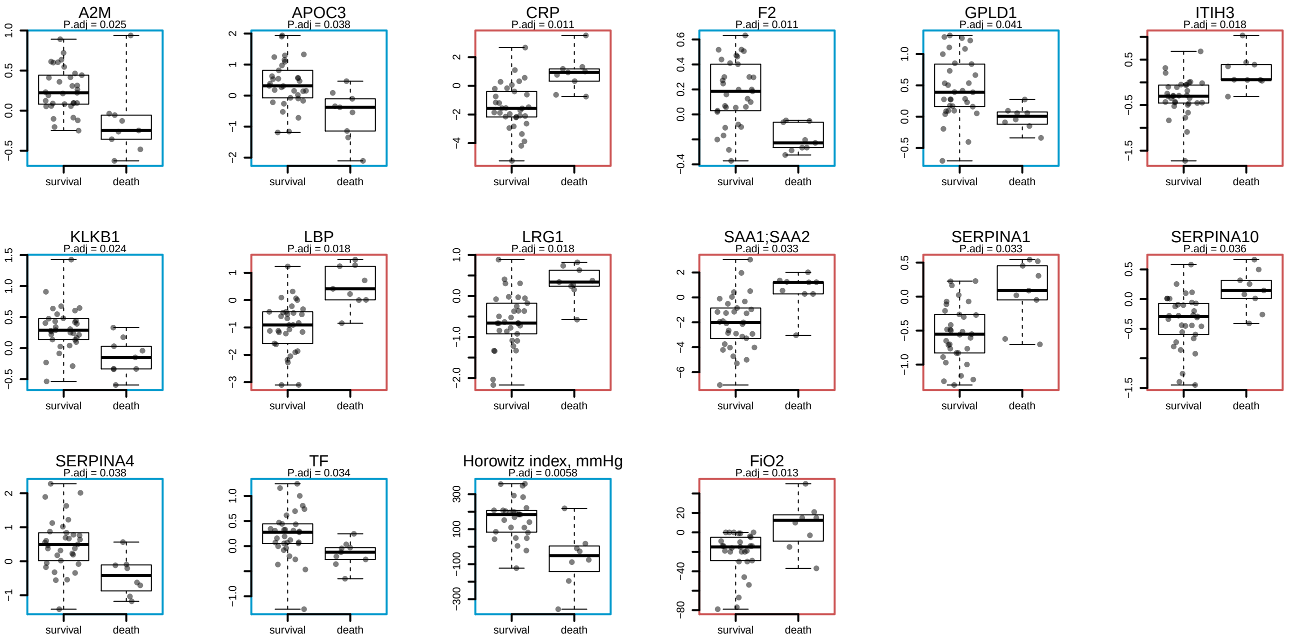
